# Exploring Psychological and Biological Mediators between Childhood Adversity and Psychosis: An updated Systematic Review and Meta-Analysis

**DOI:** 10.64898/2026.07.19.26358426

**Authors:** Geetanjali Kumar, Ines Lepreux, Lora Bici, Fizza Mustafa, Manuel Abella, Giulia Trotta, Monica Aas, Lucia Sideli, James H MacCabe, Ricardo Twumasi, Kelly Diederen, Andrea Mechelli, Morwenna Rickard, Ewan Carr, Whiskey Eromona, Rodolfo Rossi, Natalia E. Fares-Otero, Amy Hardy, Luis Alameda

## Abstract

**Background:** Childhood adversity (CA) has been identified as one of the most robust risk factors for psychotic disorders; several treatable mediating mechanisms have been proposed.

**Aims:** To conduct a systematic review and meta-analysis examining mediating pathways linking CA and psychosis.

**Method:** This PRISMA-compliant systematic review (PROSPERO: CRD42024542972). consisted of a search conducted in January 2026 on Ovid (PsycINFO, Medline, and Embase) using search terms related to psychosis, CA, and mediation analyses. Evidence was appraised by calculating the percentage of the total effect mediated in each study, grouping mediators into meaningful groups. When possible, meta-analyses using two-stage meta-analytic structural equation modelling (METASEM) were conducted.

**Results:** 117 studies were included (54 in clinical samples, 59 in non-clinical samples, and four studies in both clinical and non-clinical samples). 107 studies examined psychological mediators and 12 examined biological. The median percentages of total effect mediated across all analyses per mediator family were: 49% for dissociation (k = 24), 45% for psychosocial stressors (k = 6), 37.9% for negative schemas (k = 23), 35.2% for post-traumatic symptoms (k = 10), 31.5% for depressive symptoms (k = 14), 27.8% for anxiety (k = 11), 27.1% for attachment styles (k = 12), and 8.7% for mentalization domains (k = 5). Meta-analyses confirmed a robust mediating effect of dissociation (k = 7; N = 2143; indirect effect (I.E) =0.42 [0.17, 0.66] on psychosis; 50.49%), on delusions (k =7; N = 1053; I.E = 0.36, [0.27, 0.46]; 46.44%]) and on hallucinations (k =10; N = 5705; I.E = 0.28 [0.20, 0.36]; 57.59%). Robust mediation via depression (k =5; N= 5028; indirect effect= 0.33 [0.31, 0.35]; 31.05%) and negative schemas of the association between trauma and psychosis broadly defined (k = 7; N=10791; I.E= 0.26 [0.17, 0.35]; 26.36%) was also observed. High heterogeneity was observed across all meta-analyses. Fewer studies examined biological mediators, preventing quantitative synthesis.

**Conclusions:** Childhood adversity impacts psychosis through psychosocial mediators, particularly dissociation. Further work is required to on the potential role of biological mechanisms and its interplay with psychological mechanisms.

## 1. Introduction

Psychosis has increasingly been viewed on a continuum, with subclinical psychotic-like experiences in non-clinical samples sharing risk factors and features with clinical disorders.^1,2^ Indeed, psychosis constitutes a major public health concern, emphasising the importance of identifying its potential risk factors and developing early interventions addressing the mechanisms by which they lead to illness outcomes.

Childhood adversity (CA) has been identified as one of the most robust and potentially modifiable transdiagnostic risk factors for mental disorders.^3,4^ The broad use of the term CA is commonly equated with exposure to psychologically threatening events involving abuse (sexual, physical or emotional), neglect (physical or emotional), domestic violence, parental mental illness or drug abuse, harsh parenting, and bullying.^5^ A recent umbrella review showed that CA has the largest population attributable fraction (PAF) associated with psychotic disorders (estimated at 40.69%).^6^ As CA occurs during key developmental periods, these experiences can exert long-lasting effects on biological and psychological functioning.^7,8^ A growing body of research highlights that CA has been frequently associated with an increased occurrence, severity and persistence of psychosis symptoms across the spectrum (including high risk and first-episode).^9,10^ However, it is important to highlight that not all individuals exposed to CA go on to develop psychotic symptoms or disorders;^11^ this reflects the role of additional influences which can mediate the relationship.

Several *psychological mediators* have been proposed to explain this pathway, supported by cognitive models of psychosis.^12–15^ These include the role of negative schemas, dissociation, post-traumatic stress symptoms, and affective dysregulation. In parallel, there has been increasing interest in biological mediators;^16–18^ emerging research points to several mechanisms, including neurobiological alterations, neuroinflammation, and epigenetics.^19^ Thus, understanding these interrelated pathways is essential for developing preventive and tailored interventions for individuals with histories of CA at risk of psychosis or those who developed a psychotic disorder.

To date, only two systematic reviews published in 2020 comprehensively appraised mediational psychological and biological mechanisms linking CA and psychosis.^20,21^ Other similar work has focused only on psychological mediators,^22,23^ with the latest published in 2021. Evidence at this stage points to dissociation, negative schemas (about the self, the world, and others), mood and anxiety as the main replicated pathways mediating trauma and psychosis. More recent reviews have focused on some of these pathways individualy^24–27^ but have not integrated them comprehensively. Only Bloomfield et al. (2021) explored all families of psychological mediators by conducting a meta-analysis that combined estimates from individual mediation analyses. Of note, this approach only meta-analysed the indirect effects reported by individual studies. These estimates are difficult to compare across studies because they are influenced by the strength of the overall association between CA and psychosis. New methods such as meta-analytic structural equation modelling (METASEM),^28^ overcome this limitation by synthesising the complete mediation across studies.

The present review therefore aims to update Alameda et al. (2020) to consider biological and psychological mediators between CA and psychosis in clinical and non-clinical samples. This will include articles from reviews conducted over the past seven years that consider specific psychological pathways,^25–27^ and will additionally include a meta-analysis of the available data, which was not possible in the original search given the limited number of studies.

## 2. Methods

### 2.1. Study protocol

The study protocol was pre-registered with the International Prospective Register of Systematic Reviews (PROSPERO registration number: CRD42024542972, 19 June 2024). The review was reported according to the updated Preferred Reporting Items for Systematic Reviews and Meta-Analysis (PRISMA) guidelines^29^ (See Table S1). To ensure consistency in study identification, this review methodology replicated the approach used by Alameda et al. (2020), to which the protocol has been amended to reflect the quantitative analysis methodology. Any amendment of the protocol is described in the SM.

### 2.2. Search strategy

We conducted an initial systematic search via Ovid (PsycINFO, Medline, and Embase) from January 2019 (date of Alameda et al. (2020) search) to June 2024. Once we screened the number of new papers available up to June 2024, and extracted the available data, we amended the protocol (17 January 2026) adding details on meta-analytical methods and subsequently conducted a search update in January 2026. Following Alameda et al. (2020), the search strategy consisted of keywords relating to (1) psychosis, (2) CA, and (3) mediation terms. We used the boolean ‘AND’ operator (see search strategy and terms inserted in the supplement). Filters were used to remove duplicates. After retrieving these articles, two researchers (GK and IP) independently screened the titles and abstracts and then the eligible full texts of identified articles for relevance. Overall, there was an 85.1% (230/270) agreement and consistency between the two researchers (GK & IP); any discrepancies were resolved by a third researcher (LA). All relevant reviews on the topic were cross-referenced for potential new articles,^20–27,30^ including the 48 studies included in the original search of Alameda et al., 2020, which had cross-referenced one similar review on the topic.^23^

### 2.3. Inclusion and exclusion criteria

In relation to the PECO framework,^31^ studies required (1) (P) for clinical studies of any psychosis diagnosis based upon validated diagnostic manuals and scales, which includes the Statistical Manual for Mental Disorders (DSM) and the International Classification of Diseases (ICD), or psychosis symptoms within the general population such as psychotic-like experiences. Analyses were appraised separately depending on sample type; (2) (E) the presence of CA, defined as having occurred before the age of 18 and measured as aggregated or overall CA or specific CA subtypes (sexual abuse, physical abuse, emotional abuse, physical neglect, emotional neglect, or bullying – see “predictors” section in the SM); (3) (C) compared with individuals without CA within the sample according to (1); (4) (O) assessed for psychological or biological mediators and with validated instruments between CA as detailed above and psychosis outcomes as dependent variable. Studies were required to report a mediation analysis using a robust method testing mediation, allowing the ascertainment of a mediating/indirect effect between the predictor and the outcome. We also considered network analyses for the systematic review, as they explore mediation pathways. However, as this method does not directly report a measure of indirect effect, they were not included in the quantitative synthesis, as in Alameda et al., 2020. In the meditation path, psychosis was broadly defined (e.g positive symptoms or caseness in a case-control study); symptoms subtypes such as delusions and hallucinations. In studies conducted in non-clinical samples, we also considered subthreshold psychotic-like experiences and schizotypal traits; (5) reported original results from a peer-reviewed journal.

Studies were excluded if they: (1) were reviews, case reports, conference proceedings, study protocols, preprints, dissertations, or other grey literature; (2) included samples in which more than 20% of participants were aged over 65 years, to minimise potential confounding by age-related cognitive decline; (3) were conducted exclusively in pregnant women, to minimise potential confounding by pregnancy-related hormonal changes; (4) recruited the entire sample from prison settings, to minimise heterogeneity and potential confounding related to the higher prevalence of antisocial personality traits in these settings; or (5) were not published in English. These criteria did not exclude broader clinical samples that included some pregnant women or participants with a history of imprisonment.

### 2.4. Data extraction and quality assessment

A primary screening of the title/abstract was performed in duplicate. Once the full texts of potentially eligible articles were retrieved, a second screening was performed in duplicate. Any disagreement at either screening stage was resolved through consensus at a team meeting. General methodological data were extracted in duplicate, including: name of first author, year and country of publication; study design and name of cohort; sample size, sex frequencies and range of age (mean/sd); type of analysis and mediation effects.

Information on the nature (i.e., biological pathway or psychopathological dimension) and magnitude of the mediation was extracted, and direct, indirect, and total effects were reported, along with the percentage of the total effect mediated. Other useful information was extracted when available (for example, confounders) and quality assessments were obtained.

An alternative dataset was prepared for meta-analytical purposes (when at least 5 studies were available which examined the same mediator group, same trauma category and the same mediator and outcome). Details of the meta-analytical data collected, extraction procedures, and harmonisation are described in the SM.

Two independent reviewers (GK and IP) utilised the same modified version of the Newcastle-Ottawa Scale (NOS)^32^ to assess quality and risk of bias. This was cross-checked by a third reviewer (LA). Detailed information on the quality assessment procedure and tool is available in the SM supplementary materials.

### Data synthesis and meta-analysis

#### Systematic review

We grouped the results according to meaningful groups of mediators, types of adversity and outcome variables (see details below) of interest. These have been grouped in a descriptive figure plotting the percentage of total effect mediated across the main groups of mediators, as in our original publication.^20^ Analyses excluded from Figure 1 (e.g., due to insufficient information to calculate the percentage mediated, non-comparable analytical approaches) are listed in Table S3. Additionally, tables in both clinical and non-clinical populations reporting the % of studies and analyses showing evidence of mediation have been presented as in the original publication. The percentage of total effect mediated is the outcome of interest in the systematic review. The methodological characteristics and findings of each included study have been presented narratively, along with a descriptive table (SM).

#### Meta-analysis

We then computed random-effects models when at least five studies (as previously recommended) have examined the same mediator group^33,34^ the same trauma category, and the same mediator and outcome; additional details on meta-analytical proceedings can be found in SM.

Meta-analyses were conducted in R, following the meta-analytical structural equation modelling (METASEM) methodology—specifically, the two-stage structural equation modelling approach (TSSEM)^28^ —used in similar studies.^24^ This method allows for multiple correlational relationships to be synthesised simultaneously. TSSEM proceeds in two steps: first, pooling correlation matrices from included studies using a random-effects model with a diagonal variance-covariance structure; and second, constructing a structural equation model from the pooled correlations, with likelihood-based 95% confidence intervals. This method overcomes the issue of aggregating mediating effects without accounting for the total effect present in the previous meta-analysis.^22^ The R code used is shared in SM.

Publication bias and small study effects were assessed by examining the asymmetry in the funnel plots and with the Egger’s Regression when more than 10 studies are available.^35^ The presence of heterogeneity was evaluated using the Q-test, and its magnitude were assessed using the I² statistic. Sources of heterogeneity were examined by conducting sample type subgroup analyses (clinical vs. non-clinical) when at least five studies were available per group.

## 3. Results

117 studies met our inclusion criteria and were included in this review, out of the 3753 studies identified in the search (see flowchart; see SM for search strategy). 107 explored psychological mediators and 12 biological mediators. In total, 59 studies were conducted within the non-clinical samples, 54 in clinical samples, and 4 reported findings for both clinical and non-clinical samples (see the full list of included studies in the SM). See Table S4 for more details on clinical samples. The total number of subjects in the non-clinical studies was 135,525 (mean age = 32.81 years, SD = 17.57 years [range = 11–83], 62% females); the total number of subjects from clinical studies was 13,772 (mean age 32.96 years, SD = 10.28 years [range = 13-71], and average % female 43.4%). Table S5 describes the groups of mediators created for this review with the cluster of mediating pathways within each family. Meta-analyses were possible for the following mediators: dissociation, negative schemas and depression.

Several studies contained multiple analyses (e.g. examining different mediators, psychosis outcomes or trauma subtypes). Each analysis was considered separately in the descriptive synthesis. In total, 318 analyses were included in the review from non-clinical studies; the number of analyses in each study ranged from 1 to 90^36^ (median = 2). In total, 191 analyses were included in the review from clinical studies, ranging from 1 to 30 per study^37^ (median = 1). Collectively, eight studies were excluded from the total number of analyses due to the high number of pathways tested.^38–45^ These studies employed network analytic approaches which did not permit the identification of discrete mediation analyses comparable to those reported in the remaining studies. These studies would have distorted the numerical summaries and therefore are described narratively but are not considered in the summary tables, consistent with the approach adopted by Alameda et al (2020) (see table S6 for individual reasons of exclusion).

Four studies were rated as ‘good’ on the NOS quality assessment scale.^46–49^ All other studies were rated as ‘fair’. Cross-sectional research designs limit the ability to establish temporal ordering, which reduces methodological robustness when establishing the biological and psychological mediators between CA and psychosis. There was 93.8% consistency between the two authors GK and LP in their ratings. See Table S7, 8, 9 & 10 for individual NOS scores.

### Psychological mediators

#### Dissociative symptoms

In total, 25 studies explored the mediating role of dissociation in the relationship between childhood adversity (CA) and psychosis. 16 were included in the meta-analysis. For descriptive synthesis, an analysis was considered to support mediation if it reported a statistically significant indirect effect for the proposed pathways between CA and psychosis outcomes. Overall, 64.4% of analyses supported mediation via dissociation (47/73 analyses). Support for mediation was found in 48.8% of analyses conducted in clinical populations and 84.4% of analyses in non-clinical samples (See Table 1). One network analysis looked at dissociation alongside negative schemas and attachment.^44^

**Table 1:** Number and percentage of mediation analyses supporting mediation across clinical and non-clinical samples by mediator category.

|  | Clinical samples<br>Number of analyses<br>(number of studies) |  |  | General population<br>Number of analyses<br>(number of studies) |  |  |  |
| --- | --- | --- | --- | --- | --- | --- | --- |
| Category<br>(N total analyses/studies) | Evidence of mediation | Null mediation | % analyses supporting mediation | Evidence of mediation | Null mediation | % analyses supporting mediation | Total % analyses supporting mediation |
| Dissociation (73/25)<sup>1</sup> | 21/8 | 21/8 | 48.8% | 27/14 | 5/4 | 84.4% | 64.4% |
| Negative Schemas (78/23) | 14/8 | 29/5 | 31.8% | 22/11 | 8/4 | 64.7% | 46.1% |
| Other PTSD (19/10) | 6/4 | 1/1 | 85.7% | 8/5 | 4/3 | 66.6% | 73.7% |
| Depression (27/14)<sup>2, 3</sup> | 4/2 | 3/3 | 57.1% | 11/6 | 10/6 | 52.4% | 53.6% |
| Anxiety (42/11)<sup>4, 5</sup> | 7/3 | 19/3 | 26.9% | 7/3 | 9/4 | 43.8% | 33.3% |
| Attachment (41/12)<sup>6</sup> | 6/4 | 7/4 | 46.2% | 20/7 | 6/2 | 71.4% | 63.4% |

<sup>1</sup> One study in the clinical population conducted a serial mediation as well as individual mediation analyses. Both analyses were included (Pearce et al., 2017)
<sup>2</sup> Ostefjells et al looked at both anxiety & depression as one mediator which was excluded from this analysis
<sup>3</sup> This excludes the network analyses -Huang et al. 2022 and Jin et al., (GP studies)
<sup>4</sup> Ostefjells et al looked at both anxiety & depression as one mediator which was excluded from this analysis
<sup>5</sup> This excludes the network analysis by Jin et al., 2022 (GP Study)

|  |  |  |  |  |  |  |  |
| --- | --- | --- | --- | --- | --- | --- | --- |
| **Mentalis**<br>**ation**<br>**(93/5)** | - | 3/3 | 0% | 34/1 | 56/1 | 37.7% | 36.6% |
| **Psychos**<br>**ocial**<br>**Stressor**<br>**s (8/6)<sup>7</sup>** | 2/2 | - | 100% | 6/3 | - | 100% | 100% |
| **Biologic**<br>**al**<br>**Mediator**<br>**s (22/12)** | 9/6 | 6/4 | 60% | 1/1 | 6/3 | 14.2% | 45.4% |

<sup>7</sup> This excludes Goldstone et al., 2011; Goldstone et al., 2012 & Qjao et al., 2023) due to a high number of analyses/multiple pathways.

The median percentage of total effect mediated by dissociation was 49% (See Fig 1). Dissociation was examined particularly in relation to composite measures of CA, followed by abuse. In addition, hallucinations were the most reported outcome when examining the mediating role of dissociation between CA and psychosis. The meta-analytical findings showed that dissociation explained 50.5% of the association between trauma and psychosis broadly defined (k=7; N=2143; indirect effect=0.42 [0.17, 0.66]), 46.4% of delusions (k=7; N=1053; indirect effect=0.36, [0.27, 0.46]) and 57.59% of hallucinations (k=10; N=5705; indirect effect=0.28 [0.20, 0.36]). One study removal did not show a major change in overall indirect effects. Heterogeneity was overall high across the associations between the predictor and the mediator (‘a”), the mediator and the outcome (“b’) and the predictor and the outcome (‘c’) ((see pooled correlation matrix tables in SM).

#### Negative schema

There were 23 studies which explored the mediating role of negative schemas between CA and psychosis. 12 were included in the meta-analysis. The total percentage of analyses supporting evidence for negative schemas was 46.8% (36/77 supported evidence for a mediating effect). One network analysis looked at negative schemas alongside dissociation and attachment.^44^

The percentage of analyses supporting mediation was notably higher in non-clinical samples (64.7%) compared to clinical studies (32.6%). The median percentage of total effect mediated by negative schemas was 37.9%. A composite measure of CA was the most reported predictor of significant findings. Psychosis was the most reported outcome, followed by delusions and caseness (See Fig. 1). (see Table 1). The meta-analytical findings showed that negative schemas explained 26.5**%** of the association between trauma and psychosis broadly defined (k=7; N=10791; indirect effect= 0.26 [0.17, 0.35]), 27.4% of delusions (k=7; N=1053; indirect effect= 0.26 [0.13, 0.39]) One study removal did not show a major change in overall indirect effects. Heterogeneity was overall high across the associations between the predictor and the mediator (‘a”), the mediator and the outcome (“b’) and the predictor and the outcome (‘c’) ((see pooled correlation matrix tables in SM).

#### Posttraumatic stress symptoms

10 studies examined the mediating role of other post traumatic stress disorder (PTSD) symptoms in the relationship between childhood adversity (CA) and psychosis. Overall, 73.7% of analyses supported mediation (14/19). Of note, the percentage of significant analyses was notably higher in clinical samples (85.7%) compared with non clinical samples (66.6%). The median percentage of total effect mediated by ‘other PTSD’ was 35.2%. Abuse was the most reported predictor (see Fig. 1).

#### Depressive and anxiety symptoms

Studies examining the mediating roles of depression and anxiety were reported separately in Fig. 1 and Table 1. Depression was investigated in 14 studies; 7 were included in the meta-analysis. 53.6% (15/28) of these analyses supported mediation (see Table 1). There were similar findings across samples: 57% in clinical and 52% in non-clinical samples. For Depression, the median percentage of total effect mediated was 31.5%. All studies reported in Figure 1 reported psychosis as an outcome. Bullying was the most reported predictor, followed by composite CA. The meta-analytical findings showed that depression explained 31% of the association between trauma and psychosis (k=5; N= 5028; indirect effect= 0.33 [0.31, 0.35]). One study removal did not show a major change in overall effects (SM). Heterogeneity was overall high across the associations between the predictor and the mediator (‘a”), the mediator and the outcome (“b’) and the predictor and the outcome (‘c’) (see pooled correlation matrix tables in SM).

Anxiety was examined in twelve studies in total 33.3% of these analyses supported evidence for mediation (14/42). The median percent of total effect mediated was 27.8%, with psychosis and composite as the most frequently reported predictor and outcome variables (see Fig. 1). One study looking at the role of depression and anxiety collectively as a mediator between emotional abuse and psychosis failed to report significant findings.^50^ Two studies looked at both depression and anxiety as one mediating variable, reporting mixed findings (3/4 reporting evidence of mediating effects).^50,51^ One network analysis looked at the role of SA on depression and anxiety along with PTSD in college students.^43^

#### Attachment style

In total, 14 studies explored the mediating role of attachment styles between CA and psychosis. The total percentage of analyses supporting evidence was 63.4%, with 26/41 reporting a mediation effect. Two network analyses looked at attachment alongside other psychological mediators.^44,45^ The median percentage of total effect mediated by attachment was 27.1% (See Fig. 1). Both composite and abuse were the most frequently reported predictors, whilst delusions were the most common outcome (see Fig. 1).

#### Psychosocial stress

There were 8 studies which explored the mediating role of various psychosocial stressors between CA and psychosis. The total percentage of analyses supporting evidence for psychosocial stressors was 100% (8/8 studies supporting evidence for a mediating effect). Three studies including network analyses could not be synthesised quantitatively.^39,40,45^ Of the three, two studies examining the mediating role of life hassles reported significant suggested mediation effects between emotional and sexual abuse on hallucinations and delusions.^39,40^ The median percentage of total effect mediated by psychosocial stressors was 45%.

#### Mentalisation

Only five studies explored the mediating role of mentalisation. The total percentage of analyses supporting evidence was 36.6% with 34/93 analyses supporting evidence of a mediation effect.^36,50^ The median percentage of total effect mediated by mentalisation was 8.7% (See Fig. 1). It is important to note that these figures are heavily influenced by a single study, which contributed 90 analyses in total.^36^ One path analysis with multiple analyses reported a significant role of metacognitive beliefs as a suggested mediation between emotional and sexual abuse on hallucinations.^40^

#### Affective dysregulation

In total, three studies in the general population examined the mediating role of affective dysregulation.^52–54^ All the studies reported mixed findings. 4/8 studies reported evidence for significant mediating effects between composite CA and psychotic experiences. All three studies reported a mediating effect on at least one of their analyses.

#### Cognitive biases

In total seven studies in the general population looked at other cognitive biases. Three of them, setting from the sample examined a composite score of the Davos Assessment of Cognitive Biases Scale (DACOBS), a measure including a broad range of bias (subjective cognitive problems, safety behaviors, attributional biases and social cognition problems.^55–57^ More specifically, attributional styles such as locus of control appeared in three additional studies.^58–60^ One study examined interpretation bias.^61^ Most of the analyses supported the role of a mediating effect between childhood adversity and psychotic-like experiences in non-clinical samples (13/15).

#### Loneliness

Similarly, five studies examined the role of loneliness, with the majority conducted in non-clinical samples^45,62–64^ and one in clinical samples.^65^ Most of the analyses supported evidence for loneliness as a mediating variable between CA and loneliness (5/7).^62–65^

#### Sleep disturbances and fatigue

Four studies explored whether sleep disturbances mediate the relationship between CA and psychosis,^66–69^ which collectively reported mixed findings, with 5/6 analyses supporting evidence for mediation.

#### Anomalous experiential processes

Similarly, one study found significant effects when anomalous experiences were used as a mediator.^70^ Significant evidence was also reported for persecutory ideation^69^ and negative voice content^71^ and aberrant salience^68,70^ as mediating variables.

#### Personality features

Three studies reported significant evidence that borderline personality disorder symptoms are a mediating variable (5/5 analyses).^72–74^ One study looked more broadly at personality and similarly reported evidence for mediating effects between bullying and psychotic experiences.^61^ One study reported that personality organisation played a significant mediating role between composite CA and paranoid thinking.^75^

#### Positive attributes

Some mediating variables were measuring whether positive attributes mediated the relationship between CA and psychosis. In sum seven studies looked at positive attributes, more specifically, wisdom,^76^ resilience,^57,77,78^ mindfulness,^79^ motivation^41^ and positive attributes as a whole.^46^ All studies reported significant effects (6/9) except for resilience, which reported mixed findings^78^ and mindfulness^79^ which failed to report evidence for a significant mediation. With the exception of Metel et al. (2020), which reported unexpected positive indirect effects of resilience, studies suggested that CA increased psychosis risk by a reduction in positive attributes.

#### Other psychological processes

The psychological mediators not included in the aforementioned categories reporting significant mediating effects include IQ,^80^ psychosocial functioning,^81^ threat anticipation,^82^ cognitive functioning^83^ and common mental health disorders.^77^ Studies reported mixed findings for perceived injustice,^84^ OCD symptoms,^85^ social defeat,^54,86^ and interpersonal sensitivity.^87,88^ Other psychological mediators, which failed to report significant evidence for mediating effects, were social comparison,^63,89^ belief updating processes,^90^ mood swings, mania,^91^ intelligence,^83^ parent child conflict and feelings of safety.^52^

Four studies used network analytic approaches to examine the potential role of a range of psychological mechanisms.^38,41,42,45^ Across these studies, the networks consistently indicated multiple interconnected pathways, making it difficult to isolate the influence of single suggested mediators. See table S7,8, 9 & 10.

#### Biological mediators

Altogether, twelve studies explored the role of biological mediators between CA and psychosis.^16,17,19,83,92–99^ 45.5% (10/22) of the analyses supported evidence for mediation. See descriptive table S8 & 10 in the SM. Four studies examined neurochemical function, with only 1/4 studies providing evidence for a significant mediation for D2/3 availability,^17^ whilst the remaining 3/4 failed to find evidence to support significant mediations for glutamate/glutamine^94,95^ or dopamine release.^16^ Additionally, three studies explored the mediating role of brain structure and activation; more specifically, grey matter volume in the dorsolateral prefrontal cortex (DLPFC),^93^ brain activation in the inferior frontal gyrus activation,^96^ and brain activation in the right middle temporal gyrus,^97^ The latter two studies failed to obtain significant findings. One study examined the mediating role of sensory gating deficits (P50);^98^ significant findings were observed when emotional neglect was used as a predictor, but not for physical neglect. Two biological studies examined the role of molecular markers, specifically brain-derived neurotrophic factor (BDNF)^99^ and DNA methylation as an epigenetic process.^19^ Wang et al (2022) found significant findings for both composite and emotional neglect as predictors of the mediating role of BDNF levels on psychosis. Similarly, Alameda et al (2023) reported significant evidence on the mediating role of DNA methylation on psychosis when composite, abuse and neglect were entered as predictors. Additionally, one study looking at biological biomarkers reported significant mediation effects^92^. No meta-analysis could be conducted on biological mediators given heterogeneity in the available pathways. Finally, one study exploring the mediating role of IL6, IL6R, body mass index (BMI), and cortisol levels failed to find significant mediating effects between composite CA and schizophrenia.^83^ Meta-analytical analyses could not be conducted on biological studies.

## 4. Discussion

Our review provides a comprehensive update on the evidence for psychological and biological mechanisms linking CA and psychosis across the spectrum. We appraised 117 individual studies, covering 15 putative mediating mechanisms between CA and psychosis, categorised into conceptual groups (see Table S5). These were classified based on the percentage of total effect mediated in the association between CA and psychosis across the spectrum, where possible. Compared with the most recent review from our group, which included 48 studies up to 2019,^20^ the current review reflects continuous growth in the evidence base (Figure 2). This growth highlights the increasing recognition of CA as a key factor in psychosis and provides a stronger empirical foundation towards understanding its underlying mechanisms. Research in biological processes is comparatively slower as compared with psychological mechanisms.

Overall, the findings suggest dissociation is the most consistent and strongest mediator of the trauma and psychosis relationship, particularly with hallucinatory experiences. This conclusion was supported by both our meta-analysis and SR findings. Dissociation has been hypothesised to impact psychosis through compartmentalisation of unresolved or unbearable emotions as a survival response to persistent threats.^100^ Another putative pathway is through detachment in the form of depersonalisation or derealisation leading to disruptions in sensory-perceptual processing and subsequent anomalous experiences.^101,102^

Negative schema or beliefs had the second greatest influence on the trauma and psychosis association; this was confirmed with meta-analytical data. Consistent with cognitive models of psychosis, this suggests that CA shapes the sense of self, others and the world. Themes of defectiveness and vulnerability could play a role in the content and appraisals of anomalous experiences, which may contribute to hallucinations and delusions.^13,15,26^ This finding is also supported by evidence that modifying trauma-related beliefs during trauma-focused therapy mediates improvements in paranoia.^103^

PTSD symptoms accounted for a similar amount of variation in the trauma and psychosis relationship as negative schemas and beliefs. However, this finding was based on a smaller number of studies and some equivocal findings, possibly due to measurement issues across studies. Broadly, PTSD reflects emotional regulation (e.g. avoidance and hyperarousal in relation to trauma-related stimuli) which arises to manage emotionally laden decontextualised memories and beliefs that are characterised by a sense of current threat. These threat-focused cognitive processes may increase vulnerability to anomalous experiences, thereby influencing the content, appraisal, and response to these experiences. Unique to PTSD is also a proposed causal role for fragmented sensory representations that lack temporal and contextual integration in episodic memory, which may manifest as intrusions that are attributed externally.^14,15,104^

Attachment insecurity accounted for slightly less variance in the CA-psychosis relationship than PTSD, making it the fourth strongest mediator identified. Notably, studies which focused on abuse and hallucinations were more strongly mediated by attachment insecurity. It is important to highlight that attachment insecurity also overlaps conceptually with dissociation and negative schemas; they can be viewed as representing ‘internal working models’ (i.e. relational representations of self and others) or emotional and interpersonal regulation (e.g. avoidant, preoccupied or disorganized attachment styles).^105^ The biased metacognitive processing and mentalization linked to insecure attachment styles is proposed to increase the risk of psychosis.^106–108^

Consistent with the affective pathway to psychosis, and the mediating role of emotional regulation, anxiety and depression also partially accounted for the trauma and psychosis relationship; the former confirmed by meta-analytical data. Notably, our review extends the initial work by also identifying ‘psychosocial stressors’ as an additional core mediator. This supports the presence of a second hit hypothesis, where recent stressors, and stress sensitivity may mediate the link between trauma and psychosis. Collectively, this lends further support to the affective pathway to psychosis, which proposes that CA may disrupt emotional regulation systems, resulting in persistent negative affect and increased reactivity to daily stressors, which may exacerbate psychosis experiences.^109,110^

It is also important to acknowledge the role of emerging mediators which were less frequently examined but may still be theoretically and clinically relevant. These include loneliness, cognitive biases, anomalous experiential processes, and personality features. Furthermore, protective cognitive resources warrant consideration alongside vulnerability-focused mediators.^111^ Another promising line of research concerns mentalisation abilities, which may be disrupted in childhood and have been associated with neglect.^106^ Interestingly, none of the mediation studies examining mentalisation examined neglect as a CA subtype, representing a notable gap within the literature. Evidence for biological mediators has evolved significantly from two studies included in Alameda et al. (2020) to twelve studies in the current work. These cover a broad range of biological processes across individual studies and therefore require replication.

In understanding the trauma and psychosis relationship, it is crucial to highlight that these core and emerging mediators are unlikely to operate in isolation. Disentangling the processes by which mediator families impact psychosis is therefore complex. Accordingly, psychological models of trauma in psychosis are multifactorial and, although they have different emphases, largely implicate a role for threat-focused cognitive processing, memories, beliefs and emotional and interpersonal regulation in shaping the phenomenology of psychosis.^14,104,112,113^ Future research should seek to explore the specificity of pathways from trauma types to specific mechanistic processes and particular symptoms, as well as their overlaps.

### Strengths and Limitations

Whilst the findings of this review are consistent with previous studies, there are significant methodological limitations in the review and evidence base. Due to a lack of homogeneous variables across mediating pathways, we were unable to provide quantitative findings for multiple groups of mediators in the form of a meta-analysis. However, where sufficient homogeneous data were available, the use of meta-analytic methods to examine constructed mediator families represents a strength of this review. One limitation is that the CTQ doesn’t encompass a broad definition of CA, including bullying or non-interpersonal experiences. Additionally, CTQ and other measures fail to offer insights into CA timings, chronicity, and multiplicity.^114^ Furthermore, most studies scored lower on the NOS scale (See Tables S7, 8, 9 & 10) due to the use of a cross-sectional research design; this precludes causal conclusions.^115^ Prospective and longitudinal research designs should be conducted to strengthen the robustness of causal inferences made. Incorporating methodologies such as mediation analysis within a latent growth curve model framework will support the mapping of developmental trajectories of mediation pathways.^116^

Regarding the review methodology, whilst the mediator families were based on current constructs and previous work,^20,22,23^ these were somewhat subjective or comprised of relatively heterogeneous constructs. Thus, caution is warranted when interpreting these findings, as they relate to blurred boundaries inherent in the relative simplification of complex and heterogeneous constructs. Future research should also explore mediating variables between CA and negative symptoms, which were not included in this review. Finally, the percentage of total effect mediated should be interpreted with caution, as these estimates may partly reflect differences in study design, measurement approaches, model specification, and the magnitude of the total effect.

### Clinical implications

A summary of possible interventions depending on the mechanisms involved, including biological, can be found in Figure 3. Clinically, the findings of this review highlight the importance of assessing trauma-related mechanisms and, where present, target them through interventions to reduce the onset of psychosis or promote recovery.^117^ As noted, trauma-related mechanisms and outcomes will likely have complex interactions in any given individual, and it is therefore essential that clinicians adopt a flexible and integrative approach. Treatment priorities should be informed by an individualised biopsychosocial formulation, with interventions tailored to the predominant clinical feature.^118^ Figure 3 provides examples of therapeutic techniques that may be used to target specific mechanisms, and how trauma-informed care can be applied across them,^119^ along with medication. Of note, there are significant commonalities among the cognitive, behavioural, and experiential techniques employed in structured therapies and trauma-informed care; therefore, different therapies may modify the same target mediator. Further research is needed to establish if modifying target mechanisms does improve psychosis as hypothesised.

Multidisciplinary staff can support low-intensity trauma-informed interventions, such as psychoeducation about the role of trauma effects in psychosis. This validates and normalises symptoms, thereby reducing unhelpful appraisals and associated distress. For those willing to engage in in-depth work, we recommend prioritising an assessment of PTSD and consideration of trauma-focused therapy if clinically significant PTSD symptoms are identified, consistent with NICE.^120^ Trauma-focused therapies uniquely support reprocessing of trauma memories and associated schema, thereby reducing problematic emotional regulation strategies and their consequences. The STAR trial demonstrated that TF-Cognitive Behavioural Therapy can be integrated with Cognitive Behavioural Therapy for psychosis to safely and effectively treat PTSD, emotional disorders, and psychosis symptoms including delusions, paranoia and multimodal hallucinations in comorbid schizophrenia-spectrum disorders and PTSD.^121^ Notably, the STAR intervention did not lead to significant improvements in depersonalisation/derealisation, despite these emerging as relevant mediators of voice-hearing outcomes. Future interventions may benefit from incorporating a stronger focus on dissociative symptoms alongside trauma-related beliefs and appraisals. Research investigating EMDR for psychosis has also shown promise for PTSD and paranoia, with other multisite RCTs underway.^122–124^ Established (e.g. CBTp) and emerging (e.g. DBT, ACT, CFT, Talking with Voices, AVATAR) trauma-informed therapies for psychosis should also be offered when indicated.^117^

From a biomedical perspective, given the limited findings on the biological mechanism, recommendations beyond the use of antipsychotics should be made cautiously, as based on limited, indirect, and mostly extrapolated from PTSD and depression-only populations. Compounding this limitation, CA is rarely assessed in pharmacotherapy trials for psychotic disorders, representing a methodological gap that constrains evidence-based guidance for this population.^125^ Given the magnitude of the mediating effect observed by anxiety and depressive symptoms, using antidepressants, such as SSRIs, along with antipsychotics in those with trauma and psychosis and a mood component seems reasonable, as previously recommended.^126^ This could complement trauma-focused therapy or offer an alternative to those who are unable to engage. In those with comorbid PTSD, the latest network meta-analysis including 30 RCTs on pharmacological interventions reported that Paroxetine was the preferred choice in terms of balance between overall symptom management, management of individual sub-symptoms, and acceptability in the treatment of PTSD, followed by venlafaxine.^127^ Interestingly, Quetiapine appeared as a reasonable choice for those presenting with re-experiencing and hyper-arousal symptoms, making it a good candidate for those with PTSD and psychosis, who may also benefit from its antidepressant properties, along with Olanzapine and Risperidone, which present the strongest evidence overall across all PTSD domains. Risperidone also ranks the second best for anxiety symptoms in those with PTSD, while Olanzapine and Quetiapine rank first and second, respectively, in efficacy for depression in those with PTSD. More mixed or marginal evidence suggests that propranolol may improve trauma reactivation and can be beneficial for physical manifestations of anxiety, which it is often prescribed off-label.^128^ On the other hand, Prazozin may help with nightmares.^129^ Contrary to the field of psychological interventions, there are no RCTs testing the efficacy of these pharmacological agents in individuals with severe trauma and psychosis, or with comorbid PTSD and psychosis, highlighting an urgent need for future research.

## Conclusions

In conclusion, this review provides a comprehensive update of biopsychological mediators of the trauma and psychosis relationship. It adds further support that CA may impact psychosis through schematic beliefs and memories, and their impact on emotional and interpersonal regulation, via threat-focused processing such as dissociation. Furthermore, although a growing body of research has examined a range of biological mediators, this remains limited compared with the considerably greater attention given to psychological mechanisms, highlighting the need for further research in this domain. The review identifies potential mechanisms for targeted treatment, although a better understanding of how to modify them is required. Further, research should seek to identify what works best for whom so that mental health services can better tailor trauma-informed care to patient needs. It also represents a call for collaborative initiatives that bridge the gap between the psychosocial and biomedical fields to develop new therapeutic targets for individuals with severe trauma and psychosis, examining the interplay between biological and psychological mechanisms jointly. We hope this review stimulates collaborative work combining both.

## Data Availability

All data produced in the present study are available upon reasonable request to the authors

## CRediT authorship

Conceptualization: LA, GK, AH; Data curation: LA, GK, IP,LB, NF, RR ; Formal Analysis; GK, LA, IP,LB, NF, RR; Funding acquisition: LA; Investigation: GK, LA,IP,LB, NF, RR; Methodology: LA ; Project administration: LA, GK ; Resources: ; Software: RR, RT, NF, LA; Supervision: LA, AH; Validation: LA, RR, NF,RT; Visualization:EC, IP ; Writing – original draft: GK ; Writing – review & editing: IP,LB, FM,MA, GT,MA, LS, JM, RT, KD, AM, ED, MR, WE, RR, NF, AM, LA

## Declaration of COIs

No COI to declare

## Acknowledgments

EC is part-funded by the National Institute for Health and Care Research (NIHR) Biomedical Research Centre (BRC): Maudsley.

---

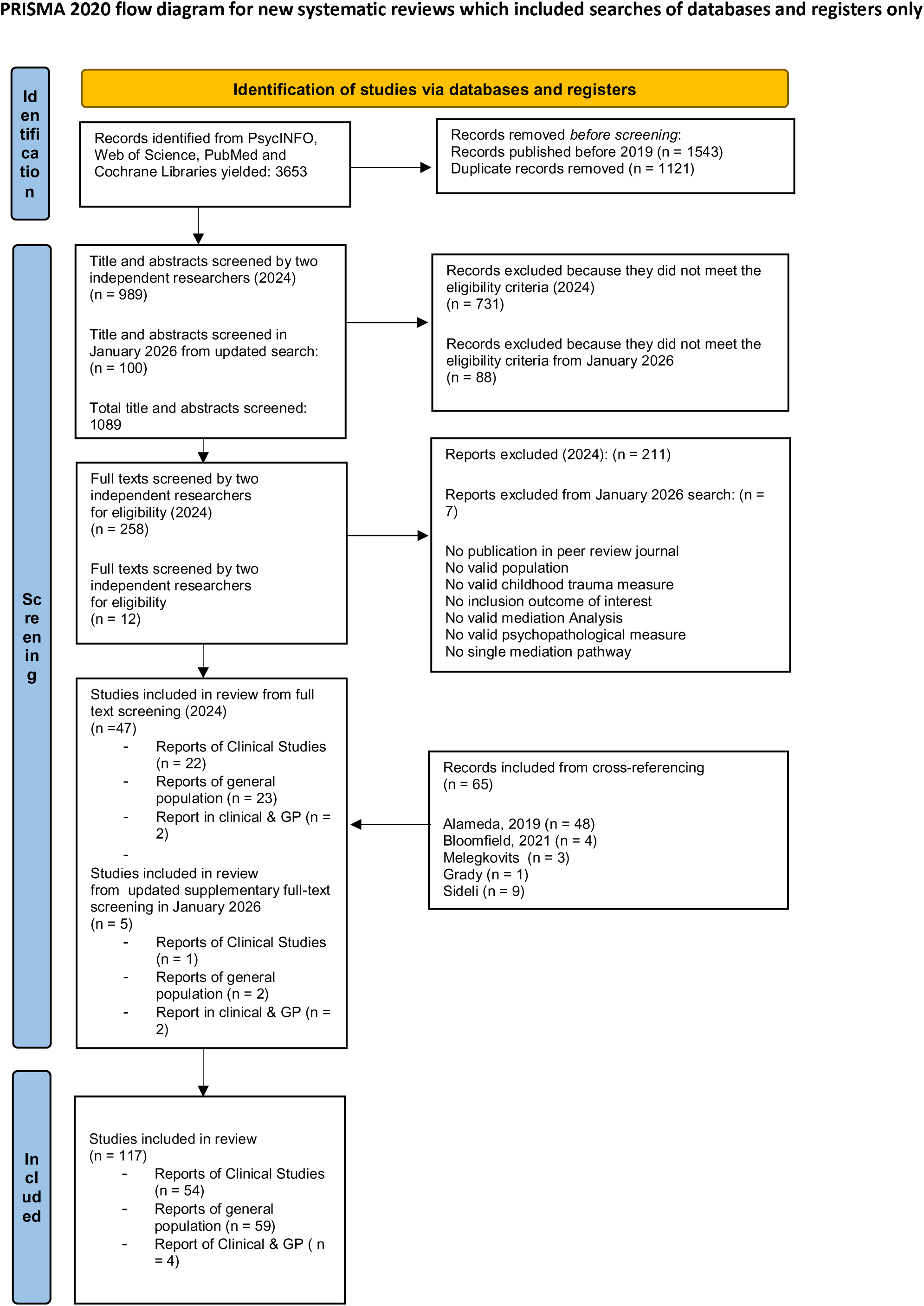
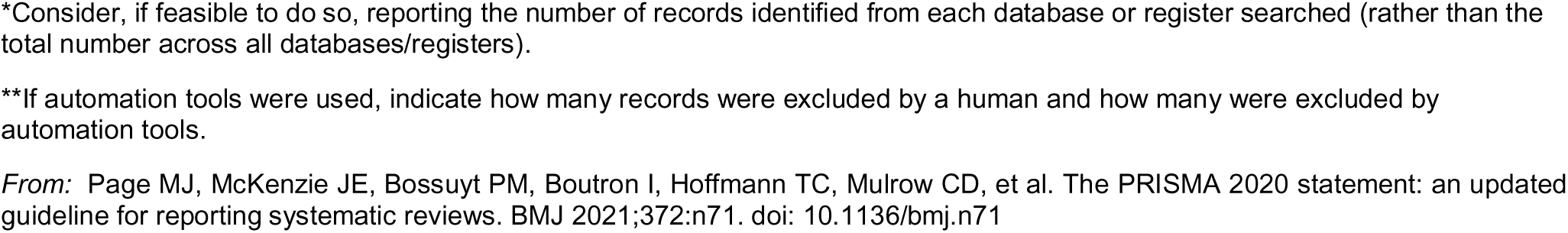

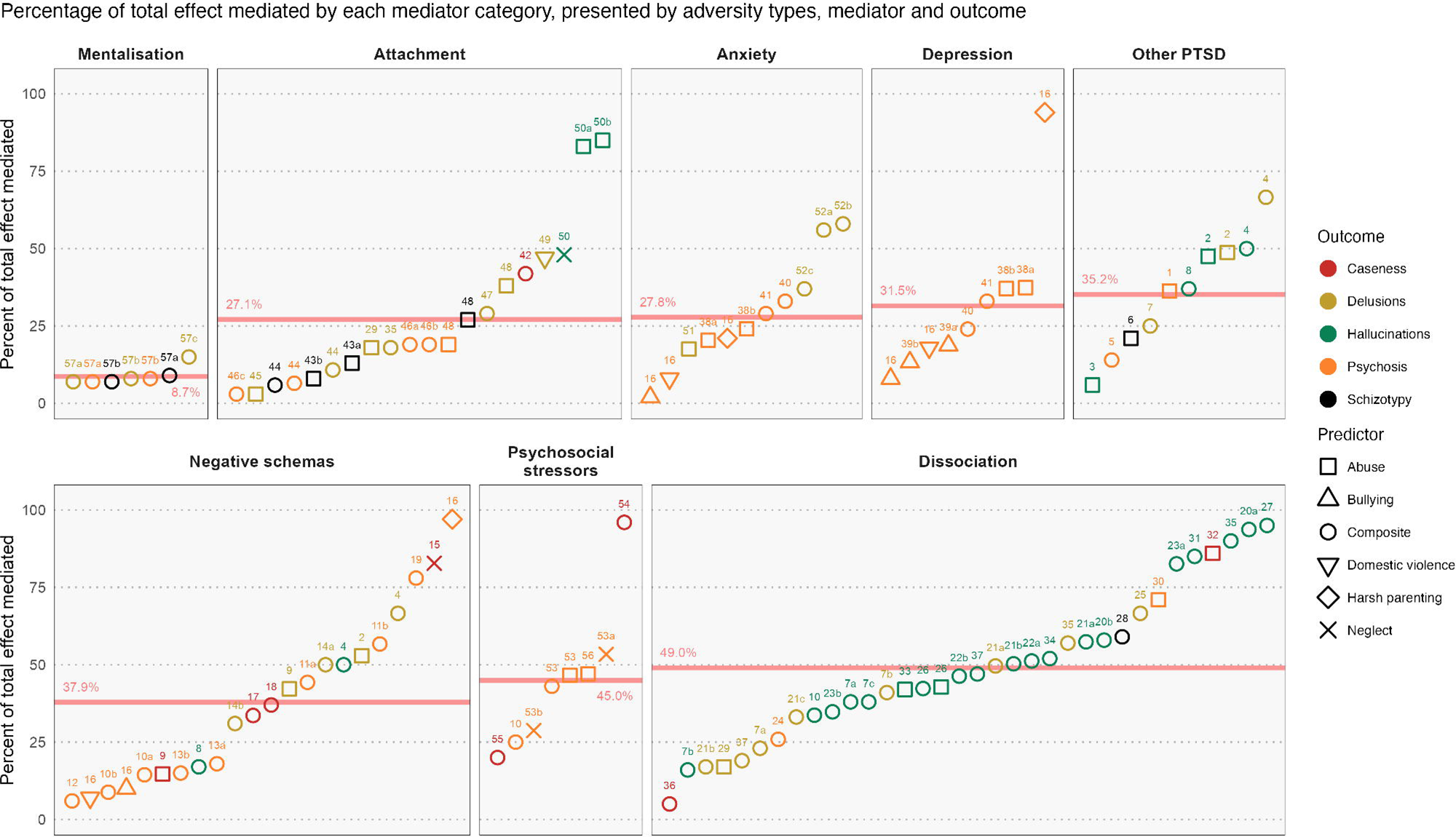

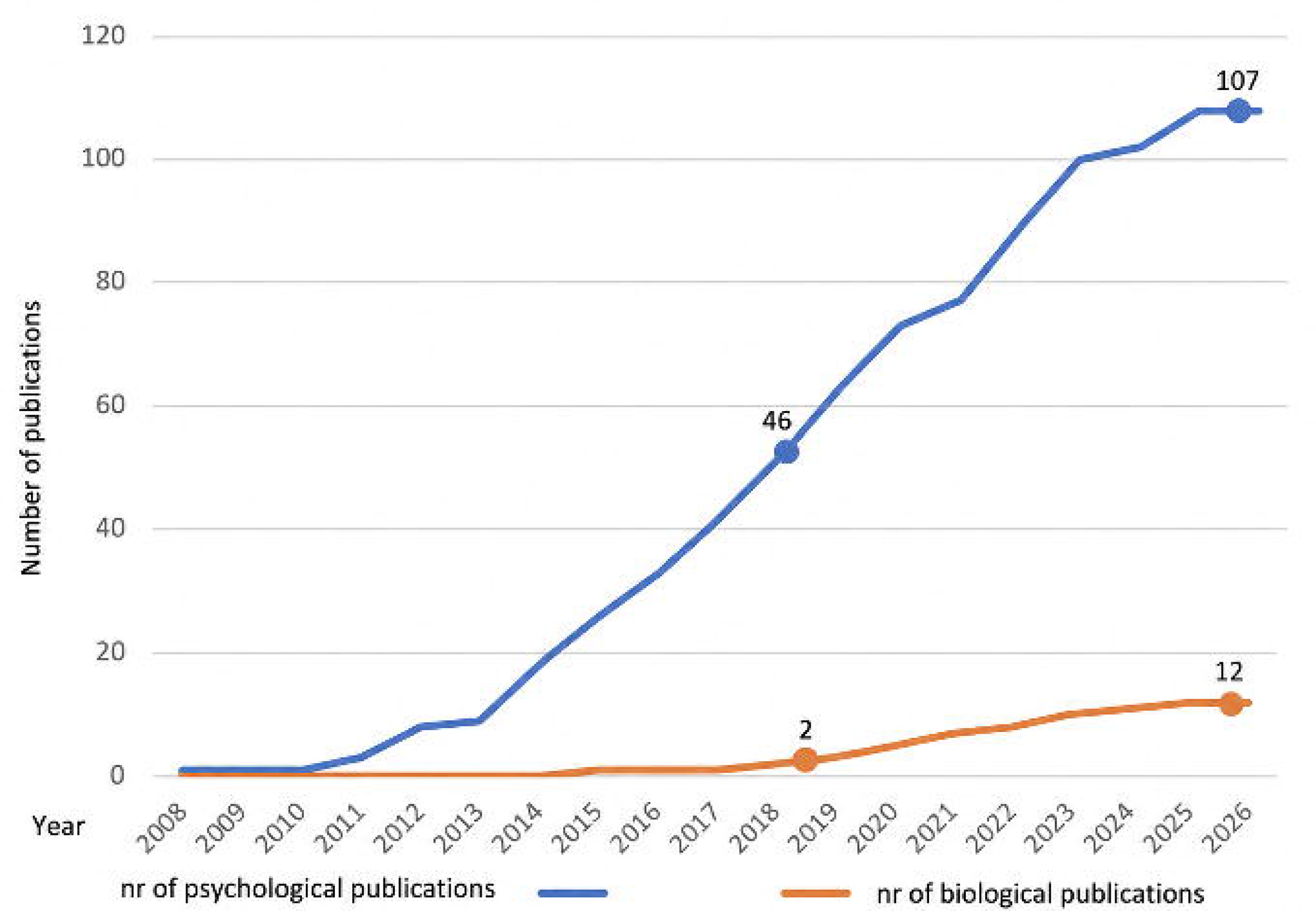

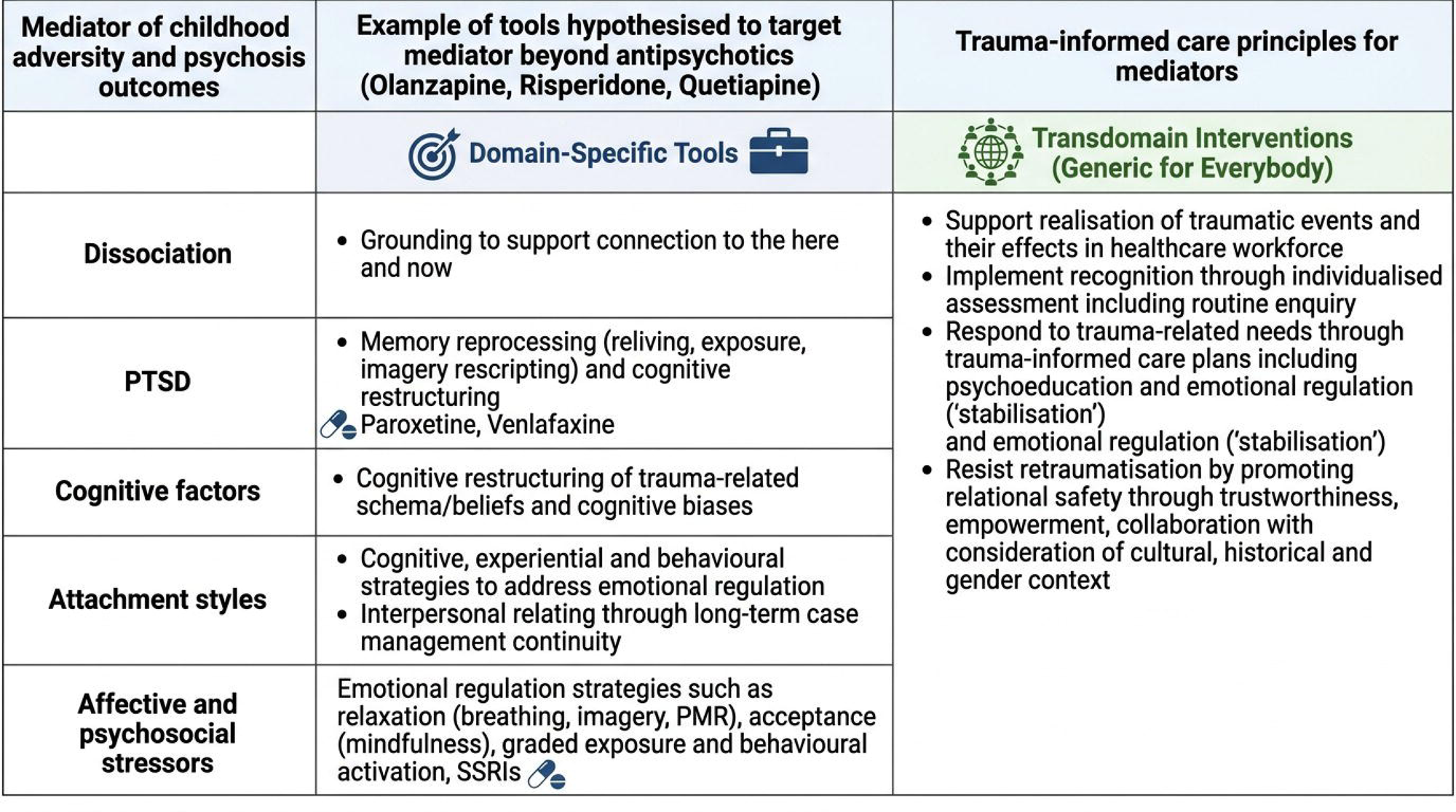

| Mediator of childhood adversity and psychosis outcomes | Example of tools hypothesised to target mediator beyond antipsychotics (Olanzapine, Risperidone, Quetiapine) | Trauma-informed care principles for mediators |
| --- | --- | --- |
|  | **Domain-Specific Tools** | **Transdomain Interventions (Generic for Everybody)** |
| **Dissociation** | Grounding to support connection to the here and now | Support realisation of traumatic events and their effects in healthcare workforce<br>Implement recognition through individualised assessment including routine enquiry<br>Respond to trauma-related needs through trauma-informed care plans including psychoeducation and emotional regulation ('stabilisation') and emotional regulation ('stabilisation')<br>Resist retraumatisation by promoting relational safety through trustworthiness, empowerment, collaboration with consideration of cultural, historical and gender context |
| **PTSD** | Memory reprocessing (reliving, exposure, imagery rescripting) and cognitive restructuring<br>Paroxetine, Venlafaxine |  |
| **Cognitive factors** | Cognitive restructuring of trauma-related schema/beliefs and cognitive biases |  |
| **Attachment styles** | Cognitive, experiential and behavioural strategies to address emotional regulation<br>Interpersonal relating through long-term case management continuity |  |
| **Affective and psychosocial stressors** | Emotional regulation strategies such as relaxation (breathing, imagery, PMR), acceptance (mindfulness), graded exposure and behavioural activation, SSRIs |  |

