## Supplementary Materials for "Exploring Psychological and Biological Mediators between Childhood Adversity and Psychosis: An updated Systematic Review and Meta-Analysis"

**Supplementary Online Content**

### Table S1. PRISMA Statement and Checklist

| **Section/Topic** | **#** | **Checklist Item** | **Page** |
| --- | --- | --- | --- |
| **TITLE** | | | |
| Title | 1 | Identify the report as a systematic review, meta-analysis or both. | 1 |
| **ABSTRACT** | | | |
| Structured Summary | 2 | Provide a structured summary including as applicable: background; objectives; data sources; study eligibility criteria, participants, and interventions; study appraisal and synthesis methods; results; limitations; conclusions and implications of key findings; systematic review and registration number. | 2-3 |
| **INTRODUCTION** | | | |
| Rationale | 3 | Describe the rationale for the review in the context of existing knowledge. | 4-5 |
| Objectives | 4 | Provide an explicit statement of objective(s) or question(s) the review addresses with reference to participants, interventions, comparison, outcomes, and study design (PICOS). | 5 |
| **METHODS** | | | |
| Eligibility Criteria | 5 | Specify the inclusion and exclusion criteria for the review and how studies were grouped for syntheses. | 6-7 & 8 |
| Information Sources | 6 | Specify all databases, registers, websites, organisations, reference lists and other sources searched or consulted to identify studies. Specify the date when each source was last searched or consulted. | 6 |
| Search Strategy | 7 | Present full electronic search strategy for at least one database, including any limits used, such that it could be repeated. | SM |
| Selection Process | 8 | State the process for selecting studies (i.e. screening, eligibility, included in systematic review, and if applicable, included in meta-analysis). | 6 - 8 |
| Data Collection Process | 9 | Specify the methods used to collect data from reports, including how many reviewers collected data from each report, whether they worked independently, any processes for obtaining or confirming data from study investigators, and if applicable, details of automation tools used in the process. | 7-8 & SM |
| Data Items | 10a | List and define all outcomes for which data were sought. Specify whether all results that were compatible with each outcome domain in each study were sought (e.g. for all measures, time points, analyses), and if not, the methods used to decide which results to collect. | SM |
|  | 10b | List and define all other variables for which data were sought (e.g. participant and intervention characteristics, funding sources). Describe any assumptions made about any missing or unclear information. | SM |
| Study Risk of Bias Assessment | 11 | Specify the methods used to assess risk of bias in the included studies, including details of the tools used, how many reviewers assessed each study and whether they worked independently, and if applicable, details of automation tools used in the process. | 8 |
| Effect Measures | 12 | Specify for each outcome the effect measures (e.g. risk ratio, mean difference) used in the synthesis or presentation of results. | 7-9 |
| Synthesis Methods | 13a | Describe the processes used to decide which studies were eligible for each synthesis, (e.g. tabulating the study intervention characteristics and comparing against the planned groups for synthesis). | 7-9 & SM |
|  | 13b | Describe any methods required to prepare the data for presentation or synthesis, such as handling of missing summary statistics, or data conversion. | 7-9 |
|  | 13c | Describe any methods used to tabulate or visually display results of individual studies and synthesis. | 7-9 |
|  | 13d | Describe any methods used to synthesise results and provide rationale for the choices. If meta-analysis was performed, describe the models, methods to identify the presence and extent of statistical heterogeneity, and software packages used. | 7-9 & SM |
|  | 13e | Describe any methods used to explore possible causes of heterogeneity among study results (e.g. subgroup analyses, meta-regression). | 9 |
|  | 13f | Describe any sensitivity analyses conducted to assess robustness of the synthesised results. | 9 |
| Reporting Bias Assessment | 14 | Describe any methods used to assess risk of bias due to missing results in a synthesis (arising from reporting biases). | 8 |
| Certainty Assessment | 15 | Describe any methods used to assess certainty (or confidence) in the body of evidence for an outcome. | 9 |
| **RESULTS** | | | |
| Study Selection | 16a | Describe the results of the search and selection process, from the number of records identified in the search to the number of studies included in the review, ideally using a flow diagram. | 10 & Flow chart |
|  | 16b | Cite studies that might appear to meet inclusion criteria, but which were excluded, and explain why they were excluded. | 10 |
| Study Characteristics | 17 | Cite each included study and present its characteristics. | 10-16 & SM |
| Risk of Bias In Studies | 18 | Present assessment of risk of bias for each included study. | 10-11 |
| Results in Individual Studies | 19 | For all outcomes, present, for each study: (a) summary statistics for each group (where appropriate) and (b) an effect estimates and its precision (e.g. confidence/credible intervals), ideally using structured tables and plots. | 10-12 |
| Results of Synthesis | 20a | For each synthesis, briefly summarise the characteristics and risk of bias among contributed studies. | 10-11 |
|  | 20b | Present results of all statistical syntheses conducted. If meta-analyses was done, present for each the summary estimate and its precision (e.g. confidence/credible interval) and measures of statistical heterogeneity. If comparing groups, describe the direction of the effect. | 11-12 |
|  | 20c | Present results of all investigations of possible causes of heterogeneity among study results. | SM |
|  | 20d | Present results of all sensitivity analyses conducted to assess the robustness of the synthesised results. | SM |
| Reporting Bias | 21 | Present assessments of risk of bias due to missing results (arising from reporting biases) for each synthesis assessed. | 10-11 |
| Certainty of Evidence | 22 | Present assessments of certainty (or confidence) in the body of evidence for each outcome assessed. | 10-11 |
| **DISCUSSION** | | | |
| Discussion | 23a | Provide a general interpretation of the results in the context of other evidence. | 17 |
|  | 23b | Discuss any limitations of the evidence included in the review. | 19-20 |
|  | 23c | Discuss any limitation of the review processes used. | 19-20 |
|  | 23d | Discuss implications of the results for practice, policy, and future research. | 20-22 |
| **OTHER INFORMATION** | | | |
| Registration and protocol | 24a | Describe sources of funding for the systematic review and other support; role of funders for the systematic review. | 23 |
|  | 24b | Indicate where the review protocol can be accessed, or state that a protocol was not prepared. | 5 |
|  | 24c | Describe and explain any amendments to information provided at registration or in the protocol. | SM |
| Support | 25 | Describe sources of financial or non-financial support for the review, and the role of the funders or sponsors in the review. | 23 |
| Competing Interests | 26 | Declare any competing interests of review authors. | 23 |
| Availability of Data, Code, and Other Materials | 27 | Report which of the following are publicly available and where they can be found: template data collection forms; data extracted from included studies; data used for all analyses; analytic code; any other materials used in the review. | 23 |

*Note.* Adapted from the PRISMA 2020 Statement (Page et al., 2021)^1^

### Table S2 Moose Checklist

| **Criteria** | | **Brief description of how the criteria were handled in the meta-analysis** |
| --- | --- | --- |
| **Reporting of background should include** | | |
| Ö | Problem definition | Childhood adversity (CA) has been identified as one of the most robust and potentially modifiable transdiagnostic risk factors for mental disorders. No studies have meta analysed the psychological mediators between CA and psychosis using meta-analytic structural equation modelling (METASEM). |
| Ö | Hypothesis statement | We hypothesised that biological and psychological factors mediate the association between childhood adversity (CA) and psychosis across clinical and general population samples. |
| Ö | Description of study outcomes | Assessed for psychological or biological mediators and with validated instruments between CA as detailed above and psychosis outcomes as dependent variable. Studies were required to report a mediation analysis using a robust method testing mediation, allowing the ascertainment of a mediating/indirect effect between the predictor and the outcome. |
| Ö | Type of exposure or intervention used | CA defined as exposure to psychologically threatening events involving abuse (sexual, physical or emotional), neglect (physical or emotional), domestic violence, parental mental illness or drug abuse, harsh parenting, and bullying. |
| Ö | Type of study designs used | Observational studies reporting mediation analysis of the association between childhood adversity and psychosis. |
| Ö | Study population | Individuals with psychosis diagnosis based upon validated diagnostic manuals and scales, which includes the Statistical Manual for Mental Disorders (DSM) and the International Classification of Diseases (ICD), or psychosis symptoms within the general population such as psychotic-like experiences. |
| **Reporting of search strategy should include** | | |
| Ö | Qualifications of searches | PsycINFO, Medline, and Embase |
| Ö | Search strategy, including time period included in the synthesis and keywords | An initial search was conducted via Ovid from January 2019 (date of Alameda et al. (2020) search) to June 2024. Subsequent updated search conducted in January 2026. Seach key words related to psychosis, CA, and mediation analyses. |
| Ö | Databases and registries searched | PsycINFO, Medline, and Embase |
| Ö | Use of hand searching | Included studies of relevant systematic reviews and the references from included studies were manually search and cross-referenced |
| Ö | List of citations located and those excluded, including justifications | Details of the literature search process are outlined in the results section and PRISMA flowchart. |
| Ö | Method of addressing articles published in languages other than English | Only articles in English were selected. |
| Ö | Method of handling abstracts and unpublished studies | Only original individual and published studies were selected. |
| Ö | Description of any contact with authors | Authors were contacted in the case of missing data or for further information via email. When no response was provided, there was a further attempt to contact. |
| **Reporting of methods should include** | | |
| Ö | Description of relevance or appropriateness of studies assembled for assessing the hypothesis to be tested | Details of inclusion and exclusion are provided in the methods section and Supplementary Materials. |
| Ö | Rationale for the selection and coding of data | Data extracted from each of the studies are relevant to study outcomes. |
| Ö | Assessment of confounding | A series of meta-regressions will be performed to examine the role of the percentage of females in the studies, FEP versus non-FEP samples, and (Newcastle Ottawa Scale) NOS scores |
| Ö | Assessment of study quality and stratification or regression on possible predictors of study results | Quality of included studies was evaluated by Newcastle Ottawa Scale (NOS). Meta-regression and sensitivity analyses were also performed to assess the influence of potential covariates. |
| Ö | Assessment of heterogeneity | Heterogeneity was assessed and other sources of heterogeneity was explored via meta-regressions and sensitivity analyses |
| Ö | Description of statistical methods in sufficient details to be replicated | Meta-analyses will be conducted in R using meta-analytic structural equation modelling (MASEM; Cheung et al., 2021), which synthesises multiple correlational relationships.  Heterogeneity will be assessed using the Q-test and I² statistic, with subgroup analyses by sample type conducted when at least ten studies per group are available.  Publication bias and small study effects will be assessed by examining the asymmetry in the funnel plots and with Egger's Regression. |
| Ö | Provision of appropriate tables and graphics | Tables highlighting meta analyses results have been included in the supplementary materials. |
| **Reporting of results should include** | | |
| Ö | Table summarizing individual study estimates and overall estimate | Included in manuscript narratively. Details in Supplementary Materials. |
| Ö | Table giving descriptive information for each study included | Included in Supplementary Materials |
| Ö | Results of sensitivity testing | Subgroup analyses are detailed in the results of manuscript as well as in the methods |
| Ö | Indication of statistical uncertainty of findings | Details of potential bias and limitations are discussed in the discussion. |
| **Reporting of discussion should include** | | |
| Ö | Quantitative assessment of bias | Bias was quantitatively assessed and reported. Details in Supplementary Materials. |
| Ö | Justification for exclusion | Exclusion criteria are detailed in the methods section. |
| Ö | Assessment of quality of included studies | Quality of studies was assessed and reported. Details of quality assessment of each study is reported in Supplementary Materials |
| **Reporting of conclusion should include** | | |
| Ö | Consideration of alternative explanations for observed results | Detailed in discussion section. |
| Ö | Generalization of the conclusions | Detailed in discussion section. |
| Ö | Guidelines for future research | Detailed in discussion section. |
| Ö | Disclosure of funding source | Followed by the conclusions. |

### Amendment of protocol

The literature search was updated in December 2025. At this stage, minor amendments were made to the original search strategy specified in the 2024 protocol. This updated version reflected methodological refinements and scope clarifications following the realisation that the magnitude of the data set after the 2026 update would allow us to conduct a meta-analysis, and not only a systematic review, as in the original submission. The updated protocol included a prespecified plan for a meta-analysis, which was the minor amendment.  The amended protocol also clarifies planned subgroup and meta-regression analyses. Some updates were also made to the review team and the timelines to reflect the revised project schedule. All amendments were implemented prior to the search and data synthesis, which will be conducted once this protocol is updated.

### Search Strategy

The present analyses did not consider separation from parents, abandonment and parental loss given the heterogenous definitions used to describe such type of adversities across studies.

**(EMBASE, Psyinfo and MEDLINE through Ovid provider, and Cochrane Libraries through Cochrane website)**

***Childhood adversity terms***

*108. sexual abuse.mp.*

*109. physical abuse.mp.*

*110. emotional abuse.mp.*

*114. psychological abuse.mp.*

*117. maltreat*.mp.*

*118. bully*.mp.*

*119. bullied.mp.*

*123. parental loss.mp.*

*124. (Separation adj5 parent).mp.*

*138. childhood trauma.mp.*

*139. early trauma.mp.*

*155. Neglect*.mp.*

*156. (trauma* adj5 experienc*).mp.*

*160. adversit*.mp.*

*161. (advers* adj5 experienc*).mp.*

*233. exp Child Abuse/*

*234. exp Physical Abuse/*

*235. exp Sexual Abuse/*

*236. exp Emotional Abuse/*

*237. exp Child Neglect/*

*238. exp Emotional Trauma/*

*239. exp BULLYING/*

*240. exp Parental Absence/*

*241. exp RAPE/*

*242. exp Domestic Violence/*

*243. exp Victimization/*

***Mediation terms***

*127. mediat*.mp.*

*130. (psycholog* adj3 mechanism*).mp.*

*131. (biolog* adj3 mechanism*).mp.*

*143. path analysis.mp*

*157. network analysis.mp.*

*170. structural equation.mp.*

*171. path analysis.mp*

*246. exp MEDIATION/*

*247. exp Structural Equation Modeling/*

*248. exp Path Analysis/*

***Psychosis terms***

*87. psychosis.mp.*

*88. psychot*.mp.*

*100. schizophr*.mp.*

*101. schizotyp*.mp.*

*102. hallucinat*.mp.*

*103. parano*.mp.*

*104. delusion*.mp.*

*105. persecut*.mp.*

*167. (disorganiz* adj5 symptom*).mp.*

*168. (disorganiz* adj5 dimension*).mp.*

*251. exp PSYCHOSIS/*

*252. exp SCHIZOPHRENIA/*

*253. exp CHILDHOOD SCHIZOPHRENIA/*

*254. exp SCHIZOTYPY/*

*255. exp HALLUCINATIONS/*

*256. exp PARANOIA/*

*257. exp DELUSIONS/*

*258. exp PERSECUTION/*

### Meta analysis collection and extraction procedures

This dataset will include the following variables: study DOI link; author name and year; sample type; percentage of female participants; mean age; NOS scores; trauma type grouped into composite categories (broadly measured), abuse, or neglect; trauma assessment instrument; mediator assessment instrument; mediator construct (e.g., for attachment, whether the measure reflects anxious or avoidant attachment); psychosis construct (composite, delusions, or hallucinations); and psychosis assessment instrument (e.g., Positive and Negative Syndrome Scale- PANSS). For the meta-analytical analyses, we will follow the framework proposed by Cheung (2022) to analyse mediating effects, as described in more detail below.

Three effect size measures were extracted: the correlation coefficient between the predictor and the mediator (path a), the correlation between the mediator and the outcome (path b), and the correlation between the predictor and the outcome (path c). These data were extracted from correlation matrices reported prior to the mediation analyses. When correlation matrices are not available, the required coefficients were extracted from preliminary analyses testing the associations corresponding to the paths described above. If these data were still unavailable, the relevant information was extracted from the reported mediation model paths. When effect sizes were not reported as correlations, they were converted to correlation coefficients using the Psychometrica conversion tool (https://www.psychometrica.de/effect_size.html). When data was missing and correlation could not be obtained, authors were contacted. Any effect sizes obtained through conversion, rather than direct extraction from correlation matrices, were recorded in the dataset for sensitivity analyses. When a mediator category included multiple measures within the same mediator family (e.g., a study reporting fearful and preoccupied attachment, both classified as anxious attachment conceptually), the corresponding correlation coefficients was combined using the methodology proposed by Olkin et al (1985), provided that the measures are judged to be clinically homogeneous (Olkin & Pratt, 1958). Decisions regarding clinical homogeneity, meaningful mediator categories, and the inclusion of specific subdomains in the meta-analyses will be discussed in group meetings led by NF and LA, and these were validated by a third senior author AH.

### Meta analysis procedures (operationalisation of predictors, mediators and outcomes).

For CA measures, when a study reports mediating pathways for more than one type of abuse or neglect, the subtype most frequently represented in the corresponding meta-analysis will be selected to minimise heterogeneity. For example, if most studies examine sexual abuse and one study examines emotional abuse, the predominant subtype (sexual abuse) will be selected. Sensitivity analyses will then be conducted with and without the least frequent subcategory (see below). When no single subtype is clearly more frequent than the others, the correlation coefficients for the relevant pathways will be combined. The same procedures will be applied to mediator categories and outcomes.

For mediating categories, based on our previous work^2^, we a priori expect to meta-analyse the following mediator families: dissociation, negative cognitive beliefs, anxiety, depression, and attachment. Drawing on the existing literature, a priori decisions have been established for these mediator families. For dissociation, studies examining composite dissociation, absorption, amnesia, and depersonalisation/derealisation will be meta-analysed separately. For depression and anxiety, overall scale scores will be meta-analysed. For attachment, anxious and avoidant attachment scores will be meta-analysed separately. Additional mediator families may be identified as new data emerge. Decisions regarding the creation of new mediator families and the selection of specific subdomains for meta-analysis will be made by consensus in group meetings led by NF, AH and LA.

With regard to the outcomes of interest, these will be grouped into: (a) composite or broad psychosis measures (e.g., case–control status, positive psychotic symptoms, psychosis-like experiences, or general schizotypy scores), and (b) specific symptom domains, namely delusions and hallucinations, which will be analysed separately. When two types of delusions are reported, the most frequent one withing the specific meta-analysis will be selected to limit heterogeneity (e.g if three studies report persecution and one study report persecution and grandiosity, persecution will be selected).

### Quality assessment procedures

Newcastle Ottawa Scale Selection

The quality assessment was carried out using the Newcastle–Ottawa Scale (see Quality Assessment Tool)^3^ for cohort studies by two independent reviewers (GK and IP). Those papers over which there was disagreement were discussed at a project group meeting. The Newcastle–Ottawa is a ten-point scale allocating points based on: the selection of cohorts (e.g. representativeness of the sample; 0–4 points), the comparability of cohorts (e.g. whether the study controls for confounding factors; 0–2 points), the identification of the exposure (e.g. objectivity of exposure measurement) and the outcomes of study participants (e.g. independence of outcome measurement, adequacy of follow-up; 0–3 points). Scores were considered as follows: “poor” quality for 3 or less; “fair” between 4 and 7 and “good” for scores of 8 or above. The agreed quality grades of each study are presented in Table S7, 8, 9 & 10. and the specific criteria used for our systematic review are specified in the Newcastle Ottawa Scale displayed below.

*Note: A study can be awarded a maximum of one star for each numbered item within the Selection and Exposure categories. A maximum of two stars can be given for Comparability.*

1. Representativeness of the exposed cohort
   1. truly representative of the average individuals with psychosis or attenuated psychotic symptoms in the community *
   2. somewhat representative of the average individuals with psychosis or attenuated psychotic symptoms in the community *
   3. selected group of users eg nurses, volunteers
   4. no description of the derivation of the cohort

0. Selection of the non exposed cohort
   1. drawn from the same community as the exposed cohort *
   2. drawn from a different source
   3. no description of the derivation of the non-exposed cohort

0. Ascertainment of exposure
   1. secure record*
   2. structured interview*
   3. written self-report* (star included here given the common use of self-reports in the field of adversity in psychosis)
   4. no description

0. Demonstration that outcome of interest was not present at start of study
   1. yes * (here we considered a star when the mediator was not present at the time of the assessment to traumatic experiences)
   2. no Comparability
0. Comparability of cohorts on the basis of the design or analysis

(in this section we considered none star if studies did not report the indirect and direct effects nor the percentage of total effect mediated. One start if they reported that information partially and two stars if they provided that information fully)*

0. Assessment of the outcome
   1. Assessment of outcome
      1. independent blind assessment *
      2. record linkage *
      3. self-report
      4. no description
0. Was follow-up long enough for outcomes to occur (prospective from birth)
   1. yes (if follow-up longer than 6 months) *
   2. no

0. Adequacy of follow up of cohorts (measures the attrition rate)
   1. complete follow up - all subjects accounted for *
   2. subjects lost to follow up unlikely to introduce bias - small number lost - > 20 % *
   3. follow up rate < 80%) and no description of those lost
   4. no statement

Table S3: Rationale for exclusion of mediation analyses from Figure 1.

| Study (Author, Year) | Mediator | Reason % Mediated not reported in Figure 1 |
| --- | --- | --- |
| Khosravi et al. (2021, Iran)^4^ | Dissociation | *SA -> a -> 1*  *(DE = 0.32*; IE = -0.23)*  *% = -82.14*  The indirect effect was in the opposite direction to the total effect (inconsistent mediation). This analysis was therefore not included in the descriptive summary presented in Figure 1. |
| Nesbit et al. (NZ, Australia, NI, 2022)^5^ | Dissociation | To maintain consistency between findings included in Figure 1, results looking at dissociative identity disorder were not included.  From the schizophrenia spectrum disorders the below pathway wasn’t included because it looked at olfactory hallucinations.  *CA -> c -> 4*  *(DE = 0.126*; IE= 0.129)*  *% = 57.3* |
| Bellido-Zanin et al.  (2018, Spain)^6^ | Dissociation | The following analysis was not included because it was reporting a serial mediation.  *ELES -> A.1 + A.2 + A.3 = 1*  *Total Mediation*  *(DE = 0.06; IE A.2 = 0.05*; IE A.3 = 0.04*)*  *% A.2 = 33.3*  *% A.3 = 27* |
| Berenbaum et al. (2008)^7^ | Dissociation & PTSD | Insufficient information was reported to calculate the percentage of the total effect mediated (only reported indirect effect  for men and woman) |
| Bortolon et al. (2017, France)^8^ | Dissociation & Negative schemas | Insufficient information was reported to calculate the percentage of the total effect mediated |
| Levi et al. (2025, UK)^9^ | Dissociation, negative schemas, attachment | Network analysis meant that % mediated  couldn’t be calculated. |
| Mertens et al.  (2021, Spain)^10^ | Dissociation & Attachment | The study reported  parallel mediation only; individual indirect effects comparable with the descriptive summary could not be extracted. |
| LoPilato et al., (2020, USA, Canada)^11^ | Negative Schemas | Analyses of other psychosis outcomes (e.g., grandiosity) were excluded to maintain consistency in the descriptive synthesis. |
| Ashford et al. (2012, UK)^12^ | Negative Schemas, Anxiety & Depression | Insufficient information was reported to calculate the percentage of the total effect mediated |
| Boyda et al. (2018, UK)^13^ | Negative Schemas | Insufficient information was reported to calculate the percentage of the total effect mediated |
| Fisher et al. (2013, UK)^14^ | Negative schemas, depression & anxiety | Multiple mediation looking at negative schemas, depression and anxiety collectively. The study reported serial mediation only; individual indirect effects comparable with the descriptive summary could not be extracted |
| Jaya et al.(2016, Germany)^15^ | Negative Schema | Insufficient information was reported to calculate the percentage of the total effect mediated |
| Jin et al.  (2022, China)^16^ | Negative Schema & Depression | Network analysis meant that % mediated  couldn’t be calculated. |
| Strelchuk et al.  (2022, UK)^17^ | PTSD | This study included two mediation analyses which had different CA age ranges. To avoid double-counting findings from the same sample, only one analysis was included in Figure 1  *CT Age 0-17 -> 2 -> B*  *(DE = 1.51*; IE = 1.03*)*  *% = 8* |
| Huang et al. (China, 2023)^18^ | Depression | Network analysis meant that % mediated  couldn’t be calculated. |
| Metel et al. (2020, Poland)^19^ | Depression | The study reported serial or parallel mediation only; individual indirect effects comparable with the descriptive summary could not be extracted.  *TEC/CECA-Q -> DACOBS ->  CD-RISC  CESD-R->   PLE* |
| Moffa et al.  (2017, UK)^20^ | Depression | Insufficient information was reported to calculate the percentage of the total effect mediated |
| Qiao et al.  (2023, Belgium)^21^ | Attachment | Network analysis meant that % mediated couldn’t be calculated. |
| Sheinbaum et al.  (2015, Spain)^22^ | Attachment | The study reported parallel mediation only; individual indirect effects comparable with the descriptive summary could not be extracted |
| Sheinbaum  (2020, Spain)^23^ | Attachment | The study reported parallel mediation only; individual indirect effects comparable with the descriptive summary could not be extracted |
| Goldstone et al.  (2012, Australia)^24^ | Mentalisation | The study reported path analysis only; individual indirect effects comparable with the descriptive summary could not be extracted |
| Nonweiler et al.  (2023, Spain)^25^ | Mentalisation | In total this study included 90 analyses; only composite scores were included in the figure to avoid too many from the same simple. |

Non significant analyses were not included.

### Table S4. Included Studies Categorised by Clinical Sample Characteristics

| Early Psychosis/ FEP | (Alameda et al., 2022^26^, 2023^27^; Evans et al., 2015^28^; Morgan et al., 2014^29^; Peach et al., 2019^30^; Setién-Suero et al., 2022^31^; Sun et al., 2018^32,33^; Wang et al., 2022^33^, 2023^34^; Xie et al., 2022)^35^ |
| --- | --- |
| UHR | (Appiah-Kusi et al., 2017^36^; LoPilato et al., 2021^11^; McDonnell et al., 2018^37^; Pan et al., 2019^38^; Rapado-Castro et al., 2020^39^; Thompson et al., 2016^40^) |
| Non- FEP | (Adewuya et al., 2025^41^; Barnes et al., 2023^42^; Baryshnikov et al., 2018^43^; Bhui et al., 2021^44^; Cancel et al., 2015^45^; Chatziioannidis et al., 2019^46^; Chiu et al., 2024^47^; Choi et al., 2015^48^; Corcoran et al., 2020^49^; Dudley et al., 2023^50^; Fuchshuber et al., 2025^51^; Goldstone et al., 2011^52^, 2012^24^; Hardy et al., 2016^53^; Humphrey et al., 2022^54^; Isvoranu et al., 2017^55^; Laskemoen et al., 2021^56^; León-Palacios et al., 2020^57^; Levi et al., 2025^9^; Mansueto et al., 2019^58^; Nesbit et al., 2022^5^; Neumann et al., 2023^59^; Østefjells et al., 2017^60^; Pearce et al., 2017^61^; Perona‐Garcelán et al., 2012^62^; Pilton et al., 2016^63^; Powers et al., 2011^64^; Quidé et al., 2018^65^; Rosen et al., 2018^66^; Schalinski et al., 2019^67^; Sengutta et al., 2019^68^; Seo & Choi, 2018^69^; Simpson et al., 2020^70^; Steenkamp et al., 2022^71^; Styła et al., 2019^72^; Uyan et al., 2022^73^; Van Dam et al., 2014^74^; Varese et al., 2012^75^; Weijers et al., 2018^76^; Wickham & Bentall, 2016^77^) |
| Early psychosis and non FEP | (Betz et al., 2020^78^; Khosravi et al., 2021^4^) |

### Table S5: Overview of Identified Psychological Mediators and Corresponding Subitems

| **Psychological Mediators Identified** | **Mediator Subitems** |
| --- | --- |
| Dissociation | Composite, Amnesia, Depersonalization/derealization, Absorption, Somatoform Dissociative Symptoms |
| Negative Schema | Negative schemas, Self-esteem, Self-concept, Shame, Self-disgust, Maladaptive schemas, Schematic Beliefs, Negative core beliefs (self- others), Negative social comparisons |
| PTSD | Composite, Avoidance, numbing, Hyperarousal, Intrusive trauma memory, Post traumatic intrusions, Trauma related beliefs |
| Depression | Composite |
| Anxiety | Composite, Worry |
| Attachment Style | Composite, Avoidant, Anxious, Disorganised, Preoccupied, Dismissing, Enmeshed, Fearful, Angry-dismissive, Withdrawn, Insecure, |
| Mentalisation | Self-Mentalizing, Other Mentalizing, Metacognitive beliefs, metacognition, Mentalizing abilities |
| Psychosocial Stress | Current Adversity, Adverse life events, Perceived Stress, Recent stress-events, Life Hassles, Stress |
| Affective Dysregulation | Composite, Emotion regulation, Mood instability |
| Cognitive Biases | Includes attributional style (LoC) and interpretation bias |
| Loneliness | Composite |
| Sleep disturbance and fatigue | Sleep disturbances, insomnia and fatigue |
| Anomalous experimental processes | Aberrant salience, Anomalous experiences, Negative voice content and Persecutory ideation |
| Personality features | Borderline personality features, Personality Organisation, Personality |
| Positive attributes | Resilience**,** Mindfulness, Wisdom, Positive attributes, Motivation and Pleasure |
| Other psychosocial | Common mental health disorders, Interpersonal sensitivity, Cognitive functioning Psychosocial functioning , Belief Updating, Parent-Child Conflict, Feeling safety, Bipolar Traits, Social Rank, OCD symptoms, Intelligence, Threat Anticipation, Negative Affect, General Psychopathology, Social Defeat, Time Perspective, Mood Swings, Perception of Injustice, IQ |

### Table S6: Studies including multiple analyses which were excluded from quantitative analyses reports

| Betz et al., 2020^78^ | Network analysis; discrete mediation analyses could not be identified. |
| --- | --- |
| Goldstone et al., 2011^52^ | It is a **path analysis/structural equation model** with multiple sequential pathways |
| Goldstone et al., 2012^24^ | It is a **path analysis/structural equation model** with multiple sequential pathways |
| Huang et al., 2023^18^ | Network analysis; discrete mediation analyses could not be identified. |
| Isvoranu et al., 2016^55^ | Network analysis; discrete mediation analyses could not be identified. |
| Jin et al., 2022^16^ | Network analysis; discrete mediation analyses could not be identified. |
| Levi et al., 2025^9^ | Network analysis; discrete mediation analyses could not be identified. |
| Qiao et al., 2024^21^ | Network analysis; discrete mediation analyses could not be identified. |

### Table S7. Overview of Psychological Mediators in clinical studies included in this review

| **Authors**  **Country** | **Sample**  **Mean Age**  **% female** | **Design** | **Measures of childhood adversity** | **Mediator(s)** | **Analysis** | **Bootstrap**  **(yes / no) / confounders (yes / no)** | **Psychosis** | **Main findings**  **Pathway**  **Total / partial mediation**  **Direct Effect (DE)**  **Indirect effect (IE)**  **% total effect mediated** | **Quality Score** |
| --- | --- | --- | --- | --- | --- | --- | --- | --- | --- |
| **Alameda et al. (Switzerland, UK, 2022) ^26^** | 330 Early Psychosis  24.75  34.2 % | Prospective | Composite  Interview | (A) Depression (MADRS) (B) Anxiousness (PANSS) (C) Both | Mediation analysis (multi-level) | No  Yes (age, sex, DUP, bullying, PN, EN) | 1. Positive symptoms early psychosis   (PANNS) | **CT -> a -> 1**  **2 months**  (DE = 1.313; IE = .423)  % = 24  **36 months**  **(**DE = 1.564; IE = .447)  % = 22  **3Y Follow Up** (DE = 1.075; IE = .392)  % = 27  **CT -> b -> 1**  **2 months**  (DE = 1.173; IE = .575)  % = 33  **36 months**  (DE = 2.058, IE = .185)  % = 8  **3Y Follow up**  (DE = 1.155; IE = .421)  % = 27  **CT -> c -> 1**  **2 months**  (DE = 1.294; IE = .546)  % = 30  **36 months**  (DE = 1.963; IE = .163)  % = 8  **All the period:**  **(**DE = 1.118; IE = .475)  % = 30 | 6 |
| **Adewuya et al. (2025, Nigeria) ^41^** | 1953  945 PLEs  95 psychosis  1008 controls  34.7  % ns | Cross sectional | Composite & subtypes (composite abuse, composite neglect)  CTQ-sf | A. Common mental disorders (Depression (PHQ-9), anxiety (GAD-7), somatic symptoms)  B. resilience (RS-14)  C. self esteem (RSEQ)  D. Cognitive functioning (MMSE) | Generalized structural equation modeling (GSEM) | Yes  confounders : age, gender, socioeconomic position | 1.. Psychosis (MINI 7) | CT → A → 1  (DE = 0.105; IE = 0.022)  % = 20.9  CT → B → 2  (DE = 0.105; IE = 0.024)  % = 22.9  CT → C → 1  Non significant  CT → D → 1  (DE = 0.225; IE = 0.041)  % = 18.1 | 6 |
| **Appiah-Kusi *et al.* (2017, UK)^36^** | 30 UHR; 38 HC  23.9  46.7% | Cross sectional    Case control study | CTQ  EA | (1) Schematic beliefs (BCSS)  1.a - Negative Self-schemas (NSS) | Regression based approach | Yes    Yes (Cannabis use, depression, anxiety) | (1) UHR caseness    (2) Paranoia  (PTS) | **EN -> BCSS-> UHR**  Partial mediation (DE = 0.261*; IE = 0.045*)  % = 14.7    **EN -> BCSS -> PSQ**  Partial mediation (DE = 1.353*; IE = 0.988*)  % = 42.2    Other adversities were not related to outcomes so were not included for mediation analyses | 6 |
| **Barnes et al. (2023, UK)^42^** | 171  Clinical sample (SSD)  42.2  38% | Cross sectional | Subscales (EA/N, PA, SA, multiple abuse) | a. Anxiety (BAI) | SEM | No  Yes (Age, Gender, Ethnicity, Socioeconomic status Marital status) | 1. Hallucinations   1.A auditory hallucinations  1.B Multimodal hallucinations   2. Delusions (SAPS)   2.A Persecutory delusions  2.B Grandiose/religious  2.C Delusions of influence | **EA & N -> a -> 2.c**  (DE = 0.96; IE = 1.24*)  % = 56  **Multiple Abuse -> a -> 2.A**  (DE = 0.17; IE = 0.23*)  % = 58  **Multiple abuse -> a -> 2.c**  (DE = 1.86; IE = 1.11*)  % = 37  All other mediators were non-significant. | 6 |
| **Baryshnikov et al.**  **(2018, Finland)^43^** | 282  Mood Disorder  42.2  74.1% | Cross sectional | Composite  TADS | (A) BPD symptoms (MSI) | Mediation analysis | Yes  Yes (age) | (1) Positive symptoms (CAPE) | **TADS -> A -> 1**  (DE = 0.03; IE = 0.11*)  % = 78 | 6 |
| **Betz et al. (2020, Finland, Germany, Italy, Switzerland, UK)^78^** | 547  (265 CHR, 282 ROP)  24.7  47.5% | cross-sectional study with additional longitudinal modeling | Subtypes (EN, PN, EN, PA, SA)  CTQ-SF | a. General psychopathology | Network analysis | Yes  No | 1. Positive symptoms (PANSS) | Types of CA showed several independent pathways to psychotic symptoms via general psychopathology (e.g. EA via depression and anxiety to suspiciousness). | 4 |
| **Bhui et al. (2021, UK)^44^** | 134 456  55.72  54.6%  480 (psychosis) | Prospective cohort study | Composite (past adversity)  ACE self-report | a. Current adversity | PLS--SEM | Yes  No | 1. Caseness (ICD-10) | **CT -> a -> 1**  (DE = 0.028; IE = 0.007)  % = 20 | 5 |
| **Chatziioannidis *et al.* (2019, Switzerland)^46^** | 63 SSP; 61 HC  40.4 SSP  30.16% SSP  39.33 HC  29.5% HC | Cross sectional    Case control study | Composite    CECA.Q | (1) Attachment (ECR-R)  1.a - Avoidance  1.b - Anxiety | Parallel Multiple mediation model | Yes    Yes (education) | (A) Caseness (SSP)  (PANSS) | **CT -> 1.b -> SSP**  Partial mediation (DE = 1.70*; IE = 1.24*)  % = 41.9    No significant mediation of attachment avoidance between CT and SSP | 6 |
| **Chiu et al. (2024, Taiwan)^47^** | 40 HC  32.7  0%  43 (SCZ)  39.6  2.3% | Cross sectional | Composite  CT (BBTS) | a. Dissociative symptoms (CADSS) & (DSS) | Conditional process analysis | Yes  Yes (DD, SUD) | Psychotic Symptoms - Caseness   1. Clinician Rated   (CADSS & PANSS-P)  a. SCZ   2. Patient Ratings (DSS & LSHS)   a. SCZ   3. Patient Ratings   (DSS & PDI)  a. SCZ | **Clinician Rating**  **CT -> a -> 1.a**  **N.S**  **CT -> a -> 2.a**  **N.S**  **CT -> a -> 3.A**  **N.S** | 5 |
| **Choi Ji *et al.* (2015, Republic of Korea)^48^** | 126 psychosis  36.1  55.6% | Cross sectional | Composite abuse (CA)    Korean CTQ | (1) Posttraumatic stress symptoms (IESR) | SEM | No    No | (1) Psychotic symptoms (PS)  PSYCH subscale of the PSY-5 factor scale of MMPI-2 | **CA-> IESR-> PS**  Partial mediation (DE = 0.30*; IE = 0.171*)  % = 36.3 | 4 |
| **Dudley et al. (2023, UK)^50^** | 292  Psychosis  42.31  28 % | Cross sectional | Composite & Subtype  (Abuse)  CTQ | Schematic beliefs (BCSS)   (A) Negative self schemas (NSS) (B) Negative other (NSO) | Mediation analysis | Yes  No | (1) Psychosis symptoms (PANSS) (2) Hallucinations   (PSYRATS) | **CT -> NSS -> 1**  (DE = 0.12*; IE = 0.033*)  % = 18.3  **CT -> NSO -> 1**  (DE = 0.12*; IE = 0.0279*)  % = 15.5  **CA -> NSS -> 2**  N.S  **CA -> NSO -> 2**  N.S | 6 |
| **Evans *et al.* (2015, UK)^28^** | 29 EP; 31 HC  18-38 EP  34.5% EP  NA HC  38.7% HC | Cross sectional    Case control study | Composite and subscales (SA, PA, EA, PN, EN)    CTQ | (1) The Self-Concept Clarity Scale (SCCS) | Mediation analysis | Yes    No | (1) Caseness  (PANSS) | **EN -> SCCS -> 1**  Total mediation (DE = 0.033; IE = 0.157*)  % = 82.8    None of the other total effects were significant so no mediation was possible    The remaining pathways between EN and caseness through SCCS and DES were not significant | 5 |
| **Fuchsuber et al. (2025, Austria)^51^** | 119  (76% clinical, other controls)  range 19–60 years  %71.4 | Cross sectional | Composite  CTQ | Personality Organisation (PO)  STIPO | Bayesian path modeling (mediation) | No  Age, sex | Paranoid thinking  BSI-53 | **CT -> PO -> paranoid thinking**  **(DE = 0.38*; IE = 0.14)**  **% ≈ 26** | 6 |
| **Goldstone et al.**  **(2011, Australia)^52^** | 100 psychosis  NA  44%  133 Non Clinical  NA  59% | Cross sectional (Emotional Trauma and Sexual Trauma) | Subtypes (Emotional Trauma and Sexual Trauma)  ETI | (A) Life hassles (SRLEs) | Path analysis | No  No | (1) Delusions   (PDI) | **Clinical**  **SA -> A -> 1**  (DE = NA; IE = 0.03*)  % = NA | 5 |
| **Goldstone et al.**  **(2012, Australia)^24^** | 100 Psychosis  NA  44%  133 Non Clinical  NA  59% | Cross sectional | Subtypes (Emotional Trauma and Sexual Trauma)  ETI | (A) Life hassles (SRLEs) (B) Metacognitive beliefs (MCQ) | Path Analysis | No  No | A.1  Hallucinations (LSHS-R)  A.2 Auditory Hallucinations  (LSHS-R) | **Clinical**  **(EA -> A -> A.1)**  (DE = NA; IE = 0.03*)  % = NA  **Clinical**  **(EA -> A -> A.2)**  (DE = NA; IE = 0.05*)  % = NA | 5 |
| **Hardy *et al.* (2016, UK)^53^** | 228 psychosis  38.2  27.6% | Cross sectional | Childhood sexual abuse (SA), Childhood emotional abuse (EA)    THQ | (1) PTSD symptoms  (SRS-PTSD)  1.a - Avoidance & Numbing  1.b - Hyperarousal  1.c - Intrusive trauma memory    (2) Cognitive bias / schemas  (BCSS)  2.a - Negative others beliefs | Mediation analysis | No    Yes (age, gender and ethnicity | (A) Positive symptoms  A.1 - Auditory hallucinations  A.2 - Persecutory hallucinations  A.3 - Ideas of reference | **SA-> 1.a -> A.1**  Total mediation (DE = 2.052; IE = 1.475*)  % = 48.74    **SA -> 1.b -> A.1**  Total mediation (DE = 2.104; IE = 1.439*)  % = 47.5    **EA-> 2.a-> A.2**  Total mediation (DE = 1.889; IE = 1.359*)  % = 52.9    Non-significant mediating effects of CSA with A1 through 2.a; CEA with A.3 through 2.a and CSA with A.1 through 1.c | 5 |
| **Humphrey et al. (UK, 2022)^79^** | 242 (clinical sample)  33.17  30.6 | Cross sectional | Composite  BBTS | 1. Disorganized attachment (PAM-R) | Path analysis | Yes  Yes (age, depression, hallucinations) | A. Paranoia (PaDS) | **BBTS -> 1 -> A**  (DE = .07*; IE = .063)  % = 47 | 5 |
| **Isvoranu *et al.* (2017, Netherlands)^80^** | 552 psychosis  30.76  25% | Cross sectional | SA, PA, EA, PN, EN    CTQ | (1) 18 items from general psychopathology (PANSS) | Network based analysis | No    Yes (all PANSS items) | (1) 6 items from positive dimension (PANSS) | Suggested mediation in pathways:  **1) EA -> Anxiety -> paranoia / suspiciousness**  **2) PA-> impulse control=> grandiosity / excitement / hostility**  % = N/A | 4 |
| **Khosravi et al. (2021, Iran)^4^** | 210  140 clinical 70 HC  NA  31.1% | Cross sectional | Subtypes (SA)  CTQ-SF | a. Amnesia  b. Depersonalization/derealization  c. Absorption | Mediation analysis | Yes  Yes (gender, comorbid disorders (borderline, DID), age, education level) | 1. Positive psychotic symptoms - Caseness  (PANSS) | **SA -> a -> 1**  (DE = 0.32*; IE = -0.23)  % = -82.14  **SA -> c -> 1**  (DE = 0.32*; IE = 0.24)  % = 86 | 5 |
| **Laskemoen et al. (2021, Norway)^56^** | 766 (SSD & BD)  30.9  49.9% | Cross sectional | Composite  CTQ | a. Insomnia   (IDS-C) | Mediation analysis  Path analysis | Yes  Yes (age, sexe, socio-economic status) | 1. Psychosis severity (PANSS) | **CT -> a -> 1**  (DE = 0.7*; IE = 0.06)  % = 25 | 6 |
| **Leon-Palacios**  **(2020, Spain)^57^** | 437  295 (HC)  142 (patients)  34.21  77.6 | Cross Sectional | Emotional Memories  Subtypes  ELES  (Threat, submission, devaluation) | (1) Fatigue  (CFQ)  (2) Aberrant Salience | Mediation Analysis | Yes  Yes (age) | (A) Ideas of Reference - caseness  (REF) | **Threat -> 1 -> REF**  (DE = 0.18*, IE = 0.03*)  % = 8.3  **Threat -> 2 -> REF**  (DE = 0.18*, IE = 0.16*)  % = 44.4  **Submission -> 1 -> REF**  (DE = 0.17*, IE = 0.03*)  % = 7.9  **Submission -> 2 -> REF**  (DE = 0.17*, IE = 0.18*)  % = 47.4  **Devaluation -> 1 -> REF**  (DE = 0.19*, IE = 0.07*)  % = 12.5  **Devaluation -> 2 -> REF**  (DE = 0.19*, IE = 0.30*)  % = 53.6 | 5 |
| **Levi et al. (2025, UK)^9^** | 609 (259 psychosis, 350 GP)  28.70  % 25 | Cross sectional | Subtypes (PA, SA, EA)  BBTS | A.insecure attachment (PAM-R)  B.dissociation (DES)  C.negative schemas (BCSS) | Network analysis | Yes  No | Psychotic experiences (CAPE-42) | **Network analysis**  Childhood trauma variables in the psychosis diagnosis network formed as an isolated cluster. | 4 |
| **LoPilato et al., (2021, USA, Canada)^11^** | 531 CHR  18.80  39.5% | Prospective | Composite & Subscales  (Threat & Deprivation) | a. Negative-self (BCSS) b. Negative-other schemas (BCSS) | Mediation analysis | Yes  Yes (depressive symptoms, sex) | 1. Prodromal Syndromes (SIPS)    1.1 delusional thinking    1.2 Suspiciousness    1.3 Grandiosity    1.4 Perceptual abnormalities | **T-> b- > 1.1**  (DE = 0.08; IE = 0.02)  % = 20  **T -> b -> 1.2**  (DE = 0.03; IE = 0.04)  % = 56  **CA -> b -> 1.1**  (DE = 0.02; IE = 0.02)  % = 50  **CA -> b -> 1.2**  (DE = 0.11; IE = 0.05)  % = 31  All other indirect effects were non-significant. | 6 |
| **Mansueto et al. (2019, Belgium)^58^** | 1119 schizophrenia  27.66  25% | Prospective | Subtypes (Abuse) | a. Mentalizing abilities | Mediation analysis | Yes  Yes (age, sexe, cannabis consumption) | 1. Positive symptoms   (PANNS) | Null Mediation | 5 |
| **McDonnell *et al.* (2018, UK)^37^** | 64 UHR  22.5  40.6% | Cross  sectional | Bullying severity in childhood / adolescence (BS)    RBQ | (1) Interpersonal sensitivity (IS) (IPSM) | Path analysis | Yes    No | (1) Paranoid ideation (PI) (SSPS) | **BS (Childhood)-> IS -> PI**  Total mediation (DE = 0.131; IE = 0.129*)  % = 49.6    Bullying in adolescence was not significantly associated with paranoid ideation | 5 |
| **Morgan *et al.* (2014, UK)^29^** | 390 FEP; 391 HC  30.5  44.1% | Cross sectional    Case control study | Parental separation / death    MRC | (a) Self-Esteem (RSES) | Multiple mediation analyses | Yes    Yes (age, gender, ethnicity, study centre, parental history of psychosis and IQ) | (1) FEP caseness based on ICD-10 | Null mediation | 6 |
| **Nesbit et al. (NZ, Australia, NI, 2022)^5^** | 99 (50 DID, 49 SSD)  DID: 45.14  SSD: 43.76  DID: 96%  SSD: 42.9% | Cross sectional | Composite  CTQ | Dissociation (DES-II)   a. Depersonalization b. Amnesia c. Absorption | Mediation analysis | Yes  No | 1. AH Frequency (Single question item) 2. AH Distress (PSYRATS distress) 3. Visual Hallucination Frequency (MUPS Visual) 4. Olfactory Hallucination Frequency (MUPS Olfactory) | **Group DID**  **CA -> a -> 2**  (DE = -0.192*; IE = 0.274)  % = 33  **CA -> a -> 3**  (DE = 0.47*; IE = 0.234)  % = 83.2  **CA -> b -> 3**  (DE = 0.062*; IE = 0.220)  % = 26.1  **CA -> a -> 4**  (DE = 0.206*; IE = 0.234)  % = 58.1  **CA -> b -> 4**  (DE = 0.185*; IE = 0.255)  % = 58  **Group SSD**  **CA -> c -> 3**  (DE = 0.127*; IE = 0.136)  % = 51.7  **CA -> c -> 4**  (DE = 0.126*; IE= 0.129)  % = 57.3 | 5 |
| **Neumann et al. (2023,**  **Germany)^59^** | 253  Psychosis  39.1  NA | Cross sectional | Composite CTQ | a. Psychosocial functioning (PSP) | Mediation Analysis  ordinary least-squares regression | Yes  No | 1. Positive symptoms (PANSS) | **CT -> a -> 1**  (DE = 0.03; IE = 0.03)  % = 60 | 6 |
| **Ostefjells et al.**  **(2017, Norway)^60^** | 261  30.2  46.4% | Cross sectional | Subtype (EA) | (A) Metacognition (MCQ) (B) Depression and anxiety symptoms (PANSS) | Serial mediation | Yes  Yes (sex, diagnosis, duration treatment) | (1) Positive psychotic symptoms (PANSS) | EA -> A -> B -> 1  (DE = 0.36*; IE = 0.05*)  % = 12  EA -> A -> 1  N.S  EA -> B -> 1  N.S | 6 |
| **Pan et al.**  **(2019, Brazil)^38^** | 2511  13.45  46.1% | Longitudinal | Composite  (CTQ) | Positive attributes (DAWBA) | Mediation Analysis | Yes    Yes (age, study site, sex, SES) | Psychotic Experiences  (CAPE) | **CTQ -> DAWBA -> PEs**  (at 3 yr follow up)  (DE = 0.1086* ; IE = 0.0166*)  % = 14 | 8 |
| **Peach *et al.* (2019, Australia)^30^** | 66 FEP  20.18  54.5% | Cross sectional | Composite    CTQ | (1) Post traumatic intrusions (CAPS)    (2) Trauma related beliefs (PTCI) | Simple mediation analyses | Yes    No | (A) Hallucinations (PANSS)    (B) Delusions  (PANSS) | **CT -> CAPS -> A**  Total mediation (DE = 0.01; IE = 0.01*)  % = 50    **CT -> CAPS -> B**  Total mediation (DE = 0.01; IE = 0.02*)  % = 66.6    **CT -> PTCI -> A**  Total mediation (DE = 0.01; IE = 0.01*)  % = 50    **CT -> PTCI -> B**  Total mediation (DE = 0.01; IE = 0.02*)  % = 66.6 | 5 |
| **Pearce et al.**  **(2017, UK)^61^** | 112  40.26  72 | Cross sectional | Composite  BBTS | (A) Dissociation (DES-R) (B) Attachment | Parallel Mediation | Yes  Yes (Comorbidity) | (1) CAPE   1.1 Voices  1.2 Paranoia | **BBTS -> A + B -> 1.1**  (DE = 0.01; IE A = 0.09*; IE B = N.S)  % A = 90  **BBTS -> A + B -> 1.2**  (DE = -0.05; IE = 0.23*; IE A = 0.16*; IE B = 0.05*)  % = 82  % A = 57  % B= 18 | 6 |
| **Perona‐Garcelán *et al.* (2012, Spain)^62^** | 71 psychosis  39.1  31.5% | Cross sectional | Composite    TQ | (1) Dissociation (DES-II)    1.a - Dissociative  Amnesia (DAM)    1.b - Absorption and Imaginative Involvement  (ABI)    1.c – Depersonalization / Derealization (DP) | Simple and multiple mediation analysis | Yes    No | (1) Hallucinations (PANSS)    (2) Delusion  (PANSS) | **TQ -> DES -> hallucinations**  Total mediation (DE = 0.20; IE = 0.21*)  % = 51.2    **TQ -> DP-> hallucinations**  Total mediation (DE = 0.20; IE = 0.19*)  % = 46.3    No mediating effects on delusions or mediating effects of ABI, DAM on hallucinations | 5 |
| **Pilton et al.**  **(2016, UK)^63^** | 55  46.16  20% | Cross Sectional | Subscales (SA, EA, PN)  CTQ | (1) Attachment (PAM)   1.a Insecure anxious  1.b Insecure avoidant | Mediation Analysis | Yes  No | (A) Hallucinations (PSYRATS) | **SA -> 1.A -> A**  (DE = 0.025*; IE = 0.163*)  % = 82.6  **EA -> 1.A -> A**  (DE = 0.025*; IE = 0.138*)  % = 84.6  **PN -> 1.A -> A**  (DE = 0.168*; IE = 0.156*)  % = 48.1 | 4 |
| **Powers et al.**  **(2011, USA)^64^** | 541  41  59% | Cross sectional | Subtype (EA)  CTQ | (A) PTSD (CAPS) | Mediation analysis | No  No | (1) Schizotypal personality disorder (SNAP) | **EA -> A -> 1**  (DE = 0.27*; IE = 0.07*)  % = 21 | 6 |
| **Rosen et al.**  **(2018, USA)^66^** | 61  NA  54% | Cross sectional | Composite  ACE | (1) ABH Negative Content (PSYRATS) (2) Depression | Regression Based mediation  Serial Mediation | Yes  Yes (age, gender, CPZ, antidepressant usage) | AVH Distress  (PSYRATS) | **ACE -> 1 -> AVH distress**  (DE = 0.11; IE = 0.14)  % = 53.8  All other mediation analyses were non significant. | 6 |
| **Schalinski *et al.* (2019, Germany)^67^** | 180 psychosis  28.6  31.7% | Cross sectional    Case control study | Composite (CT) and specific (abuse, neglect, neglect age 10)    MACE | (1) Dissociation (SDS) | Mediation Analysis | Yes    Yes (age, gender) | (1) Psychotic symptoms (PANSS) | **CT -> SDS-> positive symptoms**  Total mediation (DE = 0.20; IE = 0.07*)  % = 25.9    No mediating effects with specific trauma subscales | 6 |
| **Sengutta et al.**  **(2019, Germany)^68^** | 200  18.7  66.5% | Cross Sectional | Composite    ACE | (1) Borderline Personality features (BSL-23)    (2) Depression (PHQ-9)    (3) Anxiety (GAD-7) | Mediation Analysis | Yes    Yes (gender) | Psychotic Like Experiences (PQ-16) | **ACE -> BSL-23 -> PQ-16**  (DE = 0.12; IE = 0.198*)  % = 62.2    **ACE -> PH-9 -> PH-16**  (DE = 0.24*; IE = 0.1176*)  % = 32.9    **ACE -> GAD-7 -> PH-16**  (DE = 0.23*; IE = 0.0943)  % = 28.8 | 5 |
| **Seo et al.**  **(2018, South Korea)^69^** | 199  33.98  44.7% | Cross Sectional | Composite  CTQ | (A) Social Defeat (SD) | SEM | No  Yes (Sex, age) | (1) Paranoid Ideation (MMPI-2) | **CTQ -> A -> 1**  **(DE = 0.46*; IE = 0.13*)**  **% = 22** | 5 |
| **Seten-Suero et al. (2022, Spain)^31^** | 52 (HCs)  28.2  44%  278 (FEP)  30.6  46% | Prospective (3 years) | Composite  (CTES) | A. Recent stress-events (RSEp) | Logistic regression | Yes    Yes (age, sexe, education level, socioeconomic status, urbanism, employment, family history, cannabis use) | 1. FEP - Caseness | **CT -> a -> FEP**  (DE = 0.043; IE = 0.981*)  % = 96 | 8 |
| **Simpson et al. (2020, UK)^70^** | 78  37.64  77% | Cross sectional | Composite  CATS | a. Self-disgust | Mediation analysis | Yes  Yes (self-esteem, other as shame) | 1. Positive symptoms of psychosis (CAPE) | **CT -> a -> 1**  (DE = 0.02; IE = 0.07*)  % = 78 | 6 |
| **Steenkamp *et al.* (2022, Netherlands)^71^** | 50 NAP  31.8  42.4% | Cross sectional | Composite abuse (CA)    CECA.Q | (1) Loneliness (ESM)    (2) Depressive symptoms (ESM)    (3) Anxious Symptoms (ESM) | SEM | No    No | (A) Positive symptoms  (ESM) | **CA-> 1 -> A**  Total mediation (DE = -0.01; IE = 0.08*)  % = 11.9    No significant indirect effect of CT on  positive symptoms through depressive or anxious feelings | 4 |
| **Styla *et al.* (2019, Poland)^72^** | 45 SCZ  45 HC  42.4 SCZ  48.9% SCZ  41.78 HC  46.7% HC | Cross sectional    Case control study | Composite  Childhood adversities (CA)    CEQ | (1) Time Perspective (TP) (ZTPI) | Mediation analyses | Yes    Yes (education) | Caseness (SCZ) based on  ICD-10 | **CA -> TP -> SCZ**  Total mediation (DE = 0.352; IE = 0.249*)  % = 37 | 6 |
| **Sun *et al.* (2018, Australia)^32^** | 66 FEP  20.18  54.5% | Cross sectional | Composite and Binary    CTQ | (1) Dissociation (SCID-D-R) | Simple mediation analyses | Yes    No | (1) Positive symptoms (PANSS)  1.a -Hallucinations  1.b -  Delusions | **CTQ -> SCID-D-R -> 1.b**  Total mediation (DE = 0.01; IE = 0.02*)  % = 66.6    Non-significant mediation of dissociation between Ct and hallucinations | 5 |
| **Thompson *et al.* (2016, Australia)^40^** | 233 UHR  18.5 at baseline  48.8% | Prospective. Mean of 7.0 years (SD 3.2) follow-up | Sexual trauma (ST) score    CTQ | (1) Anxiety  (2) Depression  (3) Dissociation  (4) Mood swings  (5) Mania  (HAM-A, CAARMS) | Mediation analysis | No    No | (1) Transition to psychotic disorder  Based on CAARMS BPRS | Null mediation | 6 |
| **Uyan et al.**  **(2022, Turkey)^73^** | 100 SZO  15.38  51%  100 HC  14.9  65% | Cross Sectional | Composite  CTQ | (A) Dissociation (DES) | Mediation Model Regression Analysis | No  No | Caseness   (1) Schizophrenia Symptoms (PANSS) | **CTQ -> A -> 1**  (DE = 0.84*; IE = 0.04*)  % = 5 | 6 |
| **Van Dam *et al.* (2014, USA)^74^** | 131 psychosis  31.2  16% | Cross sectional | Composite    CTQ | (1) Attachment (PAM)  1.a - Avoidant  1.b - Anxiety | Regressions analyses following Baron and Kenny criteria | No    Yes (age and gender) | (1) Positive symptoms  (SAPS) | Null mediation | 4 |
| **Varese *et al.* (2012, UK)^81^** | 45 psychosis;  20 HC  44.6  46.7% | Cross sectional    Case control study | Composite (CATS) and specific (SA, PA, EA, N)    CATS | (1) Dissociation (DES) | Mediation analysis | Yes    No | (1) Hallucinations (LSHR-R) | **CATS -> dissociation -> hallucinations**  Partial mediation (DE = 0.15*; IE = 0.11*)  % = 42.3    **SA -> dissociation -> hallucinations**  Partial mediation (DE = 0.77*; IE = 0.57*)  % = 42.8    No mediating effects with specific trauma subscales | 5 |
| **Weijers *et al.* (2018, Netherlands)^76^** | 87 NAP  31.7  35.6% | Cross sectional | Abuse    CECA | (1) Mentalizing capabilities (MC) (HT) | Mediation analysis | Yes    No | (1) Positive PANSS | Null mediating effect with positive symptoms (only with negative partial mediation) | 5 |
| **Wickham and Bentall (2016, UK)^77^** | 72 psychosis; 72 HC  43.5  36.1% | Cross sectional    Case control study | Specific (childhood sexual abuse (SA) ; Childhood emotional neglect (EN) (bullying, PA, EA also initially explored)    CTQ and RBC | (1) Perception of injustice (BJW)  1.a - Personal  1.b - General | Mediation analysis | Yes    Yes (age, gender, SA, hallucinations and paranoia) | (1) Paranoia    (2) Hallucinations (PANSS) | **EN -> Personal ->Paranoia**  Partial mediation (DE = 0.11*; IE = 0.032*)  % = N/A    No mediating effect of general injustice on paranoia, or personal and general perception of injustice on hallucinations | 6 |
| **Xie et al. (2022, China)^35^** | 256 (HC)  23.71  18%  144 (FES)  23.50  36% | Cross-sectional | Composite  CTQ | a. IQ | Hierarchical regression  Mediation analysis | Yes  Yes (age, sex, BMI, education level) | 1. FES - Caseness | **CT → a → FES**  (DE = 0.31*; IE = 0.09*)  % = 22.5 | 5 |

ACE: Adverse Childhood Experiences; BAI: Beck Anxiety Inventory; BBTS: Brief Betrayal Trauma Survey; BCSS: Brief Core Schema Scale; BPRS: Brief Psychiatric Rating Scale; BMI: Body mass index; BS: Bullying Severity; BSI-53: Brief Symptom Inventory; BSL-23: Borderline Symptom Checklist; CAARMS: Comprehensive Assessment of At Risk Mental States; CA: Childhood Abuse; CADSS: Clinician-Administered Dissociative States Scale; CAPE: Community Assessment of Psychic Experiences; CAPS: Clinician-Administered PTDS Scale; CATS: Child Abuse and Trauma Scale; CECA: Childhood Experiences of Care and Abuse; CECA.Q: Childhood Experiences of Care and Abuse Questionnaire; CEQ: Childhood Experiences Questionnaire; CFQ: Chalder Fatigue Questionnaire; CT BBTS: Childhood trauma subscale of the Brief Betrayal Trauma Survey; CTES: Childhood Traumatic Events Scale; CTQ: Childhood Trauma Questionnaire; CTQ-SF: Childhood Trauma Questionnaire-Short Form; DAWBA: Development and Well-Being Assessment; DD: Dissociative Disorders; DES-II: Dissociative Experience Scale; DES-R: Dissociative Experiences Scale—Revised; DID Dissociative Identify Disorder; DSS: Dissociative Symptoms Scale; DUP: Duration of Untreated Psychosis; EA: Emotional Abuse; ECR-R: Experiences in Close Relationships-Revised Questionnaire; ELES: Early Life Experience Scale; EN: Emotional Neglect; ESM: Experience Sampling Method; ETI: Early Trauma Inventory; FEP: First Episode of Psychosis; FES: First-Episode Schizophrenia; GAD-7: Generalized Anxiety Disorder Questionnaire; GSEM: Generalized Structural Equation Model; HAM-A: Hamilton Anxiety Rating Scale; HT: Hinting Task; ICD-10: International Classification of Diseases, 10th revision; IDS-C: Inventory of Depressive Symptoms – Clinician-rated scale; IESR: Impact Event Scale-Revised; IPSM: Interpersonal sensitivity scale; IQ: Intelligence Quotient; LSHS: Launay–Slade Hallucination Scale; LSHS-R: Launay Slade Hallucinations Scale-Revised; MACE: Maltreatment and Abuse Chronology of Exposure Scale; MADRS: Montgomery Asberg Depression Rating Scale; MCQ: Metacognitions Questionnaire; MINI: Mini-International Neuropsychiatric Interview 5.0.0; MINI 7: Mini International Neuropsychiatric Interview, 7th Edition; MMPI: Minnesota Multiphasic Personality Inventory-2; MMSE: Mini-Mental State Examination; MRC: MRC Sociodemographic Schedule; MSI: McLean Screening Instrument; MUPS: Mental Health Research Institute Unusual Perceptions Schedule; N: Neglect; NSO: Negative Other; NSS: Negative Self Schemas; PA: Physical Abuse; PaDS: Persecution and Deservedness Scale; PAM: Psychosis Attachment Measure; PAM-R: Psychosis Attachment Measure-Revised; PANSS: Positive and Negative Syndrome Scale; PDI: Peters et al. Delusions Inventory; PHQ-9: Patient Health Questionnaire; PLS-SEM: Partial least squares structural equation model; PN: Physical Neglect; PQ-16: Prodromal Questionnaire; PSP: Personal and Social Performance Scale; PSQ: Psychosis Screening Questionnaire; PSYRATS: Psychotic Symptom Rating Scales; PTCI: Posttraumatic Cognitions Inventory; RBQ: Retrospective Bullying Questionnaire; REF: Ideas of Reference; RS-14: Resilience Scale Short Version; RSEp: Recent Stressful Events(perception); RSEQ: Rosenberg Self-Esteem Questionnaire; RSES: Rosenberg Self-Esteem Scale; SA: Sexual Abuse; SANS: Scale for the Assessment of Negative Symptoms; SAPS: Scales for Assessment of Positive Symptoms; SCID-D-R: Structured Clinical Interview for DSM-IV Dissociative Disorders; SCSS: Self-Concept Clarity Scale; SCZ: Schizophrenia; SD: Social defeat; SDS: Shutdown Dissociation Scale; SEM: Structural Equation Modelling; SNAP: Schedule for Nonadaptive and Adaptive Personality; SSPS: State Social Paranoia Scale; SRLE: Survey of Recent Life Experiences; SRS-PTSD: Self-Rating Scale for Post-traumatic Stress Disorder; SSP: Schizophrenia-Spectrum Psychosis; STIPO: Structured Interview of Personality Organization; SUD: Substance Use Disorders; TADS: Trauma and Distress Scale; THQ: Trauma History Questionnaire; TQ: Trauma Questionnaire; ZTPI: Zimbardo Time Perspective Inventory.

### Table S5. Overview of Biological Mediators examined in clinical studies included in this review

| **Authors**  **Country** | **Sample**  **Mean Age**  **% female** | **Design** | **Measures of childhood adversity** | **Mediator(s)** | **Analysis** | **Bootstrap**  **(yes / no) / confounders (yes / no)** | **Psychosis** | **Main findings**  **Pathway**  **Total / partial mediation**  **Direct Effect (DE)**  **Indirect effect (IE)**  **% total effect mediated** | **Quality Score** |
| --- | --- | --- | --- | --- | --- | --- | --- | --- | --- |
| **Alameda et al. (2023, Europe)^27^** | 883  (366 FEP, 517 controls)  35.33  45.64% | Prospective | Composite & subtypes (A & N)  CTQ | a. DNAm | Mediation analysis (DACT approach) | Not mentioned  Yes (age, sex, smoking, medication, cell-type composition, batch effect) | 1. Psychosis (Caseness) | Adversity, abuse and neglect significantly increase the risk of psychosis. Odds ratios (OR): Overall adversity (1.68), Abuse (2.16), Neglect (2.27).  None of the CpG sites displayed significant mediations when strict corrections were used.  **CT composite -> a -> 1**  28 normally significant severe adversity-associated DMPs (P < 5e-5) for mediation were identified (spans across 21 genes).  **CT abuse -> a -> 1**  34 normally significant severe adversity-associated DMPs (P < 5e-5) for mediation were identified (spans across 27 genes).  **CT neglect -> a -> 1**  29 normally significant severe adversity-associated DMPs (P < 5e-5) for mediation were identified (spans across 20 genes).  Abuse and neglect did not have overlapping mediating genes. | 7 |
| **Bhui et al. (2021, UK)^44^** | 134 456  55.72  54.6%  480 (psychosis) | Prospective cohort study | Composite (past adversity)  ACE self-report | b. Biomarkers | PLS--SEM | Yes  No | 1. Caseness (ICD-10) | **CT → b → 1**  (DE = 0.028; IE = 0.001)  % = 4 | 5 |
| **Corcoran et al. (2020, UK)^49^** | 41 (HCs)  47 (psychosis)  39.5  % NA | Cross-sectional | Composite  CLEQ | a. Working performances  b. Glutamate  (during resting state in the bilateral DLPFC and the ACC) | Mediation analysis | No  Yes (age, sexe) | 1. psychosis - Caseness  (PANSS) | Null Mediations | 4 |
| **Rapado-Castro et al. (2020, Australia)^39^** | 416 Participants  62 Included Sample  38 UHR-NT  24 UHR-T  20.2  58.1 % | Prospective | Subtype (SA)  CTQ | a. Brain activation   (Right middle temporal gyrus) | Mediation analysis | Yes  Yes (age, sex, medication, psychopathology, QI, quality of life, premorbid functioning, DUP, year entry in PACE) | 1. Transition to psychosis (CAARMS & SCID) | **SA -> a -> 1**  (DE = NR; IE = 0.4)  % = 9.2  2nd Analysis (controlled for confounding covariates): The mediation effect was no longer significant. | 7 |
| **Wang et al. (China, 2023)^34^** | 76 (HC)  29.56  44%  79 (FEP)  28.24  48% | Cross sectional | Composite & Subtypes (EN, PN) | a. Sensory gating deficits (P50) | Meditation analysis | Yes  Yes (sex, age, education level) | 1. First episode schizophrenia - Caseness (PFES) (DSM-IV) | **EN -> a -> 1**  (DE = 0.10*; IE = 0.03)  % = 23  No mediation effects on remaining variables. | 5 |
| **Wang et al. (2022, China)^33^** | 136 (HCs)  28.6  44.8%  192 (FEP)  27.2  50% | Cross-sectional | Composite & subtypes (total, EN)  CTQ | a. BDNF levels | Mediation analysis | Yes    Yes (sex, education level) | 1. Psychosis (PANSS) total score | **CT total -> a -> 1.b**  (DE = 0.53*; IE = 0.061)  % = 10    **EN -> a -> 1.b**  (DE = 0.78*; IE = 0.013)  % = 14 | 6 |
| **Cancel *et al.* (2015, France)^45^** | 21 SZO; 30 HC  32.1 SZO  29% SZO  32.9 HC  33% HC | Cross sectional | Subscales (EN)    CTQ | (1) Grey matter volume in DLPFC | Regression based approach and SEM | No    Yes  (duration of illness and parents’ education levels) | (1) Disorganization (SANS) | **EN -> DLPFC -> Disorganization**  Suggested mediation  % = NA | 4 |
| **Quidé *et al.* (2018, Australia)^65^** | 112 psychosis;  53 HC  38 psychosis  47% psychosis    38.7 HC  39.6% HC | Cross sectional    Case control study | Composite (CTQ)    CTQ | (1) Inferior frontal gyrus (IFG) activation | Mediation analysis | Yes    No | (1) Positive symptoms (PANSS) | **CTQ -> IFG activation -> PANSS positive**  Null mediation | 4 |

BDNF: Brain-Derived Neurotrophic Factor; CAARMS: Comprehensive Assessment of At Risk Mental States; CLEQ: Childhood Life Events Questionnaire; CTQ: Childhood Trauma Questionnaire; DACT: Divide-Aggregate Composite-null Test; DLPFC: Dorsolateral Prefrontal Cortex; DNAm: DNA-methylation; EA: Emotional Abuse; IFGL: Inferior Frontal Gyrus; PANSS: Positive and Negative Syndrome Scale; PFES: Patients with First-Episode Schizophrenia; PN: Physical Neglect; SA: Sexual Abuse; SEM: Structural Equation Modelling;

### Table S6. Overview of Psychological Mediators in Non-clinical studies included in this review

| **Authors**  **Country** | **Sample**  **Mean Age**  **% female**  **Sample type** | **Design** | **Measures of childhood adversity** | **Mediator(s)** | **Analysis** | **Bootstrap**  **(yes / no) / confounders (yes / no)** | **Psychosis** | **Main findings**  **Pathway**  **Total / partial mediation**  **Direct Effect (DE)**  **Indirect effect (IE)**  **% total effect mediated** | **Quality Score** |
| --- | --- | --- | --- | --- | --- | --- | --- | --- | --- |
| **Ashford *et al.* (2012, UK)^12^** | 135  19.8  91.1% | Cross sectional | Bullying subscales: direct physical aggression (DPA); direct verbal aggression (DVA); Indirect aggression (IA) 0    DIAS | (1) Interpersonal sensitivity (IPSM)    (2) Anxiety (HADS)    (3) Depression (HADS)    (4) Negative core beliefs (BCSS)  4.a - Negative self beliefs  4.b - Negative beliefs others | Multiple mediation | Yes    Yes (ethnicity, gender and other bullying categories) | (a) Paranoia (ideas of social reference) (GPTS)    (b) Paranoia  (persecution)  (GPTS) | **IA -> Depression -> a**  Suggested mediation (IE = 0.11; DE = N/A  % = N/A    **IA -> Negative self beliefs -> a**  Suggested mediation  (IE = 0.16; DE = N/A)  % = N/A    **IA -> Depression-> b**  Suggested mediation  (IE = 0.10; DE = N/A)  % = N/A    **IA -> Negative self Beliefs -> b**  Suggested mediation  (IE = 0.18; DE = N/A)  % = N/A    **DVA -> Negative beliefs others-> a**  Suggested mediation  (IE = 0.24; DE = N/A)  % = N/A    **DVA -> Negative beliefs others-> b**  Suggested mediation (IE = 0.11; DE = N/A)  % = N/A | 5 |
| **Adewuya et al. (2025, Nigeria)^41^** | 1953  945 PLEs  95 psychosis  1008 controls  34.7  % NR | Cross sectional | Composite & subtypes (composite abuse, composite neglect)  CTQ-sf | A.Common mental disorders(Depression (PHQ-9), anxiety (GAD-7), somatic symptoms)  B. resilience (RS-14)  C. self esteem (RSEQ)  D. Cognitive functioning (MMSE) | Generalized structural equation modeling (GSEM) | Yes  confounders : age, gender, socioeconomic position | 1. PLEs  (PQ-B) | CT → A → 1  (DE = 0.225; IE = 0.048)  % = 21.3  CT → B → 1  (DE = 0.225; IE = 0.052)  % = 23.1  CT → C → 1  (DE = 0.225; IE = 0.021)  N.S  CT → D → 1  (DE = 0.225; IE = 0.041)  % = 18.2 | 6 |
| **Bellido-Zanin et al.**  **(2018, Spain)^6^** | 472  25.5  73.3% | Cross Sectional | Composite  ELES | (A) Dissociation (DES-II)   A.1 amnesia  A.2 Absorption  A.3 Depersonalisation | Mediation Analysis | No  Yes (hallucination proneness/ ideas of reference) | (1) Hallucination proneness (LSHS-R) (2) Ideas of reference (REF) | **ELES -> A -> 1**  Total Mediation  (DE = 0.08; IE = 0.07*)  % = 47  **ELES -> A.1 + A.2 + A.3 = 1**  Total Mediation  (DE = 0.06; IE A.2 = 0.05*; IE A.3 = 0.04*)  % A.2 = 33.3  % A.3 = 27  **ELES -> A -> 2**  Partial Mediation  (DE = 0.13*; IE = 0.03*)  % = 19 | 6 |
| **Berenbaum et al.**  **(2008, USA)^7^** | 306  43.2  53.1% | Cross Sectional | Composite  (Interview) | (A) lifetime PTSD (CAPS) (B) PTSD symptoms (CAPS) (C) Absorption (DPS; CES) (D) Dissociation (SCID; DES-T) | Mediation analysis (Sobel test) | No  No | (1) Schizotypal traits (PDI-IV) | PTSD and absorption/ dissociation partially mediated the relationship between childhood adversity and schizotypy. | 5 |
| **Bhavsar et al.**  **(2019, UK)^82^** | 1698  NR  56.5% | Cross Sectional | Subtype (Abuse)  Interview | (A) Adverse life events (SELCoH)   A.1 Violent  A.2 Non Violent | Mediation Analysis | No  Yes (sex, age, ethnicity, education, social class) | (1) Psychosis (PSQ) | **Abuse -> A.1 + A.2 -> 1**  (OR DE = 1.53*; OR IE = 1.51* ; OR IE A.1 = 1.34*; IE A.2 = 1.13*)  % Total = 47  % A.1 = 33  % A.2 = 14 | 6 |
| **Blose et al.**  **(2023, USA)^83^** | 346  19.10  52 | Retrospective | Composite  CTQ | (1) Dissociation (DES II)  (2) Peritraumatic Dissociation (PDEQ) | Mediation Analysis | Yes  Yes (Gender) | Schizotypal Personality (SPQ) | **CTQ -> 1 -> SPQ**  (DE = 0.05; IE = 0.0644*)  % = 58.5  **CTQ -> 2 -> SPQ**  N.S | 6 |
| **Bortolon *et al.* (2017, France)^8^** | 425  36.23  79.1% | Cross sectional | Specific (PA, EA)    CTQ | (1) Maladaptive schemas (MS)  (SQ-SF):  1.a - Abandonment    (2) Dissociation  (DES):  2.a - Defensive dissociation | PLS-SEM | Yes    Yes (age, gender, psychopathology) | (1) Auditory hallucination (AH) (LSHS-R) | **PA -> 2.a -> AH**  Suggested mediation (DE = N/A; IE = 0.1081*)  % = N/A    **EA ->1.a-> AH**  Suggested mediation  (DE = N/A; IE = 0.055*)  % = N/A | 5 |
| **Bortolon and Raffard (2018, France)^84^** | 403  33.24  82.1% | Cross sectional | Composite (CT)    CTQ | (1) Defensive dissociation  (DES) | SEM | Yes    Yes (age, gender, anxiety, depression) | (A) Seeing visions (LSHS)    (B) Hearing voices  (LSHS) | **CT-> 1 -> A**  Total mediation (DE = 0.008; IE = 0.122*)  % = 93.7    **CT-> 2 -> B**  Partial mediation  (DE = 0.090*; IE = 0.124*)  % = 57.94 | 6 |
| **Bortolon & Raffard**  **(2019, France)^85^** | 179  24  83.4 | Cross Sectional | Composite  ACE | (1) Avoidance  (IES-R)  (2) Intrusions  (IES-R)  (3) Shame  (ESS) | Regression Analysis | Yes  Yes (gender, age) | Hallucination proneness  (LSHS) | **ACE -> Avoidance -> LSHS**  N.S  **ACE -> Intrusions -> LSHS**  (DE = 1.185; IE = 0.707*)  % = 37.3  **ACE -> Shame -> LSHS**  (DE = 1.185 ; IE = 0.386*)  % = 17.3 | 5 |
| **Boyda and McFeeters (2015, Northern Ireland)^86^** | 7403  46  51.4% | Cross sectional | Specific (SA, EN)    Questionnaire | (1) Social functioning  1.a - Loneliness  (NA) | Logistic mediation analyses | Yes    No | Psychotic-like experiences (PSQ) | **SA -> Loneliness -> PSQ**  Total mediation (DE = 1.38; IE = 0.22*)  % = N/A    Total mediation    **EN -> Loneliness-> PSQ**  Total mediation (DE = 0.94; IE = 0.46*)  % = N/A | 5 |
| **Boyda *et al.* (2018, UK)^13^** | 302  36  69.9% | Cross sectional | Composite, subscales (EA, SA)    ACE-IQ | (1) Early Maladaptive Schemas (EMS)  (YSQ-SF)  1.a – Defectiveness / Shame  1.b – Dependency / Incompetence  1.c – Enmeshment / Undeveloped self  1.d - Emotional inhibition | Multiple mediation  analyses | No    Yes (age, gender, urbanicity, ethnicity, socio-economic status, drug use) | (1) Psychotic experiences (PE)  (CAPE) | **EA -> 1.b -> PE**  Suggested mediation (DE = N/A; IE = 0.083*)  % = N/A    **SA-> 1.b-> PE**  Suggested mediation (DE = N/A; IE = 0.073*)  % = N/A    **EA ->1.c-> PE**  Suggested mediation  (DE = N/A; IE = 0.063*)  % = N/A    **SA -> 1.c -> PE**  Suggested mediation (DE = NA; IE = 0.043*)  % = N/A | 5 |
| **Chen et al.**  **(2022, China)^87^** | 262  19.73  63.7 | Cross Sectional | Subtypes  RBQ (quantity, duration) | (1) Self Esteem (RSES)  (2) Personality  (EPQ-S)  (3) Interpretation Bias (AST) | Multiple Mediation | Yes  Yes (age, gender, education) | Psychotic  Experiences (CAPE) | **Quantity -> 1 -> CAPE**  N.S  **Quantity -> 2 -> CAPE**  (DE = 0.03;* IE = 0.0278*)  % = 40%  **Quantity -> 3 -> CAPE**  N.S  **Duration-> 1 -> CAPE**  N.S  **Duration -> 2 -> CAPE**  (DE = 0.08*; IE = 0.0350*)  % = 27.1%  **Duration -> 3 -> CAPE**  (DE = 0.08*; IE = 0.0114*)  % = 8.7% | 6 |
| **Cole *et al.* (2016, UK)^88^** | 200  19.96  82.5% | Cross sectional | Composite Childhood maltreatment (CM)    CATS | (1) Dissociation:  1.a - Dissociative amnesia (DA) (DES-II)    1.b1 -Depersonalization-  (CDS)    1.b2 - Depersonalization (DES-II)    1.c - Absorption (Abs)  (DES-II) | (A) Simple and    (B) multiple mediation analysis | Yes    No | (1) Hallucination-proneness (HP) (LSHS-R)    (2) Delusional ideation (DI)  (PDI) | (A) Simple  **CM ->Dissociation-> HP**  Partial mediation  (DE = 2.92*; IE = 3.94*)  % = 57.4    **CM ->Dissociation -> DI**  Partial mediation (DE = 10.90*; IE = 10.75*)  % = 49.6    (B) Multiple  **CM -> Abs -> HP**  Total mediation (DE = 1.64; IE = 3.45*)  %=50.29    **CM -> DA -> DI**  Partial mediation  (DE = 7.45*; IE = -3.68*)  % = -16.99    **CM-> Abs -> DI**  Partial mediation  (DE = 7.45*; IE = 7.18*)  % = 33.1    Remaining items within dissociation were not significant | 5 |
| **Croft et al.**  **(2022, UK)^89^** | 3360  NR  63 | Longitudinal | Composite  CT Questionnaire | (1) Belief updating  (DTD Task)  1.a Decision Noise | Counterfactual mediation analysis | No  Yes (Maternal education) | Psychotic Like experiences  (SIPS & CAARMS) | **CT -> DTD -> PLEs**  Null Mediation | 7 |
| **Dhondt et al.**  **(2022, Ireland) ^90^** | 6039  NR  49.31 | Longitudinal | Composite  (Parent report questions) | (1) Affective dysregulation - Internalising Problems (SDQ)  (2) Affective dysregulation -  Externalising Problems (SDQ)  (3) Self Concept (PHS)  (4) Parent Child Conflict  (5) Feelings of Safety | Mediation Analysis | No  Yes  (age, gender, socio-economic status, minority status, Urbanicity) | Psychotic Experiences  (APSS) | **CA -> 1 +3 -> APSS**  (OR DE = 1.86*; OR IE 1 = 1.05*, OR IE 3 = 1.04*)  % total = 12.97  % 1 = 7.01  % 3= 5.96  All other mediators were non-significant. | 7 |
| **Fekih-Romdhane et al.**  **(2024, Lebanon)^91^** | 4158  21.91  64.4 | Cross Sectional | Cyberbullying Composite  (RCBI-II) | (1) Insomnia severity  (ISI) | Mediation Analysis | No  Yes (Country, sociodemographic variables, substance use) | Positive Psychotic experiences  (CAPE-42) | **RCBI -> 1 -> CAPE-42**  (DE = 0.56*, IE = 0.10*)  % = 15.5 | 6 |
| **Fisher *et al.* (2012, UK)^92^** | 212  27  65.4% | Cross sectional | Specific (EA, PA)    CTQ | (1) Depression (BDI)    (2)Anxiety (BAI)    (3) Negative schematic beliefs (BCSS)  3.a - Negative self-schemas  3.b - Negative others schemas | Mediation analysis | Yes    Yes (gender, age, ethnicity, family history) | (1) Paranoia (PSQ) | **EA -> Anxiety -> Paranoia**  Total mediation (DE = 1.16; IE = 1.05*)  % = 17.57    No mediation effects of depression, negative self-schemas and other schemas | 6 |
| **Fisher *et al.* (2013, UK)^14^** | 6692  12.9  50.9% | Prospective    From childhood (8, 21, 33, 47, 61, 73 months) to mean age of 12.9 | Specific (Harsh parenting (HP), domestic violence (DV) and bullying victimization (BV))    Questionnaire to mothers ; Bullying and Friendship Interview Schedule (BI) | (1) External locus of control (LoC) (12 item version of NSIE)    (2) Self Esteem (shortened form of Harter`s Self Perception Profile for Children)    (3) Affective symptoms (DAWBA and SMFQ)  3.a - Anxiety  3.b - Depression | Multiple mediation analysis | Yes    Yes (gender, ethnicity, birth weight, family history of schizophrenia, depression or suicide, child’s IQ, and general family adversity) | (1) Psychotic Symptoms (PS)(PLIKSi) | **HP -> Anxiety-> PS**  Total mediation (DE = 1.02; IE = 1.01*)  % = 21    **DV -> Anxiety ->PS**  Total mediation (DE = 1.06; IE = 1.00*)  % = 8    **BV -> Anxiety -> PS**  Partial mediation (DE = 1.14*; IE = 1.00*)  % = 2    **HP-> Depression ->PS**  Total mediation (DE = 1.00; IE = 1.03*)  % = 94    **DV -> Depression -> PS**  Total mediation (DE = 1.04; IE = 1.01*)  % = 18    **BV -> Depression -> PS**  Partial mediation (DE=1.14*; IE = 1.01*)  % = 8    **HP -> LoC -> PS**  Total mediation (DE = 1.01; IE = 1.01*)  % = 47    **DV-> LoC -> PS**  Total mediation (DE = 1.03; IE = 1.00*)  % = 1    **BV -> LoC -> PS**  Partial mediation (DE = 1.13*; IE = 1.02*)  % = 13    **HP -> Self-esteem -> PS**  Total mediation (DE = 1.00; IE = 1.01*)  % = 97    **DV -> self-esteem-> PS**  Total mediation (DE = 1.04; IE = 1.00*)  % = 7      **BV -> Self esteem-> PS**  Partial mediation (DE = 1.13*; IE = 1.01*)  % = 10    **HP -> all mediators-> PS**  Total mediation (DE = 0.97; IE = 1.04*)  % = 100    **BV -> all mediators-> PS**  Partial mediation (DE = 1.10*; IE = 1.04*)  % = 29    **DV-> all mediators -> PS**  Total mediation (DE = 1.03; IE = 1.02*)  % = 42 | 8 |
| **Frydecka et al.**  **(2020, Poland)^93^** | 6772  26.6  61.2 | Cross Sectional | Composite CA & Subscales  TEC (EA, EN, PA)  CECA.Q (SA) | (1) Cognitive biases (DOCOBS-18) | Serial Mediation analysis | Yes  Yes (age, sex, education) | Psychotic Like Experiences (PQ-16) | **EA-> 1 -> PLE**  (DE = 1.6438*; IE = 0.5237)  % = 24  **EN-> 1 -> PLE**  (DE = 1.1437*; IE = 0.4900)  % = 30  **PA-> 1 -> PLE**  (DE = 1.4383*; IE = 0.4520)  % = 24  **SA-> 1 -> PLE**  (DE = 1.1598*; IE = 0.7448)  % = 39  **CT -> 1 -> PLE**  (DE = 1.16244*; IE = 0.5496)  % = 25 | 6 |
| **Fung et al.**  **(2024, Hong Kong)^94^** | 468  25.6  91% | Cross Sectional | Composite  BBTS (Childhood) | (A) Dissociation (MDI)   A.1 Disengagement  A.2 Identity dissociation  A.3 Emotional Constriction  A.4 Memory Disturbance  A.5 Depersonalisation  A.6 Derealization   (B) Somatoform Dissociative Symptoms (SDQ-5)   (C )International Trauma Questionnaire  C.1 Experiencing  C.2 Avoidance  C.3 Sense current threat  C.4 Affective dysregulation  C.5 Negative Self concept  C.6 Disturbances in relationships | Multiple Mediation Analysis | Yes  No | (1) Psychic Experiences   (CAPE-P)  1.1 Bizarre Experiences  1.2 Delusional ideations  1.3 Perceptual Anomalies | **BBTS -> A.4 + C.3 + A.2 -> 1.2**  (DE = 0.16*; IE = 0.06*; IE A.4 = 0.086*; IE C.3 = 0.052*; IE A.2 = 0.049)  Total % = 28.6  A.4 % = 40.9  C.3 % = 24.8  A.2 % = 23.4  **BBTS -> A.2 + A.4 + B -> 1.1**  (DE = 0.06; Total IE = N.S; IE A.2= 0.101*; IE A.4 = 0.076*; IE B = 0.035*)  Total % = N.S  A.2 % = 33.7  A.4 % = 25.3  B % = 11.7  **BBTS -> A.2 + B + A.6 -> 1.3**  (DE = 0.11*; IE = 0.21; IE A.2 = 0.122*; IE B = 0.122*; IE A.6 = 0.051)  Total % = 65.6  A.2 % = 38.1  B % = 38.1  A.6 % = 16 | 5 |
| **Gaweda et al.**  **(2020, Poland)^95^** | 3495  51.1  62.9% | Cross Sectional | Composite  TEC & CECA.Q | Cognitive biases (DOCOBS-18) | SEM | Yes  No | Psychotic Like Experiences (PQ-16) | **CT-> DOCOBS -> PLEs**  (DE = 0.38*; IE = 0.211*)  % = 35.7 | 5 |
| **Gaweda *et al.* (2019, Germany)^96^** | 649  51.1  55.2% | Cross sectional | Composite (CT), Abuse, neglect    CTQ | (1) Aberrant salience Inventory (ASI)    (2) Anomalous self-experiences (IPASE) | Parallel multiple mediation models | Yes    Yes (gender) | Psychotic-like experiences  (PLE) (PQ) | **CT -> 1 + 2 ->PLE**  Total mediation (DE = 0.05; IE 1 = 0.076*; IE 2 = 0.1443*)  % 1 = 28.1  % 2 = 53.4    **Neglect -> 1 + 2 -> PLE**  Total mediation (DE = 0.01; IE 1 = 0.0451*; IE 2 = 0.1404*)  % 1 =26.5  % 2 =82.58    **Abuse -> 1 + 2-> PLE**  Partial mediation (DE = 0.10*; IE 1 = 0.092*; IE 2 = 0.1188*)  % 1 = 29.67  % 2 = 38.32 | 6 |
| **Gibson *et al.* (2019, USA)^97^** | 945  20.13  75.6% | Cross sectional | Composite (CT)  CTQ | (1) Perceived Stress  (PSS)    (2) Dissociation (DES)    (3) Cognitive bias  3.1 Negative self-schemas (BCSS)  3.2 Negative others- schemas (BCSS)  3.3 External locus of control  (RI-E) | Multiple mediation analyses | Yes    Yes (gender, race, age) | (A) Psychotic-Like Experiences  (PLE) (PQ) | **Multiple mediation**  **CT-> 1 -> PLE**  Total mediation (DE = 0.04; IE = 0.0405*)  % = 25.31    **CT-> 2-> PLE**  Total mediation (DE = 0.04; IE = 0.054*)  % = 33.7  **CT -> 3.1 -> PLE**  Total mediation (DE = 0.04; IE = 0.007*)  % = 14.43    **CT -> 3.2 -> PLE**  Total mediation (DE = 0.04; IE = 0.0141*)  % = 8.81  **CT -> 3.3-> PLE**  Total mediation (DE = 0.04; IE = 0.0071*)  % = 4.43    Only multiple mediation analyses presented and considered as the total effect of simple analyses are missing | 6 |
| **Goldstone et al.**  **(2011, Australia)^52^** | 100 psychosis  NA  44%  133 Non Clinical  NA  59% | Cross sectional (Emotional Trauma and Sexual Trauma) | Subtypes (Emotional Trauma and Sexual Trauma)  ETI | (B) Life hassles (SRLEs) | Path analysis | No  No | (2) Delusions   (PDI) | **Non Clinical**  **EA -> A -> 1**  (DE = NA; IE = 0.14*)  % = NA |  |
| **Goldstone et al.**  **(2012, Australia)^24^** | 100 Psychosis  NA  44%  133 Non Clinical  NA  59% | Cross sectional | Subtypes (Emotional Trauma and Sexual Trauma)  ETI | Life hassles (SRLEs)  Metacognitive beliefs (MCQ) | Path Analysis | No  No | A.1  Hallucinations (LSHS-R)  A.2 Auditory Hallucinations  (LSHS-R) | **Non Clinical**  **(EA -> A -> A.1)**  (DE = NA; IE = 0.1*)  % = NA  **Non Clinical**  **(EA -> A -> A.2**  (DE = NA; IE = 0.06*)  % = NA |  |
| **Gomez & Freyd.**  **(2017, USA**)^98^ | 192  19.81  65% | Cross Sectional | Composite SA  (SES) | (1) Dissociation (CES) | Mediation Analysis | Yes  No | (A) Hallucinations (CIDI -B&Es) | **SA -> 1 -> A**  (DE = 0.232**; IE = 0.168*)  % = 42 | 5 |
| **Goodall *et al.* (2015, UK)^99^** | 283  26.8  72% | Cross sectional | EA analysed in mediation    CTQ | (1) Adult Attachment (ECR-R)  1.a - Attachment avoidance (AAv)  1.b - Attachment anxiety (AAn) | Parallel multiple mediation analysis | Yes    No | (1) Schizotypy  (SPQ-B) | **EA -> AAv -> Schizotypy**  Partial mediation (DE = 0.35*; IE = 0.04*)  % = 13    **EA -> Aan -> Schizotypy**  Partial mediation (DE = 0.35*; IE =0.06*)  % = 8 | 5 |
| **Huang et al. (China, 2023)^18^** | 2808  General population  22.66  78% | Cross sectional | Composite & Subtypes (EN, PN, SA, PA, EA)  CTQ-SF | Depressive Symptoms (BDI)  Motivation and Pleasure (MAP-SR)  Bipolar Traits (BIP2) | Network analysis | Yes  No | Schizotypy (MSS)  1.A. Positive Schizotype | Childhood trauma was closely connected with schizotypy and motivation. | 4 |
| **Jaya *et al.* (2016, Germany)^15^** | 2350  32.5  37% | Cross sectional | Composite of Social Adversity (SocA) including bullying and abuse    BVQ /NEMESIS | (1) Social rank with the Social Comparison Scale (SCS)    (2) Negative schemas  Brief Core Schema Scales (BCSS)    (3) Loneliness  (UCLA) | SEM | Yes    No | (1) Positive (PS) and negative symptoms (CAPE)    (only positive considered) | **SocA-> Loneliness -> PS**  Suggested mediation (DE = N/A; IE = 0.02*)  % = N/A    **SocA -> BCSS-> PS**  Suggested mediation (DE = N/A; IE = 0.12*)  % = N/A    No mediation for SCS on positive symptoms | 4 |
| **Jin et al.**  **(2022, China)^16^** | 3479  19.67  51.2 | Cross Sectional | Subscales (CSA)  CTQ | (1) PTSD (TSQ)  (2) Depression (PHQ-9)  (3) Anxiety (GAD-7) | Network Analysis | Yes  No | Psychosis (PS) | Psychosis, depression, anxiety, and PTSD symptoms were positively correlated.  The node anxiety had the highest strength centrality, followed by ‘emotional cue reactivity (PTSD)’ and depression. | 4 |
| **Jones et al.**  **(2024, UK)^100^** | 227  23  58.8 | Cross Sectional | Composite  STR | Combined Dissociation Score  (DES, shut-D) | Mediation Analysis | Yes  No | Hallucination proneness  (RHS) | **STR -> Dissociation -> RHS**  (DE = 0.16, IE = 0.9*)  % = 85 | 5 |
| **Levi et al. (2025, UK)^9^** | 609 (259 psychosis, 350 GP)  28.70  % 25 | Cross sectional | Subtypes (PA, SA, EA)  BBTS | A.insecure attachment (PAM-R)  B.dissociation (DES)  C.negative schemas (BCSS) | Network analysis | Yes  No | Psychotic experiences (CAPE-42) | **Network analysis**  Childhood trauma variables in the no psychosis diagnosis network formed as an isolated cluster. | 4 |
| **Lincoln *et al.* (2017, Germany)^101^** | 562  24.31  49.3% | Prospective (follow up at 4, 8, 12 months) | Composite (CT)    NEMESIS | (1) Emotion regulation (ER)  (ERSQ) | SEM | Yes    No | (1) Subthreshold psychotic experience (CAPE)  1.a - Distress  1.b - Frequency | **CT -> ER-> Distress**  Partial mediation (DE = 0.069*; IE = 0.005*)  % = 7.1    No mediating effect for symptom frequency | 6 |
| **Marwaha *et al.* (2014, UK)^102^** | 8580 + 7403 Baseline  2406 FU  NA  NA | Cross sectional | Sexual abuse (SA)    Questionnaire | (1) Mood instability (MI) (BPD section of the SCID-II) | Mediation analysis | No    Yes (age, gender, marital status, employment status and ethnicity, PTSD, current affective state and hypomanic symptoms) | (1) Psychotic  Phenomena (PSQ)  1.a - Probably psychosis (PP)  1.b - Paranoid ideation (PI)  1.c - Auditory Hallucinations (AH) | **SA -> MI -> PP**  Suggested mediation  % = 34.6    **SA -> MI -> PI**  Suggested mediation  % = 34.5    **SA-> MI ->AH**  Suggested mediation  % = 25.3 | 4 |
| **Marwaha and Bebbington (2015, UK)^103^** | 5689  N/A  N/A | Cross-sectional | Sexual abuse (Non consensual intercourse (NCI), contact abuse (CA))    Questionnaire | (1) Anxiety (CIS-R)    (2) Depressive symptoms (CIS-R)    (CIS-R) (analyzed together) | Mediation analysis | No    Yes (gender, age, ethnicity, education, being brough by both partens until 16) | (1) Psychotic symptoms (PSQ) | **NCI -> 1, 2 -> PSQ**  Partial mediation (DE = 4.08*; IE (together) = 2.41*)  % anxiety = 20.4  % depression = 37.4    **CA-> 1, 2-> PSQ**  Total mediation (DE = 2.14; IE (together) = 1.60*)  % anxiety = 24.1  % depression = 37.1    IE shows combined effects of 1, 2  Percentages shows mediators examined separately | 5 |
| **McCarthy-Jones (2018, Ireland)^104^** | 5788  51.71  55.4% | Cross sectional | Childhood Sexual abuse (CSA)    Questionnaire from APMS | (1) Anxiety (CIS-R)    (2) Obsessional thought (CIS-R)    (3) Compulsions (CIS-R)    (4) PTSD (APMS questionnaire)    (5) Depression (CIS-R) | Regression-based approach | Yes    Yes (age, gender, ethnicity, education, IQ, depression) | Auditory verbal hallucinations (AVH) (PSQ) | **CSA-> 3 -> AVH**  Partial mediation (DE = 5.15*; IE = 1.10*)  % = 5.41    **CSA -> 4 -> AVH**  Partial mediation (DE = 5.15*; IE = 1.11*)  % = 5.93    Not significant effect of anxiety, obsessions and depression between CSA and AVH | 6 |
| **Mertens et al.**  **(2021, Spain)^10^** | 89  T1: 547  20.6  86  T2 (1.7 Years): 214  21.4  78  T3: 104  23.1  62.  T4 (4.4 years): 89  24.8  61.8 | Longitudinal | Subtype (EA)  ITEC | (1) Insecure attachment (RQ)  1.1 Dismissing  1.2 Preoccupied  1.3 Fearful  (2) Dissociation (DES-II) | Parallel & Serial Mediation Analysis | Yes  No | Paranoid Traits  (A) Self Reported  (SPQ)  (B) Interview Based (SCID-II) | Dismissing was not a significant mediator for both self-reported or interview based. Hence only occupied and fearful attachment were tested as parallel mediators.  **Parallel Mediations**  **EA -> 1.2 + 2 -> A**  (DE = 0.115*; IE 1.2 = 0.014*, IE 2 =0.042*, Total IE= 0.056*)  % 1.2= 8.1  % 2 = 24.6  Total IE % = 32.7  **EA -> 1.2 + 2 -> B**  (DE = 0.164*; IE 1.2 = 0.046*, IE 2 = 0.043*, Total IE= 0.088*)  % 1.2= 18.3  % 2 =17.1  Total IE % = 35  **EA -> 1.3 + 2 -> A**  (DE = 0.111*; IE 1.3 = 0.019*, IE 2 = 0.041*, Total IE = 0.060*)  % 1.3= 11.1  % 2 = 24  Total IE % = 35  **EA -> 1.3 + 2 -> B**  (DE = 0.182*; IE 1.3 = 0.024*, IE 2 =0.047*, Total IE= 0.070)  % 1.3= 9.6  % 2 = 18.7  Total IE % = 27.8  4 | 7 |
| **Metel *et al.* (2020, Poland)^19^** | 2684  26.37  62.3% | Cross sectional | Composite    TEC  CECA-Q | (1) Cognitive biases (DACOBS)    (2) Resilience (CD-RISC)    (3) Depressive symptoms (CESD-R) | Multiple mediation analysis | Yes    No | (1) Psychotic like experiences (PLE) (PQ) | **TEC/CECA-Q -> DACOBS -> CD-RISC CESD-R-> PLE**  Partial mediation (DE = 0.162*; IE = 0.163*)  % = 4 | 4 |
| **Moffa et al.**  **(2017, UK)^20^** | 2000 sample: N=8,580  2007 sample: N=7,403  Mean age: NR  % = NR | Cross Sectional | Composite Bullying  (Questionnaire) | (1) Worry (RV-CIS) (2) Sleep disturbance (RV-CIS) (3) Anxiety (RV-CIS) (4) Depression (RV-CIS) (5) Mood Instability (DSM-IV) (6) Persecutory Ideation | KHB Mediation Analysis | No  No | Hallucinations (PSQ) | Bullying had direct effects on worry, persecutory ideation, mood instability.  Depression, sleep and anxiety did not  mediate the link between bullying and persecutory ideation.  Bullying led to hallucinations indirectly, via persecutory ideation and depression. | 4 |
| **Murphy *et al.* (2015, UK)^105^** | 785  16.2  56.1% | Cross sectional | Composite    ELES | (1) Negative social comparisons (SCS)    (2) Trauma-related thoughts and beliefs (PTCI) | Moderated mediation analysis | Yes    Yes (recent victimization) | (1) Psychotic experiences (APSS) | **ELES-> PTCI-> APSS**  Partial mediation (non lonely patients) (DE = 0.049*; IE = 0.039*)  % = 44.3    **ELES -> PTCI -> APSS**  Total mediation (lonely patients) (DE = 0.032;  IE = 0.042*)  % = 56.7    No mediation by negative social comparisons either in lonely or non-lonely patients | 6 |
| **Ng et al. (2025, Hong Kong)^106^** | 143 473  Exposure GWAS: 143,473 (European ancestry UK Biobank participants)  NR  NR | Two-sample mendelian randomisation | Composite  Childhood Trauma Screener (GWAS proxy) | A. Cognitive   1. Functional executive  2. intelligence  Survey | Two-step Mendelian Randomisation (Mediation) | No  No | 1. schizophrenia | **CT → A1 → 1**  (DE ≈ 1.43*; IE = 0.34)  % = 19.26  **CT → A2 → 1**  (DE ≈ 1.70*; IE = 0.07)  % = 4.14  All other pathways were not included as they report socioeconomic mediators rather than psychological. | 5 |
| **Nonweiler et al.**  **(2023, Spain)^25^** | 1156  23.29  76.2% | Cross Sectional | Composite & Subtypes  CTQ Composite  CECA.A  (Parental A, Maternal A, Role reversal, Parental loss) | (1) Self Mentalizing (TMMS)  1.a Attention to emotions  1.b Emotional Clarity  (2) Other Mentalizing (Ment-S) | Parallel Multiple Mediation | Yes  No | (A) Schizotype (MMS)  A.1 Positive  (B) Positive PLEs (CAPE)  (C ) Paranoia (SPQ)  C.1 Suspiciousness  C.2 Ideas of reference | A.1) Positive Schizotypy  **CTQ-B-> 1.a + 1.b + 2 -> A.1**  (DE = 0.4511*; IE = 0.0846)  % = 15.7  **CTQ-B-> 1.a -> A.1**  (DE = 0.4511*; IE = 0.0456*)  % = 8.51  **CTQ-B-> 1.b-> A.1**  (DE = 0.4511*; IE = 0.0393*)  % = 7.34  **CTQ-B-> 2 -> A.1**  N.S  **CECA -> 1.a + 1.b + 2 -> A.1**  (DE = 0.2936*; IE = 0.0932*)  % = 24.1  **CECA Paternal -> 1.a -> A.1**  (DE = 0.2936*; IE = 0.743*)  % = 19.2  **CECA Paternal -> 1.b-> A.1**  N.S  **CECA Paternal -> 2 -> A.1**  N.S  **CECA Maternal -> 1.a + 1.b + 2 -> A.1**  (DE = 0.4934*; IE = 0.0915*)  % = 15.6  **CECA Maternal -> 1.a -> A.1**  (DE = 0.4934*; IE = 0.0390*)  % = 6.7  **CECA Maternal -> 1.b -> A.1**  DE = 0.4934*; IE = 0.0422*)  % = 8.6  **CECA Maternal -> 2 -> A.1**  N.S  **Role Reversal -> 1.a + 1.b + 2 -> A.1**  (DE = 0.0630*; IE = 0.0088*)  % = 12.2  **Role Reversal -> 1.a -> A.1**  (DE = 0.0630*; IE = 0.0069*)  % = 10  **Role Reversal -> 1.b-> A.1**  DE = 0.0630*; IE = -0.0010)  % = 1  **Role Reversal -> 2 -> A.1**  N.S  B) Positive PLEs (CAPE)  **CTQ-B-> 1.a + 1.b + 2 -> B**  (DE = 1.3257*; IE = 0.1069*)  % = 19.3  **CTQ-B-> 1.a ->B**  (DE = 1.3257*; IE = 0.1069*)  % = 6.88  **CTQ-B-> 1.b-> B**  (DE = 1.3257*; IE = 0.1242*)  % = 8  **CTQ-B -> 2 -> B**  N.S  **CECA Paternal -> 1.a + 1.b + 2 -> B**  (DE = 1.2034*; IE = 0.2350*)  % = 16.3  **CECA Paternal -> 1.a -> B**  (DE = 1.2034*; IE = 0.1954*)  % = 13.6  **CECA Paternal -> 1.b-> B**  N.S  **CECA Paternal -> 2 -> A.2**  N.S  **CECA Maternal -> 1.a + 1.b + 2 -> B**  (DE = 1.2644*; IE = 0.2365*)  % = 48.2  **CECA Maternal -> 1.a -> B**  (DE = 1.2644*; IE = 0.1020*)  % = 20  **CECA Maternal -> 1.b -> B**  (DE = 1.2644*; IE = 0.1281*)  % = 8.5  **CECA Maternal -> 2 -> B**  N.S  **Role Reversal -> 1.a + 1.b + 2 -> B**  (DE = 0.1438*; IE = 0.0196*)  % = 12  **Role Reversal -> 1.a-> B**  (DE = 0.1438*; IE = 0.0162*)  % = 10  **Role Reversal -> 1.b -> B**  **N.S**  **Role Reversal -> 2 -> B**  **N.S**  C.1) Suspiciousness  **CTQ-B-> 1.a + 1.b + 2 -> C.1**  (DE = 0.5575*; IE = 0.0960*)  % = 14.6  **CTQ-B -> 1.a -> C.1**  (DE = 0.5575*; IE = 0.0437*)  % = 6.69  **CTQ-B-> 1.b -> C.1**  (DE = 0.5575*; IE = 0.0524*)  % = 8.01  **CTQ-B-> 2 -> C.1**  N.S  **CECA Paternal -> 1.a + 1.b + 2 -> C.1**  (DE = 0.4743*; IE = 0.0915*)  % = 16.1  **CECA Paternal -> 1.a-> C.1**  (DE = 0.4743*; IE = 0.0682*)  % = 12.1  **CECA Paternal -> 1.b -> C.1**  N.S  **CECA Paternal -> 2 -> C.1**  N.S  **CECA Maternal -> 1.a + 1.b + 2 -> C.1**  (DE = 0.5377*; IE = 0.0996*)  % = 15.6  **CECA Maternal -> 1.a ->C.1**  (DE = 0.5377*; IE = 0.0389*)  % = 6.1  **CECA Maternal -> 1.b-> C.1**  (DE = 0.5377*; IE = 0.0581*)  % = 9.11  **CECA Maternal -> 2 -> C.1**  N.S  **Role Reversal -> 1.a + 1.b + 2 -> C.1**  (DE = 0.0458*; IE = 0.0063*)  % = 12.1  **Role Reversal -> 1.a -> C.1**  (DE = 0.0458*; IE = 0.0069*)  % = 13.2  **Role Reversal -> 1.b -> C.1**  (DE = 0.0458*; IE = -0.0013*)  % = 2.4  **Role Reversal -> 2 -> C.1**  **N.S**  C.2) Ideas reference  **CTQ-B -> 1.a + 1.b + 2 -> C.2**  (DE = 0.2520*; IE = 0.1108*)  % = 30.5  **CTQ-B-> 1.a -> C.2**  (DE = 0.2520*; IE = 0.0550*)  % = 15.16  **CTQ-B-> 1.b -> C.2**  N.S  **CTQ-B-> 2 -> C.2**  N.S  **CECA Paternal -> 1.a + 1.b + 2 -> C.2**  (DE = 0.3154*; IE = 0.1154*)  % = 26.8  **CECA Paternal -> 1.a -> C.2**  (DE = 0.3154*; IE = 0.0899*)  % = 20.9  **CECA Paternal -> 1.b -> C.2**  N.S  **CECA Paternal -> 2 -> C.2**  N.S  **CECA Maternal -> 1.a + 1.b + 2 -> C.1**  (DE = 0.3761*; IE = 0.1155*)  % = 23.5  **CECA Maternal -> 1.a ->C.1**  (DE = 0.3761*; IE = 0.0476*)  % = 6.1  **CECA Maternal -> 1.b -> C.1**  (DE = 0.3761*; IE = 0.0624*)  % = 12.6  **Role Reversal -> 1.a + 1.b + 2 -> C.1**  (DE = 0.0344*; IE = 0.0079*)  % = 18.6  **Role Reversal -> 1.a -> C.1**  (DE = 0.0344*; IE = 0.0080*)  % = 19  **Role Reversal-> 1.b -> C.1**  **N. S**  **Role Reversal -> 2 -> C.1**  **N.S** | 5 |
| **O'Neill et al.**  **(2021, UK, USA & Australia)^107^** | *n* Total = 269  *n* UK = 137  32.89  87.5%  *n* USA = 127  32.21  94.4%  *n* Australia = 35  33.57  82.8% | Cross Sectional | Sexual Abuse  Subtype  SAQ | (1) Dissociation  (DES)  1. a.- absorption  1.b - depersonalisation  1.c- amnesia | Mediation Analysis | Yes  Yes (age, education, employment) | Psychotic Like Experiences (APSS) | **SAQ -> 1.a -> PLEs**  (DE = 0.10* ; IE = 0.057)  N.S  **SAQ -> 1.b-> PLEs**  (DE = 0.10* ; IE = 0.249*)  % = 71.4  **SAQ -> 1.c -> PLEs**  (DE = 0.10* ; IE = 0.022)  N.S | 5 |
| **Paetzold**  **(2023, Belgium)^108^** | 1682  13.4  63 | Cross Sectional | Composite & subtypes  JVQ  Bullying Prevalence (BP)  Interview  Bullying Severity (BS)  Interview | Threat Anticipation  (Availability Test) | Mediation Analysis | Yes  Yes (age, gender, ethnicity, cognitive functioning) | (A) Prodromal Symptoms  (PQ-16)  A.1 Anomalous experiences | **JVQ -> Threat Anticipation -> A.1**  (DE = 0.26, IE = 0.05*)  % = 16.1  **BP -> Threat Anticipation -> A.1**  (DE = 0.20, IE = 0.04*)  % = 16.7  **BS -> Threat Anticipation -> A.1**  (DE = 0.24, IE = 0.04*)  % = 14.9 | 6 |
| **Perona-Garcelán *et al.* (2014, Spain)^109^** | 318  21.41  78.9% | Cross sectional | Composite    Trauma Questionnaire (TQ) | (1) Dissociation  1.a - Tellegen Absorption Scale (TAS)  1.b - Depersonalization (CDS)    (2) Southampton Mindfulness Questionnaire (SMQ) | Multiple mediation analysis | Yes    No | (1) Hallucination proneness (HP) (LSHS-R) | **TQ -> Absorption-> Scale HP**  Total mediation (DE = 0.12; IE = 0.38*)  % = 82.6    **TQ-> Depersonalisation->HP**  Total mediation (DE = 0.12; IE = 0.16*)  % = 34.78 (together)    No mediation by mindfulness | 5 |
| **Pinto-Gouveia *et al.* (2014, Portugal)^110^** | 255  36.36  68.2% | Cross sectional | Composite (including threat, submissiveness and feeling unvalued)    ELES | (1) External shame (OAS) | Path analysis | Yes    Yes (confounders not specified) | (1) Paranoia (GPS) | **ELES OAS GPS**  Partial mediation (DE = 0.12*; IE = 0.0608*)  % = 33.6 | 6 |
| **Qiao et al.**  **(2023, Belgium)^21^** | 865  15.50  67% | Cross Sectional | Composite  JVQ | (1) Stress (ESM)  (2) Negative affect (ESM)  (3) Loneliness (ESM)  (4) General Psychopathology (BSI)  (5) Attachment Insecurity (IPPA-R)  (6) Threat Anticipation (AT) | Network Analysis | Yes  No | Psychotic Experiences  (PQ-16) | Depression, anxiety, negative affect, loneliness, and threat anticipation displayed highest centrality in the formation of the network.  Connective role of anxiety, hostility, and somatization in the shortest paths linking childhood adversity and PEs. | 4 |
| **Rossi et al.**  **(2023, Italy) ^111^** | 1010  18.7  49.31 | Cross Sectional | Composite  ITEM | (1) Resilience  (RSA-11)  1.1 Personal  1.2 Interpersonal  (2) Attachment  (RQ)  2.1 Preoccupied  2.2 Fearful  2.3 Dismissing | Parallel mediation | Yes  No | Psychotic Like Experiences  (iPQ-16) | **ITEM -> 1.1 -> iPQ-16**  (DE = 0.30*; IE = 0.076*)  % = 20.1  Parallel Mediation  **ITEM -> 2.1 + 2.2 + 2.3 -> iPQ-16**  (DE = 0.23; IE 2.1= 0.085*, IE 2.2 = 0.083*, IE 2.3 = 0.013*)  % 2.1 = 19.2  % 2.2 = 18.7  % 2.3 = 2.9 | 6 |
| **Rössler *et al.* (2016, Switzerland) ^112^** | 820  31.36  55.8% | Cross sectional | Composite (CTQ) and specific (EA, PN, EN)    CTQ | (1) Stress sensitivity (SS) (PSS, PANAS-N, SSCS) | Bivariate and mediated multinomial logistic regression path models | Yes    Yes (trauma types, education) | (1) Subclinical psychotic experience (SPE)  (SIAPA, SPQ-B German version, PARA, and STS and SNS subscales from SCL-90-R) | **CTQ -> SS -> SPE**  Partial mediation (DE = 0.44*; IE = 0.50*)  % = 43.05    **EA -> SS -> SPE**  Total mediation (DE = 0.47; IE = 0.578*)  % = 46.6    **EN -> SS -> SPE**  Total mediation (DE = -0.31; IE = 0.44*)  % = 53.38    **PN-> SS-> SPE**  Total mediation (DE = 0.27; IE = 0.28*)  % = 28.71 | 6 |
| **Saladino et al. (2025, Italy)^113^** | 1894  15.46  %50 | Longitudinal SEM | Subtype (emotional neglect)  CTQ-SF emotional neglect subscale | A. Depression (DASS-21) | Three-wave SEM | Yes  Maternal education, paternal education, gender | 1.Psychoticism  (PID-5-BF) | **EN → A → 1**  Non significant | 5 |
| **Sheinbaum et al.**  **(2015, Spain)^22^** | 214  21.4  78% | Cross Sectional | Subtypes (antipathy, role reversal)  CECA | (A) Attachment style (ASI)   A.1 Enmeshed  A.2 Fearful  A.3 Angry- dismissive  A.4 Withdrawn | Parallel multiple Mediation | Yes  Yes (depressive symptoms) | (1) Schizophrenia spectrum phenomenology   (CAARMS)  1.1 Positive symptoms  (2) Paranoid PD Ratings (SCID)  (3) Schizotypal PD Ratings  (SCID) | **Antipathy -> A.1 + A.2 + A.3 + A.4 -> 1.1**  (DE = 0.5*; IE A.3 = 0.13*)  % A.3 = 19  All other mediators were N.S  **Antipathy -> A.1 + A.2 + A.3 + A.4 -> 2**  (DE = 0.21; IE = 0.35*; IE A.1 = 0.13*; A.3 = 0.21*)  % total = 62  % A.1 = 23  % A.3 = 38  All other mediators were N.S  **Antipathy -> A.1 + A.2 + A.3 + A.4 -> 3**  (DE = 0.21; IE = 0.25*; IE A.1 = 0.14*; A.3 = 0.13*)  % total = 55  % A.1 = 30  % A.3 = 27  All other mediators were N.S  **Role Reversal -> A.1 + A.2 + A.3 + A.4 -> 2**  (DE = 0.18; IE = 0.2*; IE A.1 = 0.11)  % total = 52  % A.1 = 29  All other mediators were N.S  **Role Reversal -> A.1 + A.2 + A.3 + A.4 -> 3**  (DE = 0.26*; IE = 0.17*; IE A.1 = 0.12*  % total = 39  % A.1 = 28  All other mediators were N.S | 6 |
| **Sheinbaum et *al.* (2014, Spain)^114^** | 546  20.6  82.3% | Cross sectional | Composite factor of physical and emotional trauma (P/E)    CTQ | (1) Attachment style (RQ)  1.a - Dismissing  1.b - Preoccupied  1.c - Fearful | Parallel multiple mediation analyses | Yes    No | (1) PLEs (CAPE)    (2) Paranoid beliefs (SPQ)    (3) Schizotypy (WSS) | **P/E -> Fearful -> Schizotypy**  Partial mediation (DE = 0.140*; IE total = 0.028*; IE fearful = 0.010*)  % total = 16.6  % fearful = 5.9    **P/E -> Fearful -> Suspiciousness**  Partial mediation (DE = 0.420*; IE total = 0.093*; IE fearful = 0.056*)  % total = 18  % fearful = 10.8    **P/E -> Fearful -> PLE**  Partial mediation (DE = 0.822*; IE total = 0.142*; IE fearful = 0.063*)  % total = 14.7  % fearful = 6.5    None of the other mediators were significant | 5 |
| **Sheinbaum**  **(2020, Spain)^23^** | 169  28  80.5 | Longitudinal | Composite factor of physical and emotional maltreatment (P/E)  CTQ | (1) Preoccupied attachment (RQ)  (2) Dismissing attachment (RQ)  (3) Fearful attachment (RQ)  (4) Disorganized attachment (ADA) | Parallel Multiple Mediation Analysis | Yes  No | (A) Paranoid Beliefs (SPQ)  (B) Positive Schizotypy (WSS) | **P/E -> 1,2,3 -> A**  (DE = 0.48*, IE total = 1.104*, IE fearful = 0.078*)  % total = 17.9  % fearful = 13.4  **P/E -> 1,2,4 -> A**  (DE = 0.459*, IE total = 0.125*, IE disorganized = 0.101*)  % total = 21.4  % Disorganized = 17.3  **P/E -> 1,2,3,4 -> A**  (DE = 0.419*, IE total = 0.165*, IE disorganized = 0.088*)  % total = 28.6  % Disorganized = 15  No other mediators were significant. | 8 |
| **Shevlin *et al.* (2015, UK)^115^** | 7403  51.12  56.8% | Cross sectional | Specific CSA and/or CPA and CSA + CPA composite scores    Questionnaire | (1) Loneliness (SFQ) | Mediation analysis | No    Yes (age, gender, education, ethnicity, cannabis use and adult CSA and CPA) | (1) Psychosis diagnosis (SCAN) | **CSA + CPA -> Loneliness -> Psychosis**  Partial mediation (DE = N/A; IE = 0.722*)  % = N/A    No mediation of the CPA, CSA separately | 5 |
| **Sitko *et al.* (2014, UK)^116^** | 5877  34.5  50.5% | Cross sectional | History module  Specific (Rape, sexual molestatin, PA, Physical assault/attack)    UM-CIDI - Life Event | (1) Attachment style (AAQ)  1.a - Secure (reversed)  1.b - Avoidance  1.c - Anxious | Regression based approach | Yes    Yes (age and gender) | (1) Lifetime psychotic symptoms (LPS) (Beliefs and Experiences module of the  UM-CIDI)  1.A - Paranoia  1.B - Hallucinations | **Neglect -> 1.c + 1.b -> 1.A**  Total mediation (DE = 0.047; IE = N/A)  % both = 100    **Rape -> 1.c -> 1.B**  Partial mediation (DE = 0.088*; IE = N/A)  % = 3.0 | 6 |
| **Strelchuk et al.**  **(2022, UK)^17^** | Adolescents  2952  NR  57.8  Adults  2492  NR  NR  2952 | Longitudinal | Composite  Trauma Interview  TP 1: Age 0-14  TP2: Age 0-17 | (1) PTSD age 15 (DAWBA)  (2) PTSD age 24 (PCL-5) | Mediation analysis | Yes  Yes (Sex, IQ, Social class, Family History Mental health | Psychotic Experiences (PLIKSi)  (A) Adolescent PEs  (B) Adult PEs | **CT Age 0-14 -> 1 -> A**  (DE = 1.40*; IE = 1.05*)  % = 14  **CT Age 0-17 -> 2 -> B**  (DE = 1.51*; IE = 1.03*)  % = 8 | 8 |
| **Van Nierop *et al.* (2014, Netherland)^117^** | 6646  44  55% | Cross sectional | Composite (CT)    NEMESIS-1 | (1) Social defeat (SD)  (NEMESIS questionnaire)    (2) Affective dysregulation (AD)  (NEMESIS questionnaire) | Multiple mediation  analyses | Yes    Yes (CT, age, gender, cannabis use and affective dysregulation) | (1) Psychosis diagnosis (PD) DSM-IV (SCID-I)    (2) Extended psychosis phenotype (EPP) (NEMESIS interview) | **CT -> AD-> EPP**  Partial mediation (DE = N/A; IE = 0.04*)  % = 49.7    **CT -> SD -> PD**  Partial mediation (DE = N/A; IE = 0.04*)  % = 86.6    No significant mediation of SD on the link between CT and EPP and of AD on the link between CT and PD | 6 |
| **Wolke *et al.* (2014, UK)^118^** | 4720  17.1  56.5% | Prospective, assessed for adversity at 8 and 11 yoa, and for symptoms at 12.9 and 18 yoa | Two composite scores peer victimization (PV) (child-reported and mother-reported bullying)    BFIS  SDQ | (1) Depression symptoms at age 12, 13, 14 yoa (SQFM) | Path analysis | No    Yes (gender, any DSM-IV Axis I diagnosis, IQ and internalizing / externalizing behavior) | (1) Psychotic experiences (PE) at age18 yoa (PLIKSi) | **PV-> Depression (12.9) -> PE (18)**  Partial mediation (DE = 0.13*; IE child reported = 0.03*; IE mother reported = 0.02*)  % child reported = 18.75  % mother reported = 13.3 | 7 |
| **Yamasaki *et al.* (2016, Japan)^119^** | 4277  9.8  46.9% | Cross sectional | Composite score (peer victimization (PV))    OVBQ | (1) Dissociation (CBCL)    (2) Depressive symptoms (SMFQ)    (3) External locus of control (shortened  version of the CNSIE) | SEM | No    No | (1) Hallucinations (CBCL) | **PV -> Dissociation -> CBCL**  Total Mediation (DE = 0.02; IE = 0.038*  % = 95    No mediation by depression or external locus control | 4 |
| **Zhang et al.**  **(2022, China) ^120^** | 5873  19.36  57.3 | Cross Sectional | Composite & Subtypes  CTQ (N,A) | (1) Wisdom (SD-WISE) | Mediation Analysis | Yes  Yes (sex, age) | Psychic Experiences (  CAPE) | **CTQ -> SD-WISE -> CAPE**  (DE = 0.1059*; IE = 0.05855*)  % = 22  **N -> SD-WISE -> CAPE**  (DE = 0.1162*; IE = 0.061* )  % = 43  **A -> SD-WISE -> CAPE**  (DE = 0.1347*; IE = 0.029*)  % = 18 | 5 |

AAQ: Acceptance and Action Questionnaire; ACE: Adverse Childhood Experiences; ACE-IQ: Adverse Childhood Experiences International Questionnaire; ADA: Adult Disorganized Attachment scale; APMS: Adult Psychiatric Morbidity Survey; APSS: Adolescent Psychotic-Like Symptom Screener; ASI: Attachment Style Interview; AST: Ambiguous situation task; AT: Availability Test; BAI: Beck Anxiety Inventory; BCSS: Brief Core Schema Scale; BDI: Beck Depression Inventory; BFIS: Bullying and Friendship Interview Schedule; BIP2: Bipolar II Scale; BSI: Brief Symptom Inventory; CAPE: Community Assessment of Psychic Experiences; CAPE-P: The Positive Symptoms Frequency subscales of the Community Assessment of Psychic Experiences; CAPE-42: Community Assessment of Psychic Experience-42; CAPS: Clinician-Administered PTDS Scale; CAARMS: Comprehensive Assessment of At Risk Mental States; CATS: Child Abuse and Trauma Scale; CBCL: Child Behavior Checklist; CD-RISC: Connor-Davidson Resilience Scale; CDS: Cambridge Depersonalization Scale; CECA: Childhood Experiences of Care and Abuse; CECA.Q: Childhood Experience of Care and Abuse Questionnaire; CES: Curious Experiences Survey; CESD-R: Center for Epidemiologic Studies-Depression Scale; CISI: Composite International Diagnostic Interview; CIS-R: Clinical Interview Schedule-Revised; CNSIE: Childhood Nowicki–Strickland Internal–External; CSA: Childhood Sexual Abuse; CT: Childhood trauma; CTQ: Childhood Trauma Questionnaire; CTQ-SF: Childhood Trauma Questionnaire-Short Form; DACOBS-18: Davos Assessment of Cognitive Biases Scale; DAWBA: Development and Well-Being Assessment; DES-II: Dissociative Experience Scale; DES-T: Dissociative Experiences Scale; DPS: Dissociative Processes Scale; DTD: Draws-to-decision; EA: Emotional Abuse; ECR-R: Experiences in Close Relationships-Revised Questionnaire; EN: Emotional Neglect; EPQ-S: Eysenck personality questionnaire (short form revised version); ERSQ: Emotion Regulation Skills Questionnaire; ESM: Experience Sampling Method; ESS: Experience of Shame Scale; GAD-7: Generalized Anxiety Disorder Scale; GPS: General Paranoia Scale; GPTS: Green et al. Paranoid Thought Scales; HADS: Hospital Anxiety and Depression Scale; IES-R: Impact of Event Scale-Revised; IPASE: Inventory of Psychotic-Like Anomalous Self-Experiences; IPAAR: Inventory of Parent and Peer Attachment Revised; IPSM: Interpersonal sensitivity scale; ISI: Insomnia Severity Index; IPQ-16: 16-item Prodromal Questionnaire; ITEC: Interview for Traumatic Events in Childhood; ITEM: International Trauma Exposure Measure; JVQ: Juvenile Victimisation Questionnaire; KHB: Karlson–Holm–Breen; LSHS-R: Launay Slade Hallucinations Scale-Revised; MAP-SR: Motivation and Pleasure Scale - Self-Report; MDI: Multiscale Dissociation Inventory; MENT-S: Mentalization Scale; MSS: Multidimensional Schizotypy Scale; NA: Not available; NEMESIS: Netherlands Mental Health Survey and Incidence Study; NSIE: Nowicki–Strickland Internal–ExternaL; OAS: Other As Shamer Scale; OVBQ: Olweus Bully/Victims Questionnaire; PA: Physical Abuse; PAGE-R: Exceptional Experiences Questionnaire-Revised; PANAS-N: Negative Affect Subscale of the Positive and Negative Affect Scale; PARA: Paranoia Checklist; PCL-5: Post-traumatic Stress Disorder Checklist for DSM-5; PDEQ: Peritraumatic Dissociative Experiences Questionnaire; PDI: Peters et al. Delusions Inventory; PDI-IV: Personality Disorder Interview-IV; PHQ-9: Patient Health Questionnaire; PHS: Piers-Harris scale; PLIKSi: Semi-structured Psychosis Interview; PLS-SEM: Partial least squares structural equation model; PN: Physical Neglect; PQ: Prodromal Questionnaire; PQ-16: Prodromal Questionnaire-16; PS: Psychosis Screener; PSQ: Psychosis Screening Questionnaire; PSS: Perceived Stress Scale; PTCI: Posttraumatic Cognitions Inventory; RBQ: Retrospective Bullying Questionnaire; RCBI-II: Second Revision of the revised Cyber Bullying Inventory; REF: Ideas of Reference; RHS: Revised Hallucination Scale; RI-E: Rotter I-E Scale; RQ: Relationship Questionnaire; RSA-11: Resilience Scale for Adults; RSES: Rosenberg Self-Esteem Scale; SA: Sexual Abuse; SAQ: Sexual Abuse Questionnaire; SCAN: Schedules for Clinical Assessment in Neuropsychiatry; SCID-II: Structured Clinical Interview for DSM-IV Personality Disorders; SCID-D-R: Structured Clinical Interview for DSM-IV Dissociative Disorders Revised; SD-WISE: San Diego Wisdom Scale; SCL-90-R: Symptom Checklist-90-R; SCS: Social Comparison Scale; SDQ: Strengths and Difficulties Questionnaire; SDQ-5: 5-item Somatoform Dissociation Questionnaire; SEM: Structural Equation Modeling; SES: Sexual Experiences Survey; SELCoH: South East London Community Health Study; SFQ: Social Functioning Questionnaire; Shut-D: Shutdown dissociation scale; SIAPA: Structured Interview for Assessing Perceptual Anomalies; SIPS: Structured Interview for Psychosis-Risk Syndromes; SMFQ: Short Mood and Feelings Questionnaire; SNS: Schizophrenia Nuclear Symptom Scale; SPQ: Schizotypal Personality Questionnaire; SPQ: Self-Reported Paranoid Traits; SPQ-B: Schizotypal Personality Questionnaire-Brief; SQFM: Short Mood and Feelings Questionnaire; SQ-SF: Young Schema Questionnaire Short Form; SSCS: Screening Scale for Chronic Stress; STR: Subjective Experiences of Early Life Trauma; STS: Schizotypal Signs Scale; TEC: Traumatic Experience Checklist; TMMS: Trait Meta-Mood Scale-24; TSQ: Trauma Screening Questionnaire; UCLA: UCLA Loneliness Scale; UM-CIDI - Life Event: modified version of the Composite International Diagnostic Interview; WSS: Wisconsin Schizotypy Scales; YSQ-SF: Young Schema Questionnaire-Short Form.

### Table S7 . Overview of Biological Mediators examined in non-clinical studies included in this review

| **Authors**  **Country** | **Sample**  **Mean Age**  **% female** | **Design** | **Measures of childhood adversity** | **Mediator(s)** | **Analysis** | **Bootstrap**  **(yes / no) / confounders (yes / no)** | **Psychosis** | **Main findings**  **Pathway**  **Total / partial mediation**  **Direct Effect (DE)**  **Indirect effect (IE)**  **% total effect mediated** | **Quality Score** |
| --- | --- | --- | --- | --- | --- | --- | --- | --- | --- |
| **Dahoun et al. (2019, UK)^121^** | 24 (GP)  23.4  45.8% | Cross sectional | Composite  CTQ | a. Dopamine release | Mediation analysis | Yes  Yes (sex, nicotine,and alcohol, cannabis, morphin, stimulants) | 1. 1. Dexamph-induced positive symptoms (PANSS) | **CT -> a -> 1**  Null Mediation | 6 |
| **Ng et al. (2025, Hong Kong)^106^** | 143 473  Exposure GWAS: 143,473 (European ancestry UK Biobank participants)  NR  NR | Two-sample mendelian randomisation | Composite  Childhood Trauma Screener (GWAS proxy) | IL6  IL6R  BMI  Cortisol | Two-step Mendelian Randomisation (Mediation) | No  No | 2. schizophrenia | All pathways were not reported because they were not significant. | 5 |
| **Okada et al.**  **(2024, Japan)^122^** | Time 1  219  11.5  47.9  Time 2  211  13.6  47.4 | Longitudinal | Composite Bullying victimisation (BV) | Glutamate-glutamine (Glx) Levels | Partial least squares SEM | Yes    Yes (age, sex, SES, IQ, Help seeking intention) | Subclinical Psychotic Experiences (DISC-C)    Timepoint 1  Timepoint 2 | **BV -> GLx -> DISC-C**  (Suggested pathway of interest)    No indirect effects were observed in this pathway. | 6 |
| **Smigielski et al.**  **(2021, Switzerland)^123^** | 24  27.38  0 | Cross Sectional | Composite  CTQ | D2/3R availability | Mediation analysis | Yes  No | Odd beliefs (PAGE-R) | **CTQ -> D3/3R availability -> Odd beliefs**  (DE = 0.192; IE = 0.131*)  % = 41 | 5 |

CTQ: Childhood Trauma Questionnaire; D2/3R: Postsynaptic D2/3 Receptor Availability; DISC-C: Diagnostic Interview Schedule for Children; PANSS: Positive and Negative Syndrome Scale; SEM: Structural Equation Modeling

### Composite Dissociation Meta-SEM results

Pooled Correlation Matrix (Random−Effects Model)

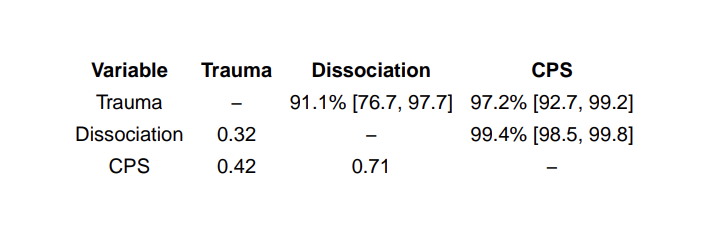

Below diagonal: pooled random−effects correlations. Above diagonal: I² with 95% likelihood−based CI (% of variance due to between−study heterogeneity).

Path Coefficients and Mediation Effects

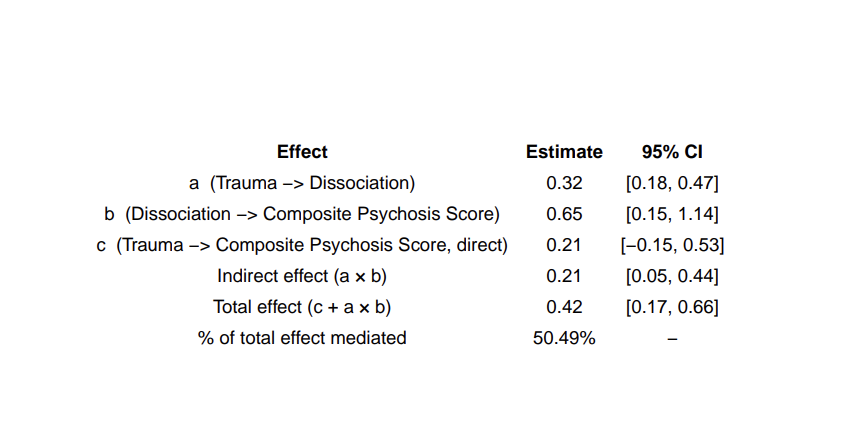

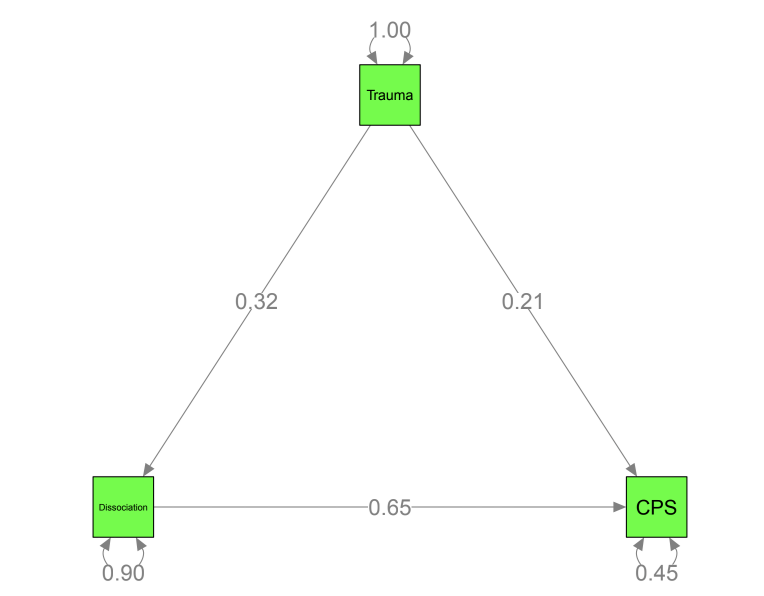

Per−Study Indirect Effects: Trauma −> Dissociation −> CPS

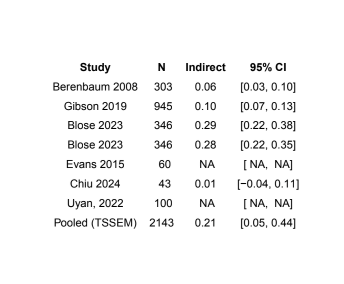

Leave−One−Out Sensitivity Analysis: Results Table

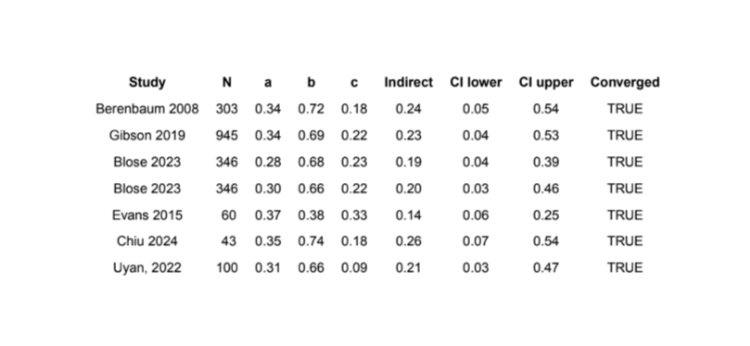

Leave−One−Out Sensitivity Analysis
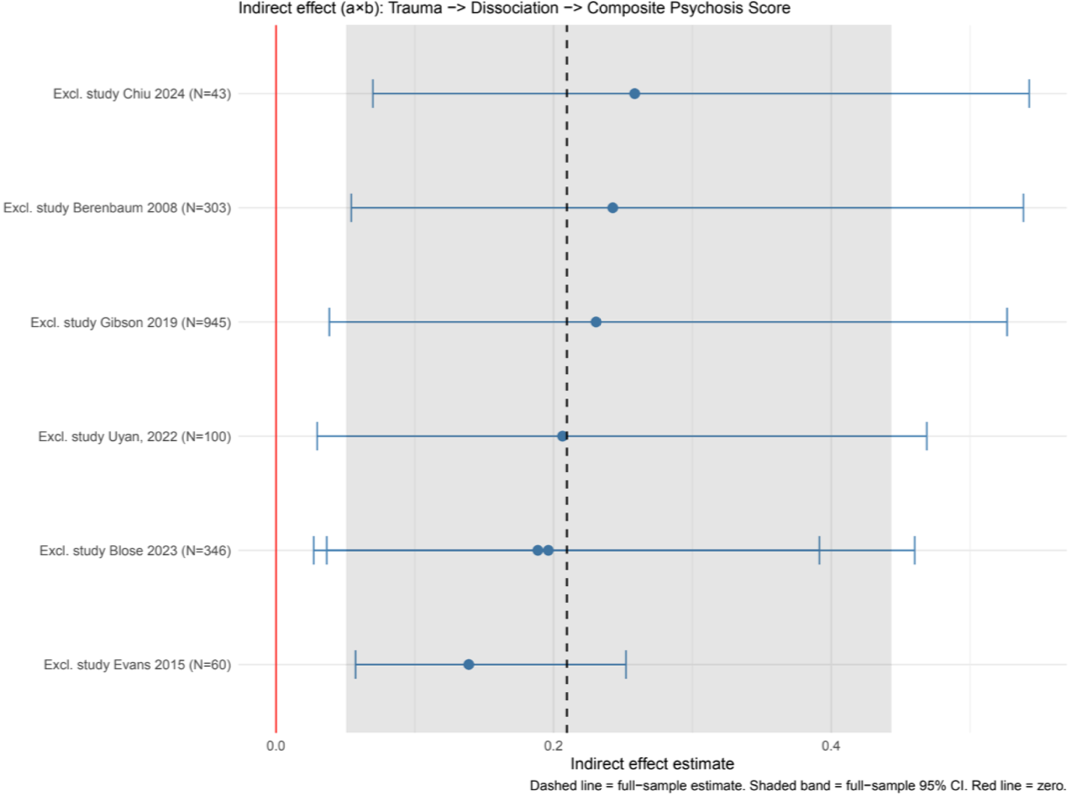

### Hallucinations Dissociation Meta-SEM results

Pooled Correlation Matrix (Random−Effects Model)

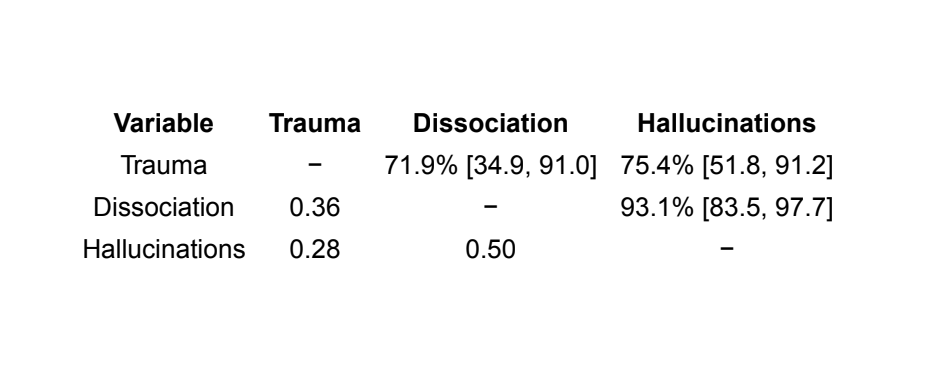

Below diagonal: pooled random−effects correlations. Above diagonal: I² with 95% likelihood−based CI (% of variance due to between−study heterogeneity).

Path Coefficients and Mediation Effects

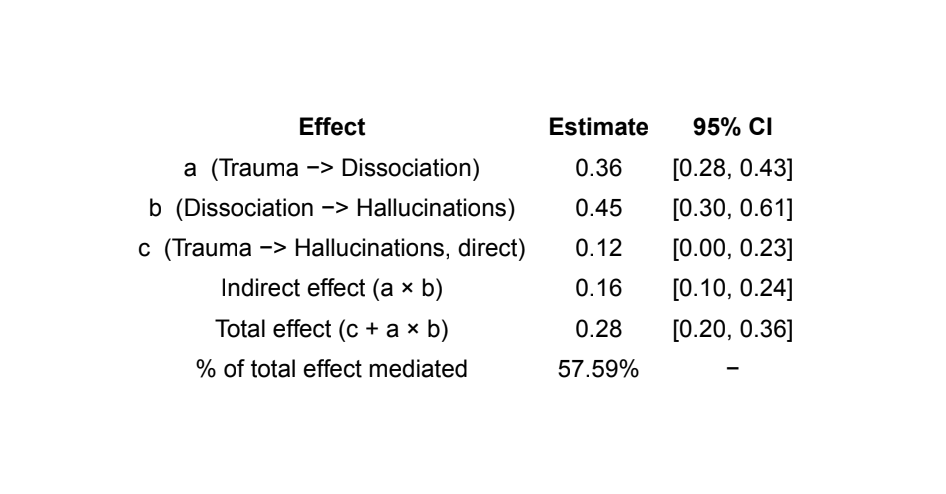

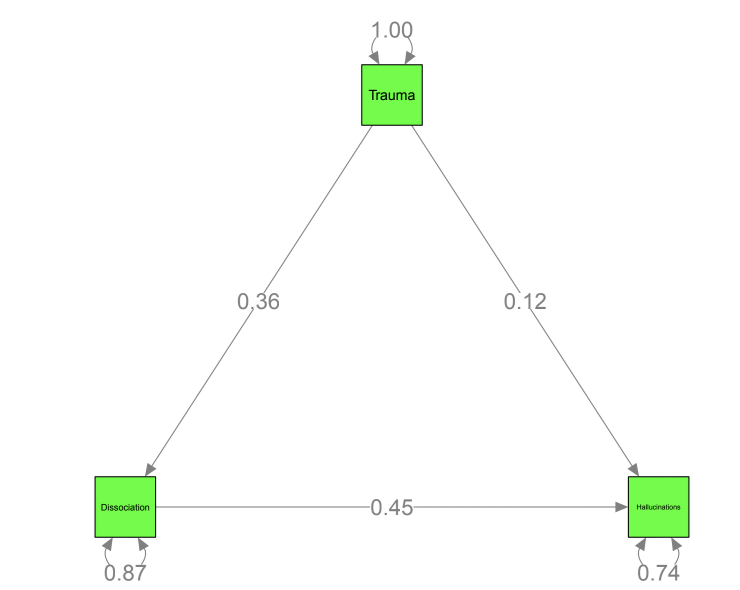

Per−Study Indirect Effects: Trauma −> Dissociation −> Hallucinations

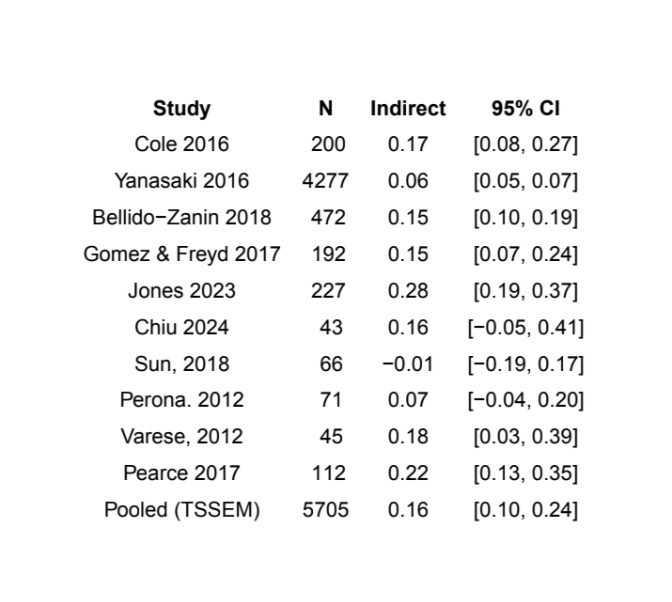

Leave−One−Out Sensitivity Analysis: Results Table

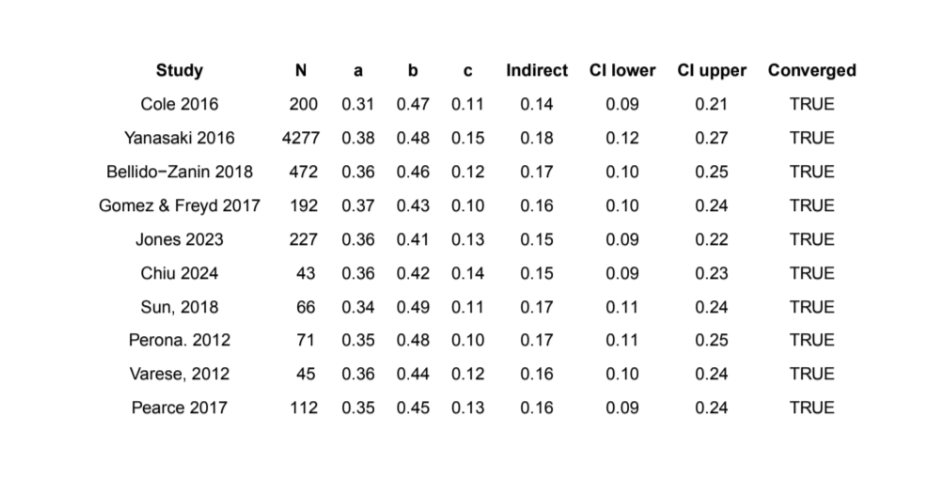

Leave−One−Out Sensitivity Analysis

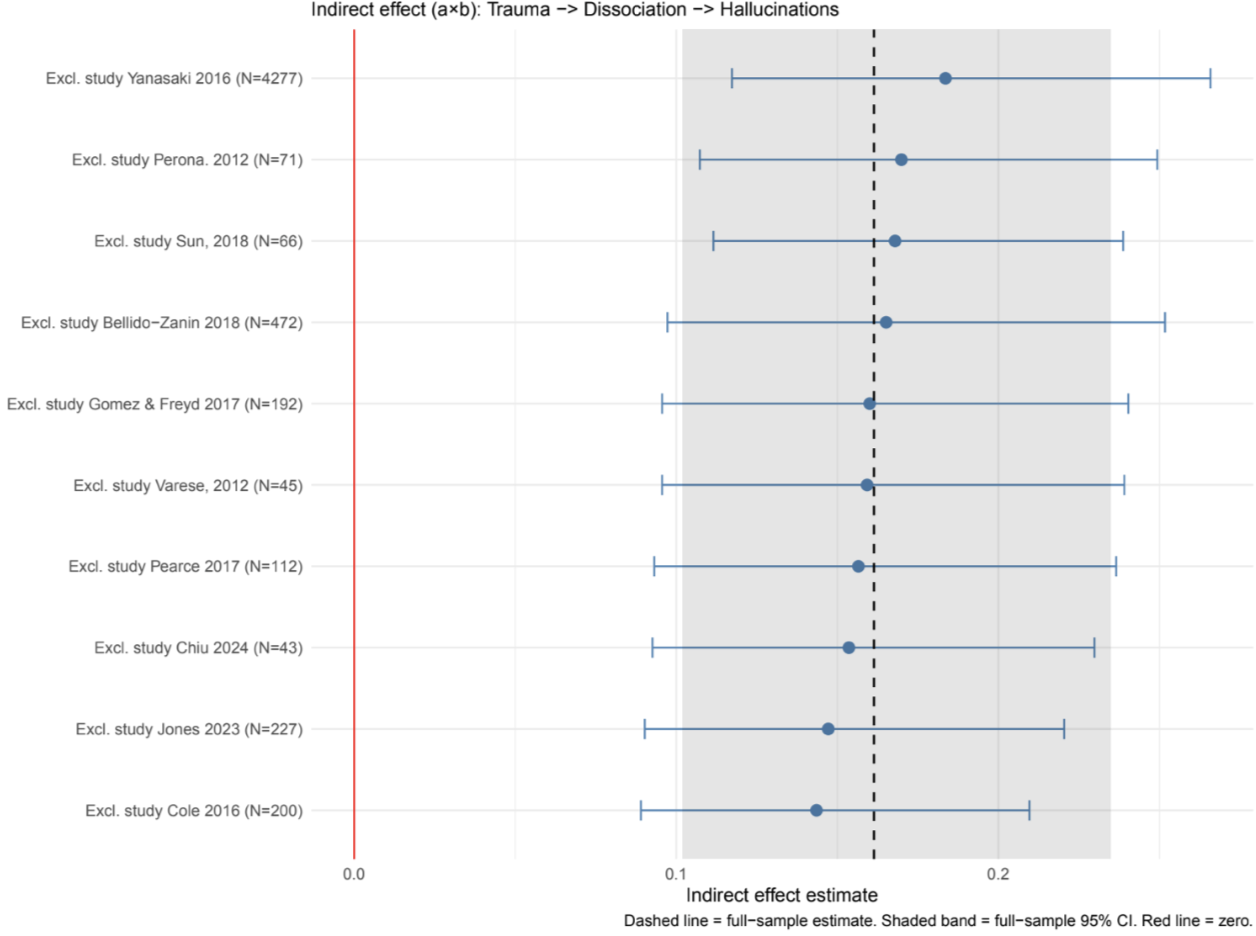

### Delusions Dissociation Meta-SEM results

Pooled Correlation Matrix (Random−Effects Model)

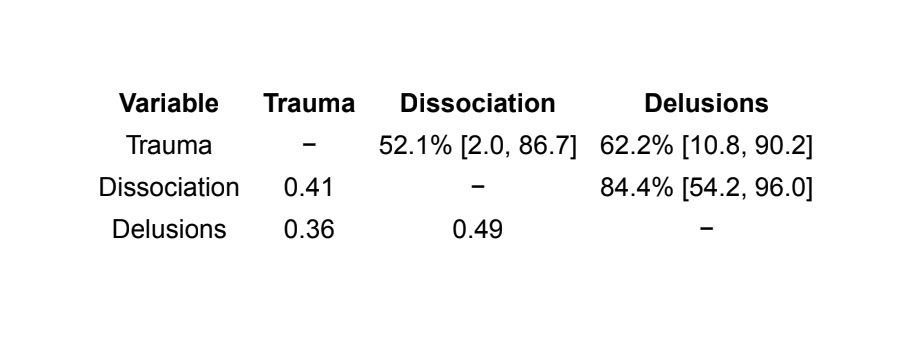

Below diagonal: pooled random−effects correlations. Above diagonal: I² with 95% likelihood−based CI (% of variance due to between−study heterogeneity).

Path Coefficients and Mediation Effects

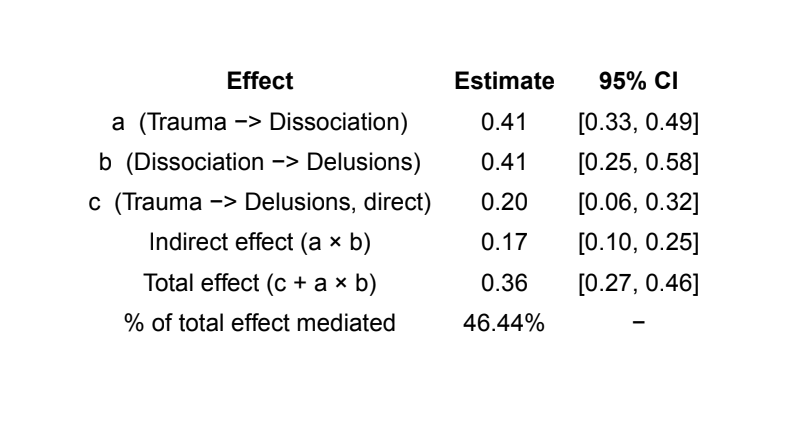

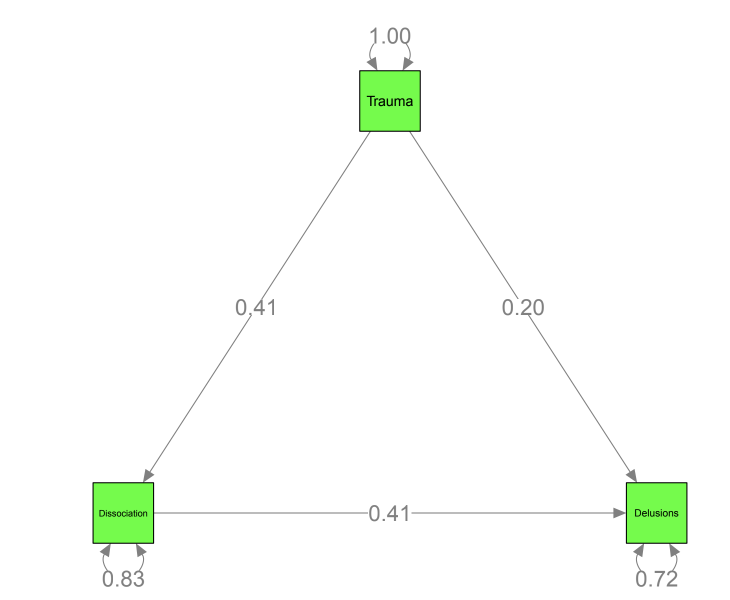

Per−Study Indirect Effects: Trauma −> Dissociation −> Delusions

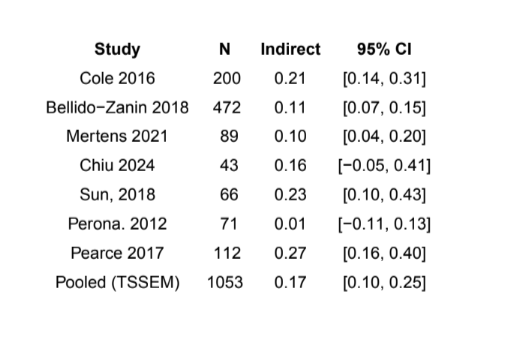

Leave−One−Out Sensitivity Analysis: Results Table

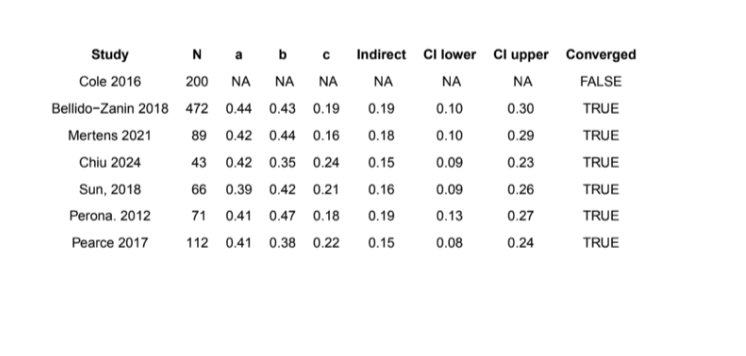

Leave−One−Out Sensitivity Analysis

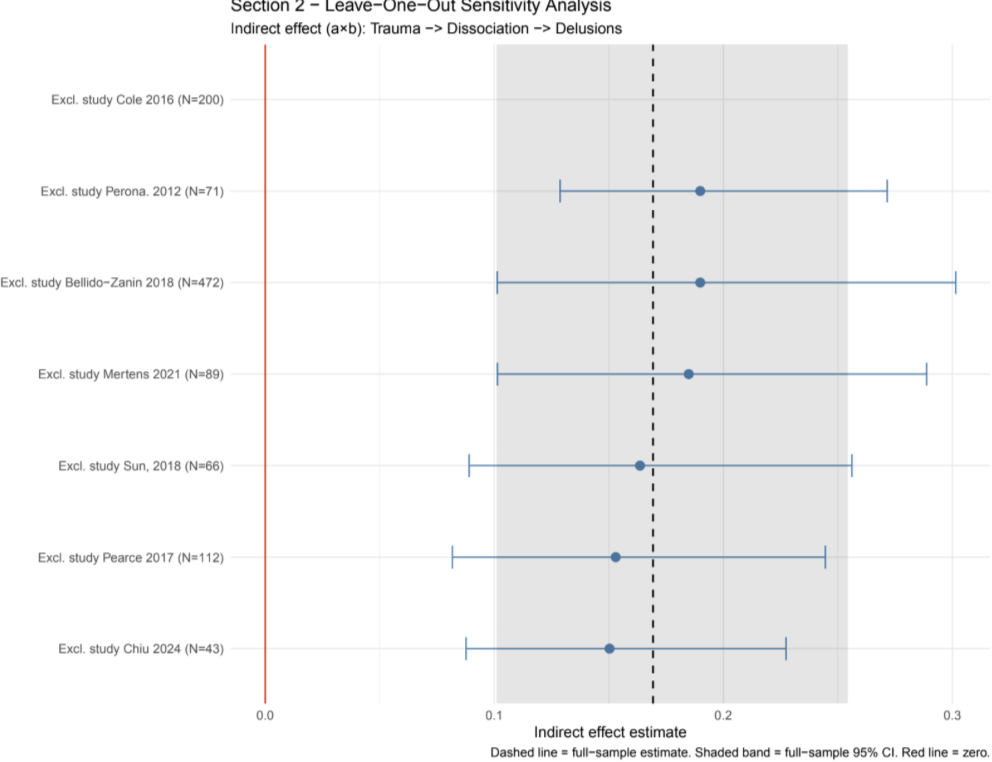

### Composite Negative Schema Meta-SEM results

Pooled Correlation Matrix (Random−Effects Model)

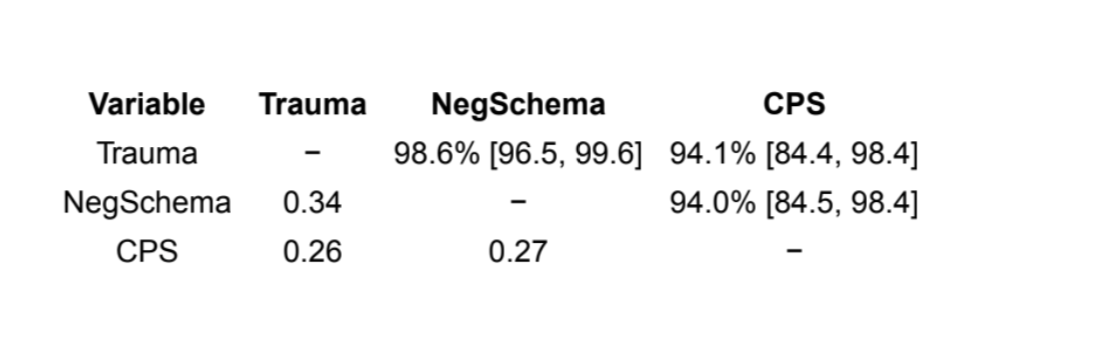

Below diagonal: pooled random−effects correlations. Above diagonal: I² with 95% likelihood−based CI (% of variance due to between−study heterogeneity).

Path Coefficients and Mediation Effects

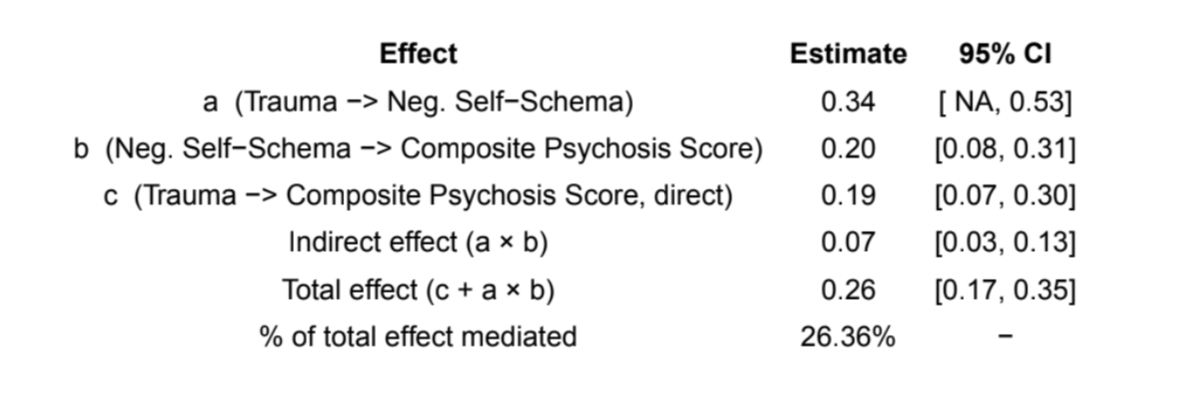

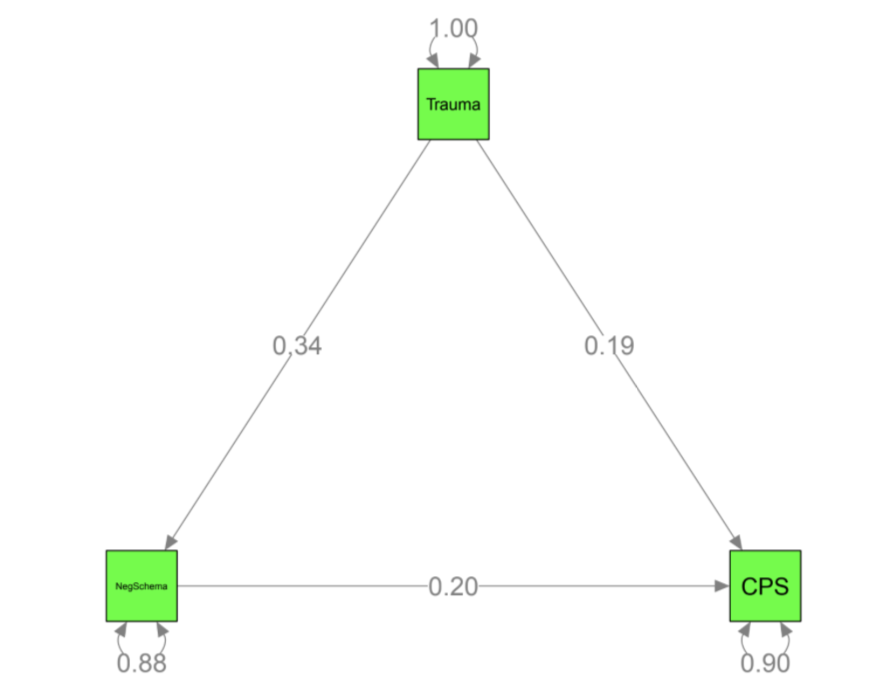

Per−Study Indirect Effects: Trauma −> Neg. Self−Schema −> CPS

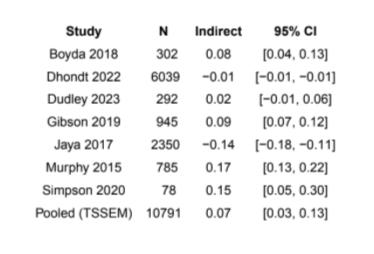

Leave−One−Out Sensitivity Analysis: Results Table

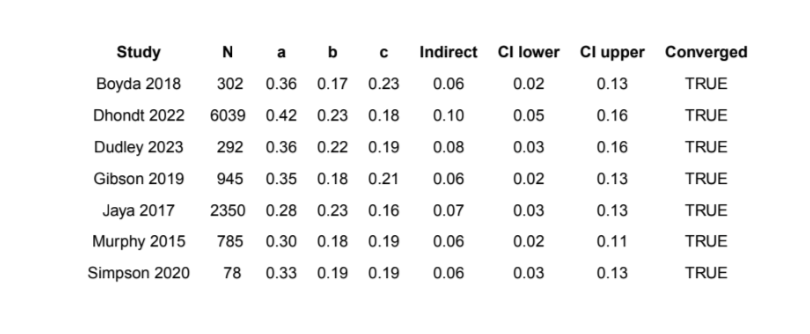

Leave−One−Out Sensitivity Analysis

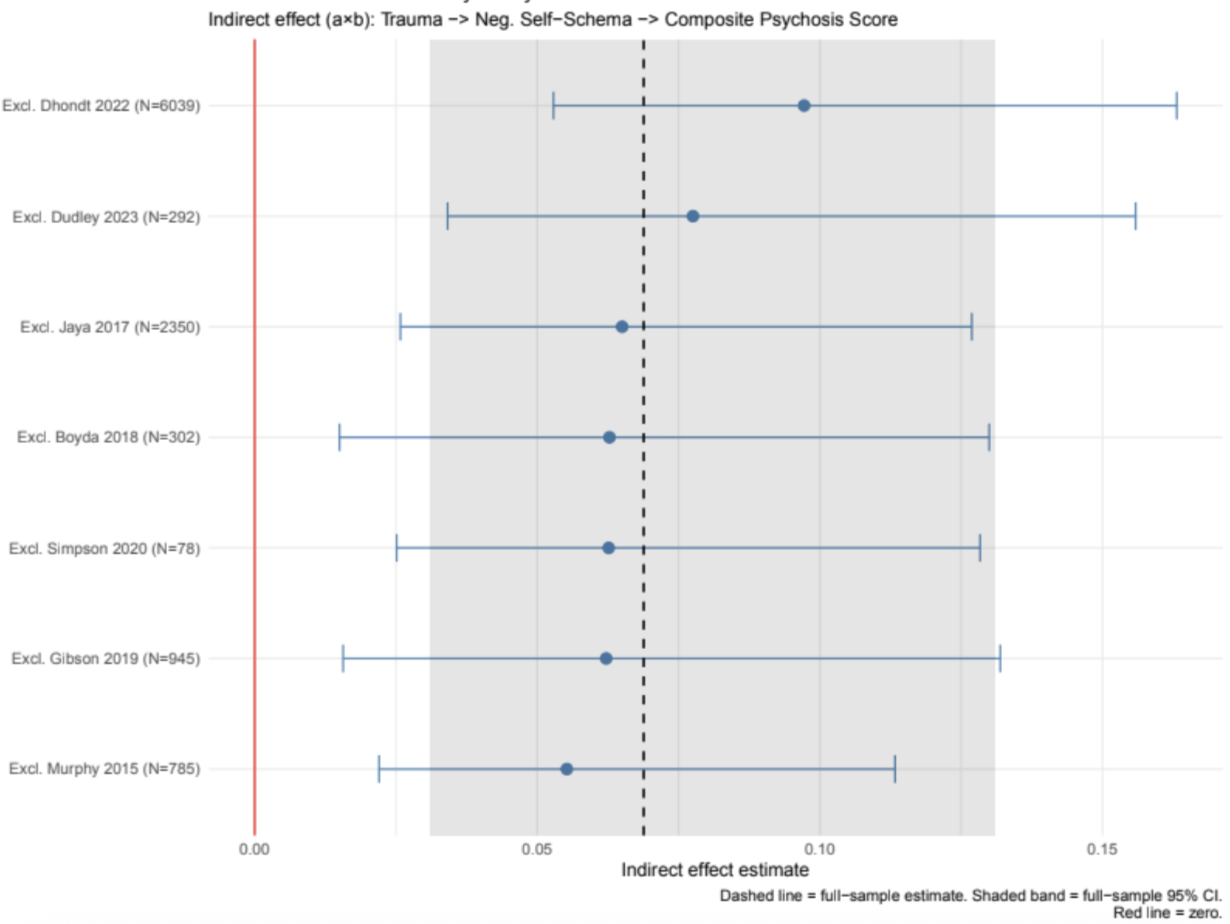

### Delusions Negative Schema Meta-SEM results

Pooled Correlation Matrix (Random−Effects Model)

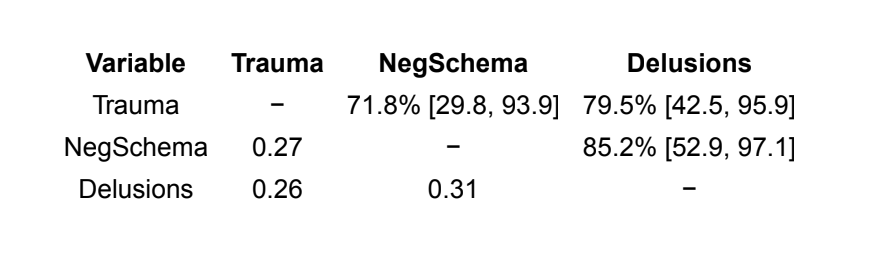

Below diagonal: pooled random−effects correlations. Above diagonal: I² with 95% likelihood−based CI (% of variance due to between−study heterogeneity).

Path Coefficients and Mediation Effects

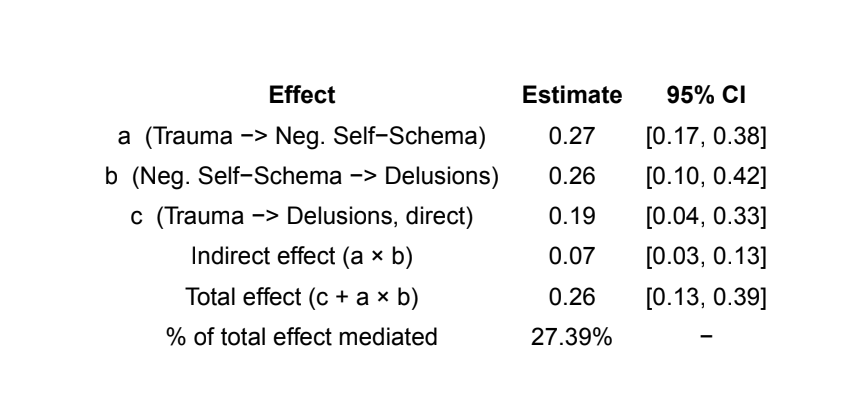

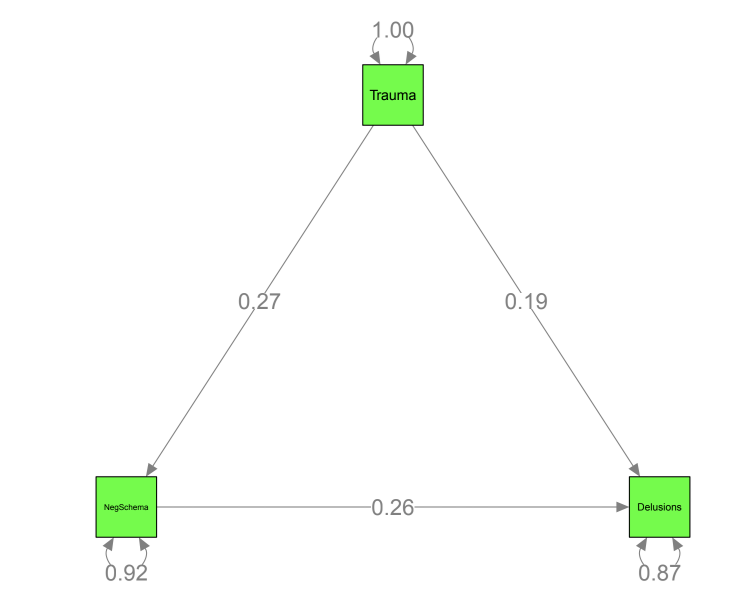

Per−Study Indirect Effects: Trauma −> Neg. Self−Schema −> Delusions

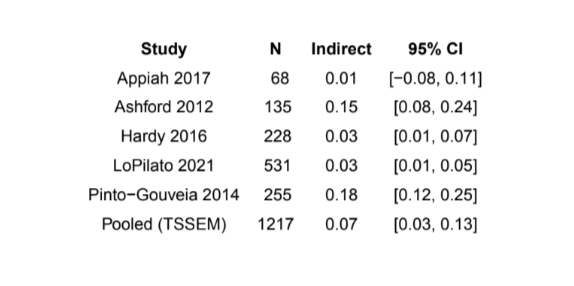

Leave−One−Out Sensitivity Analysis: Results Table

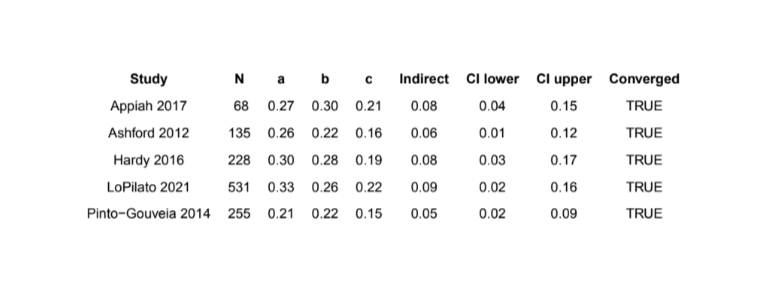

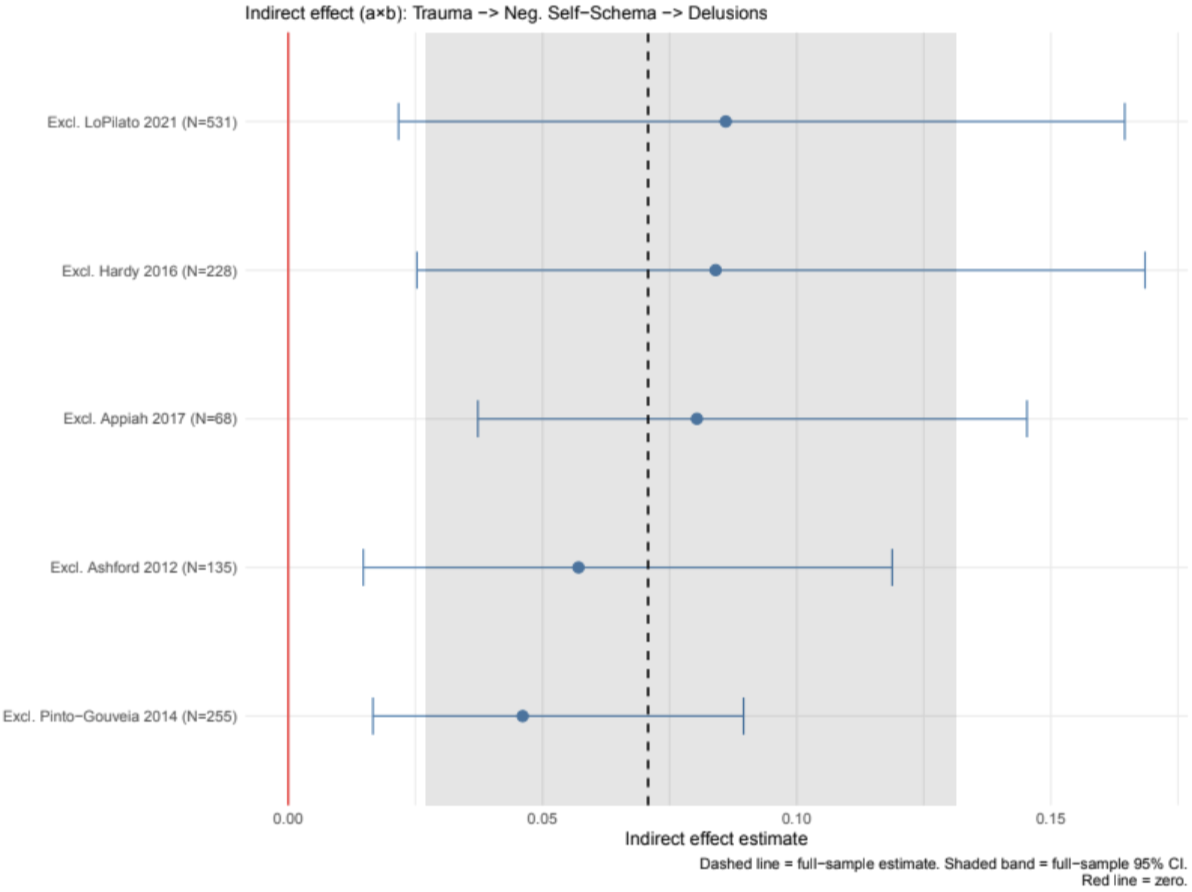

### Composite Depression Meta-SEM results

Pooled Correlation Matrix (Fixed−Effects Model)

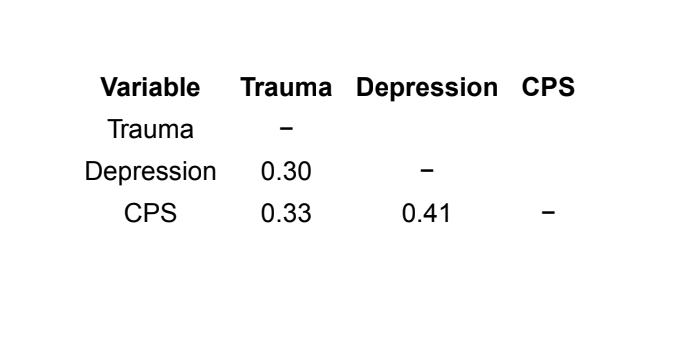

Note. Values below the diagonal are pooled fixed−effects (common−effect) correlations. The upper triangle is

intentionally left blank.

Homogeneity test (fixed−effects): Q(12) = 73.68, p < .001.

A fixed−effects model was used because the random−effects Stage−1 estimation reached the heterogeneity boundary (tau^2

estimates near 0; OpenMx status code 6), so the random−effects solution was unreliable and a common−effect model was

retained.

I2 is not reported: I2 is the proportion of total variance attributable to between−study variance (tau^2), which is

fixed to zero under a fixed−effects model. With no tau^2 to estimate, I2 is undefined; the Q statistic above is the

corresponding test of homogeneity.

Path Coefficients and Mediation Effects

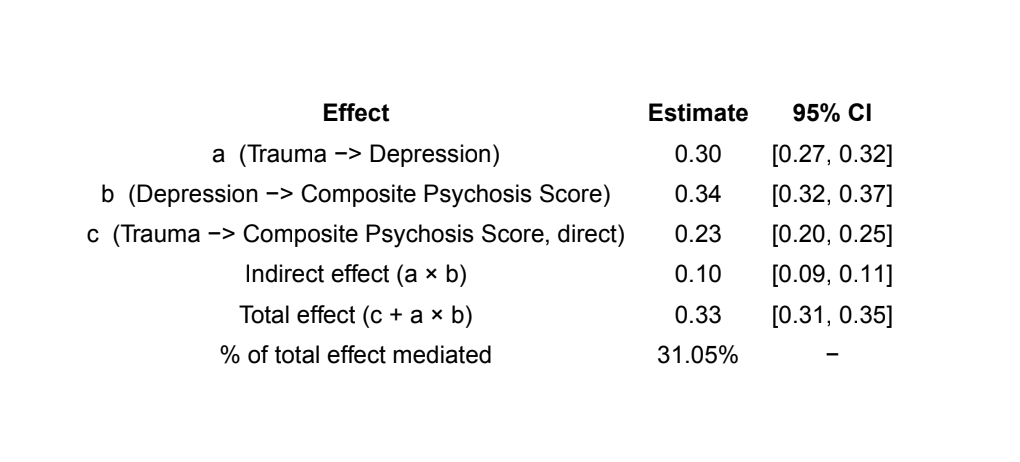

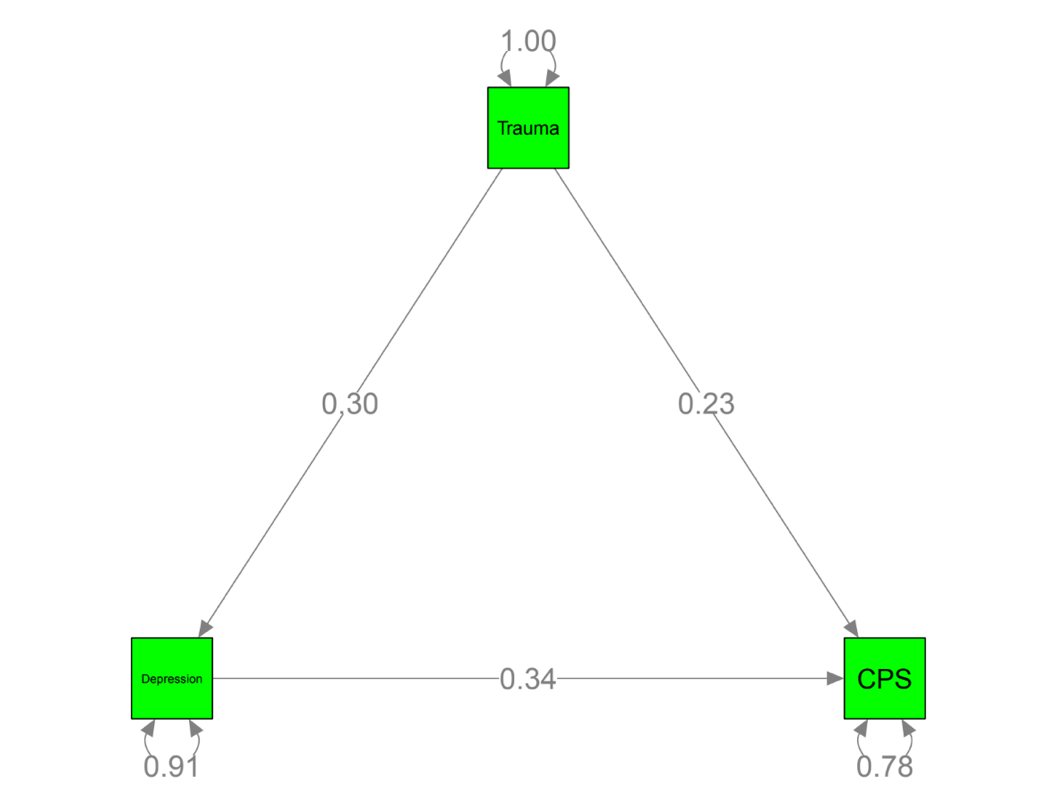

Per−Study Indirect Effects: Trauma −> Depression −> CPS

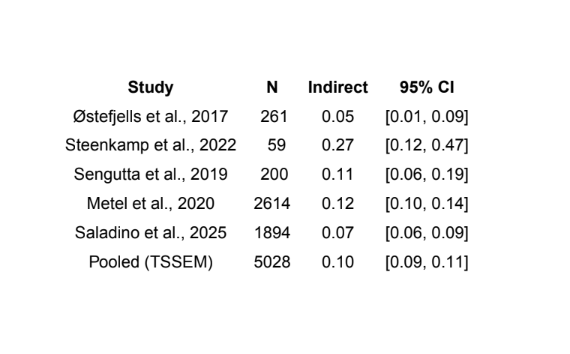

Leave−One−Out Sensitivity Analysis: Results Table

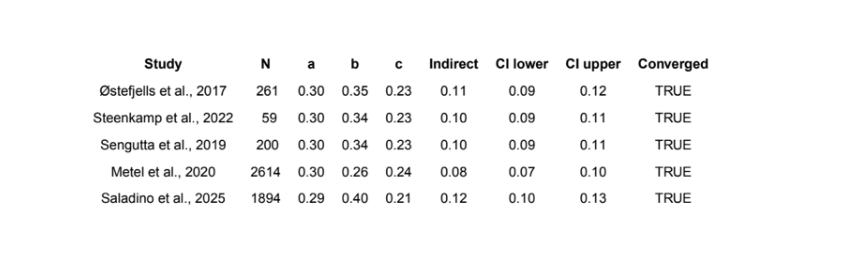

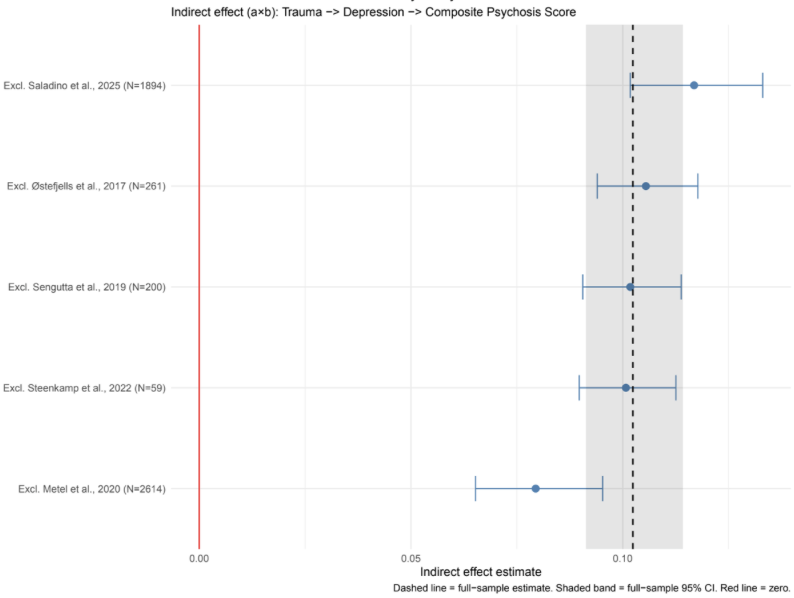
